# A real-world pan-cancer catalogue of mutational signatures from 111,711 tumors

**DOI:** 10.1101/2023.05.18.23290188

**Authors:** Donghyuk Lee, Min Hua, Difei Wang, Lei Song, Tongwu Zhang, Xing Hua, Kai Yu, Xiaohong R. Yang, Stephen J. Chanock, Jianxin Shi, Maria Teresa Landi, Bin Zhu

**Author notes:** Corresponding author: Bin Zhu.

## Abstract

Tumor mutational signatures record the genomic footprints of endogenous and exogenous mutagenic processes and can refine cancer diagnosis and treatment. However, their clinical translation has been limited because most reference catalogues and fitting algorithms are calibrated for whole-exome or whole-genome sequencing, whereas routine diagnostics often use targeted sequencing panels that sample uneven genomic regions. Here we introduce SATS, a computational framework designed for mutational signature analysis in targeted sequencing data. Applying SATS to 111,711 tumors from the AACR Project GENIE, we generated a real-world, panel-calibrated pan-cancer catalogue of mutational signatures for targeted sequencing and evaluated the reproducibility of lung, breast and colorectal signatures in 22,388 additional tumors from a later GENIE release and three external cohorts. Because these tumors were profiled in routine clinical care and linked to clinical records, the resulting catalogue is anchored in a real-world oncology setting rather than in a research-sequencing cohort. The catalogue spans a broad spectrum of cancer types and subtypes, revealing pervasive heterogeneity in signature prevalence. Integrating this catalogue with matched clinical records, we identified signatures enriched in early-onset hypermutated colorectal cancer and signatures associated with cancer prognosis and response to immunotherapy. Together, SATS and this catalogue provide a mutational signature resource for studying cancer etiology, prognosis and treatment response from clinical targeted sequencing.

## Introduction

Somatic mutational signatures capture the characteristic footprints left in cancer genomes by DNA-damage processes, defective repair pathways, treatment and environmental exposures. Large-scale whole-exome and whole-genome sequencing (WES/WGS) studies have catalogued these signatures across cancer types^1,2^ and have driven the development of computational tools for signature extraction and refitting^3–7^. By linking mutational patterns to underlying biological and environmental processes, signature analysis has provided insights into cancer etiology and has shown potential to inform cancer detection^8–10^ and therapeutic stratification^11–14^.

That potential has yet to reach clinical oncology. Although large WES/WGS studies and research-sequencing cohorts such as The Cancer Genome Atlas (TCGA) and Pan-Cancer Analysis of Whole Genomes (PCAWG) study have defined the major mutational signatures in cancer, most diagnostic oncology testing is performed with targeted sequencing panels because they are faster, cheaper, more tolerant of low-cellularity specimens, compatible with clinical turnaround time, and provide deeper coverage of actionable genes than WES/WGS^15–17^. As a result, targeted panels are embedded in real-world oncology workflows and have generated much of the sequencing data linked to clinical decisions and outcomes^18^. Even as larger panels, WES and WGS become more available, existing and ongoing clinical archives contain large numbers of tumors profiled across diverse targeted assays, making methods that model assay-specific coverage necessary for current real-world data and adaptable to future datasets with mixed sequencing breadth. Indeed, these diverse targeted assays interrogate heterogeneous genomic regions and yield sparse mutation counts, making direct application of WES/WGS-calibrated signature catalogues and fitting algorithms unreliable (Supplementary Note 1). A further limitation is that many existing signature resources were developed from research cohorts rather than from routine hospital testing linked to diagnostic, treatment and outcome information. Thus, the field lacks a panel-calibrated mutational-signature catalogue derived from real-world clinical targeted sequencing and connected to matched clinical records. This gap has limited the clinical adoption of mutational signatures.

To fill this gap, we developed SATS (Signature Analyzer for Targeted Sequencing), a computational framework designed specifically for mutational signature analysis in targeted sequencing data. SATS accounts for panel-specific sequence context and mutation opportunity, enabling signature extraction and refitting from sparse, heterogeneous targeted-panel data. We applied SATS to 111,711 tumors profiled by targeted sequencing from the AACR (American Association for Cancer Research) Project GENIE (Genomics Evidence Neoplasia Information Exchange, version 13.0-public), one of the largest real-world clinico-genomic cancer registries^19,20^, to construct a real-world, panel-calibrated pan-cancer catalogue of mutational signatures tailored to clinical targeted sequencing. We further assessed the reproducibility of lung, breast and colorectal signature sets in 13,237 held-out tumors from AACR Project GENIE version 17.0 and 9,151 tumors from independent OrigiMed, Molecular Taxonomy of Breast Cancer International Consortium (METABRIC) and Fudan University Shanghai Cancer Center (FUSCC)/Fudan cohorts^21–23^. A National Cancer Institute (NCI) interactive catalogue browser and a web-based refitting tool were developed to support application of the catalogue to targeted-panel cohorts.

By applying SATS to AACR Project GENIE, we combined panel-calibrated mutational signature analysis with a large real-world clinico-genomic resource in which targeted sequencing is linked to clinical, treatment and outcome information. The resulting catalogue spans 102 OncoTree cancer types, organized into 23 cancer categories used for downstream analysis, and 757 cancer subtypes, enabling mutational signatures to be evaluated across the diversity of tumors encountered in clinical oncology practice. This linkage to detailed clinical and treatment information from the same patients also enables investigation of the etiological, prognostic and therapeutic relevance of targeted sequencing-derived signatures. For example, we found that POLE deficiency-related signatures were enriched in early-onset hypermutated colorectal cancers, and that tumor mutational burden (TMB) attributed to the smoking-related signature, rather than aging-related burden or total TMB, was significantly associated with improved immunotherapy response in non-small cell lung cancer. Together, these analyses show that panel-calibrated mutational signature analysis can extend beyond technical feasibility to clinically interpretable, outcome-linked applications in real-world targeted sequencing data.

## Results

### The AACR Project GENIE Study

We constructed the real-world pan-cancer catalogue of mutational signatures from targeted sequencing data in AACR Project GENIE, a large, multi-institutional clinico-genomic cancer registry. GENIE version 13.0-public includes 111,711 primary, metastatic and unspecified or hematologic tumors profiled by targeted sequencing across 16 participating hospitals and cancer centers (Fig. 1a). Because these data were generated in clinical sequencing practices rather than in a centrally designed research WES/WGS study, GENIE provides a direct setting for evaluating mutational signatures in the targeted-panel data used in routine oncology. The registry also contains matched clinical annotations, with treatment and outcome information available for downstream clinical analyses.

**Fig 1.**
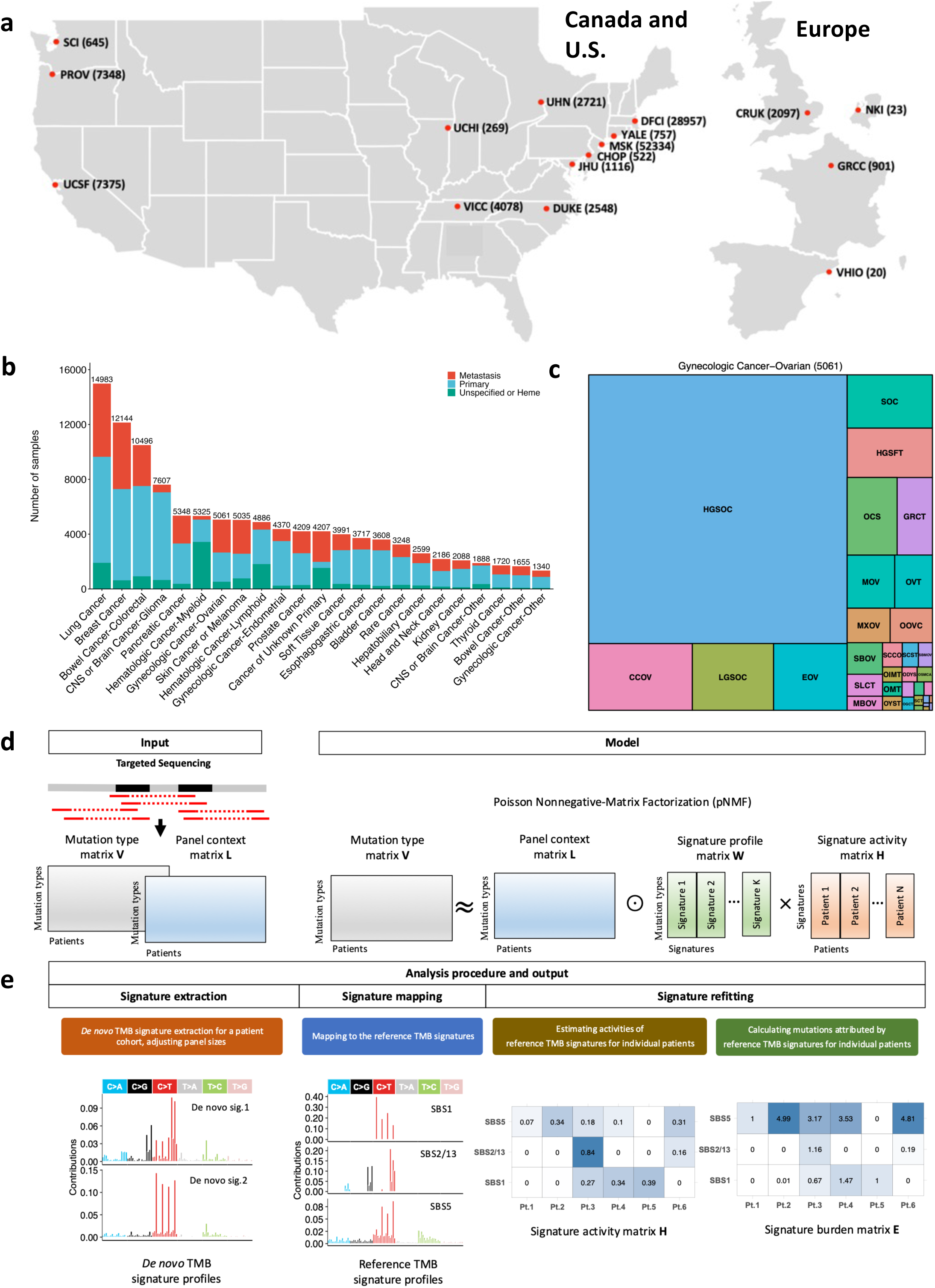
**Overview of the AACR Project GENIE cohort and SATS workflow**. a, Participating AACR Project GENIE sites in the analyzed GENIE version 13.0-public cohort of 111,711 tumors across 16 hospitals and cancer centers; numbers in parentheses indicate tumors contributed by each site. b, Numbers of tumors analyzed across the 23 cancer categories used for downstream analysis, stratified by primary, metastatic and unspecified or hematologic tumor status; numbers above bars indicate cancer-category totals. c, Treemap of ovarian cancer subtypes in AACR Project GENIE. Subtypes were annotated using OncoTree, and tile area is proportional to the number of tumors in each subtype. d, SATS input and panel-calibrated model. Somatic mutations from targeted sequencing are summarized as a mutation-type matrix V, and panel-specific trinucleotide opportunities are summarized as a panel-context matrix L. SATS fits a Poisson non-negative matrix factorization (pNMF) model to estimate TMB-normalized signature profiles and tumor-level signature activities while incorporating panel-specific mutation opportunities. e, SATS analysis workflow. SATS extracts de novo TMB-normalized signatures from grouped targeted-sequencing samples, maps de novo profiles to COSMIC v3.4 TMB reference signatures by recurrent mapping, and refits mapped signatures in individual tumors using an expectation-maximization algorithm to estimate signature activities and signature-attributed mutation burdens. Site abbreviations: CHOP, Children’s Hospital of Philadelphia; CRUK, Cancer Research UK Cambridge Centre; DFCI, Dana-Farber Cancer Institute; DUKE, Duke Cancer Institute; GRCC, Gustave Roussy Cancer Campus; JHU, Johns Hopkins Sidney Kimmel Comprehensive Cancer Center; MSK, Memorial Sloan Kettering Cancer Center; NKI, Netherlands Cancer Institute; PROV, Providence Health & Services Cancer Institute; SCI, Swedish Cancer Institute; UCHI, University of Chicago Comprehensive Cancer Center; UCSF, University of California, San Francisco; UHN, Princess Margaret Cancer Centre, University Health Network; VHIO, Vall d’Hebron Institute of Oncology; VICC, Vanderbilt-Ingram Cancer Center; YALE, Yale Cancer Center. Ovarian cancer subtype abbreviations: CCOV, Clear Cell Ovarian Cancer; EOV, Endometrioid Ovarian Cancer; GRCT, Granulosa Cell Tumor; HGSFT, High-Grade Serous Fallopian Tube Cancer; HGSOC, High-Grade Serous Ovarian Cancer; LGSOC, Low-Grade Serous Ovarian Cancer; MBOV, Mucinous Borderline Ovarian Tumor; MOV, Mucinous Ovarian Cancer; MXOV, Mixed Ovarian Carcinoma; OCS, Ovarian Carcinosarcoma/Malignant Mixed Mesodermal Tumor; ODYS, Dysgerminoma; OGCT, Ovarian Germ Cell Tumor; OIMT, Immature Teratoma; OMT, Mature Teratoma; OOVC, Ovarian Cancer, Other; OSMCA, Ovarian Seromucinous Carcinoma; OVT, Ovarian Epithelial Tumor; OYST, Yolk Sac Tumor; SBMOV, Serous Borderline Ovarian Tumor, Micropapillary; SBOV, Serous Borderline Ovarian Tumor; SCCO, Small Cell Carcinoma of the Ovary; SCST, Sex Cord Stromal Tumor; SCT, Steroid Cell Tumor, NOS; SLCT, Sertoli-Leydig Cell Tumor; SOC, Serous Ovarian Cancer.

The GENIE cohort encompasses 102 cancer types, which we organized into 23 cancer categories used for downstream analysis, including 14,983 lung and 12,144 breast tumors (Fig. 1b). It also includes 757 cancer subtypes defined by OncoTree^24^, providing subtype granularity that is important for clinically ascertained cohorts, where histologic representation can differ from research WES/WGS studies. For example, while the TCGA ovarian cancer WES study^25^ predominantly features high-grade serous ovarian cancer, the GENIE cohort comprises 34 ovarian cancer subtypes, including high-grade serous ovarian cancer, clear cell ovarian cancer, low-grade serous ovarian cancer, and endometrioid ovarian cancer (Fig. 1c). Together, the scale, clinical ascertainment and subtype diversity of GENIE make it a strong foundation for building a broad, panel-calibrated mutational-signature catalogue for real-world targeted-sequencing data.

### Mutational Signature Analysis using SATS

To build this catalogue, we developed SATS, a computational framework designed for targeted sequencing that detects mutational signatures in targeted-sequencing cohorts and estimates their activities in individual tumors. Although this study focuses primarily on single base substitution (SBS) signatures, SATS also supports double base substitution (DBS) analysis, which we used to generate a DBS catalogue.

SATS operates on a mutation type matrix **V** that contains the counts of SBS across 96 mutation types defined by 32 trinucleotide contexts, such as a C to G substitution at the trinucleotide context TCT (represented as T[C>G]T). To adjust for differences in targeted genomic regions across panels, SATS incorporates a panel context matrix **L**, which specifies the number of trinucleotide contexts in the panel where each mutation type (e.g., TCT for T[C>G]T substitutions) could potentially occur (Supplementary Note 2). Accordingly, SATS estimates tumor-mutational-burden (TMB) signature profiles rather than raw tumor-mutational-count (TMC) profiles (Supplementary Note 3). SATS uses a Poisson non-negative matrix factorization (pNMF) model to decompose the mutation type-by-patient matrix **V** into a mutation type-by-signature matrix **W** representing signature profiles and a signature-by-patient matrix **H** quantifying signature activities, while explicitly adjusting for the panel-specific context matrix **L** (Fig. 1d).

SATS proceeds in three main steps: signature extraction, signature mapping and sample-level signature refitting (Fig. 1e). For signature extraction, SATS applies the context-adjusted pNMF model implemented in signeR^4^ to identify *de novo* signatures from grouped targeted-sequencing samples. Because sparse targeted-panel data can yield blended *de novo* profiles, for example profiles that merge SBS1, SBS2/13 and SBS5, SATS next maps each *de novo* profile to TMB reference signatures from the Catalogue of Somatic Mutations in Cancer (COSMIC) v3.4 pan-cancer catalogue^1^ (Fig. 1e and Supplementary Fig. 1). This mapping is performed with penalized non-negative least squares (pNNLS)^26^ and L1 penalization, retaining only reference signatures with recurrent, substantial contributions across repeated mappings. This recurrence filter allows SATS to evaluate the full COSMIC catalogue while producing a concise, data-supported signature set for downstream refitting. *De novo* profiles with poor COSMIC mapping were not treated as new signatures; only recurrent COSMIC-mapped signatures were retained for individual-tumor refitting.

After signature mapping, we developed a panel-calibrated expectation-maximization (EM) algorithm to estimate signature activities in individual tumors. In addition, this refitting step computes signature burden, defined as the expected number of mutations attributed to each mapped signature in each sample. Although signature refitting methods have been described previously^6,7^, most were developed for WES/WGS and do not explicitly model the variable trinucleotide context matrices induced by targeted panels. signeR incorporates panel size during signature extraction, but its refitting step uses a Bayesian prior with fixed hyperparameters that can influence signature activity estimates when mutation counts are sparse. In contrast, SATS estimates non-negative activities by maximizing the panel-calibrated Poisson likelihood without fixed exposure priors, reducing reliance on prior assumptions. In simulations across four cancer types, SATS EM refitting achieved lower mean squared refitting errors than signeR refitting across most signatures and cancer types (Supplementary Fig. 2).

We evaluated factors influencing SATS performance using simulated data and pseudo-targeted sequencing data generated by down-sampling WES and WGS datasets. Signature detection and burden estimation improved when signatures had more distinctive, less-flat profiles, higher prevalence and larger sequencing panels (>1 Mb) (Supplementary Note 4). In breast cancer simulations, increasing the number of tumors enabled most common signatures to be detected with very low false discovery rates, including SBS3, a therapeutically important but difficult-to-detect signature associated with homologous recombination deficiency (HRD) (Supplementary Note 5).

We then benchmarked SATS by separating two tasks that are often conflated in mutational signature analysis: *de novo* identification of active signatures in targeted-sequencing cohorts and refitting of those signatures to estimate tumor-level burdens. For active-signature detection, SATS recovered the prespecified active signatures more consistently than SigProfilerExtractor^5^ and Mix^27^ when applied to the same targeted-panel simulations (Supplementary Note 6 and Supplementary Fig. 3). For burden estimation, SATS used recurrent pNNLS mapping to convert d*e novo* profiles into a restricted set of reference signatures, followed by panel-calibrated EM refitting. This constrained reference-set strategy was important: in breast cancer simulations, supplying a broad WES-derived reference set introduced false attributions to signatures absent from the simulated tumors, whereas SATS maintained accurate burden estimates once a targeted-panel-supported reference set was used, including in subset and single-sample lung simulations (Supplementary Notes 7, 9 and 10 and Supplementary Figs. 4, 5 and 6). These benchmarks support SATS as an integrated workflow combining d*e novo* signature detection, recurrent reference-signature mapping and panel-calibrated refitting.

We next evaluated SATS in an orthogonal paired-platform benchmark using 72 tumors profiled by both WGS and targeted sequencing. We compared SATS with SigProfilerAssignment^7^, Mix^27^ and deconstructSigs^6^ using a restricted set of common kidney-cancer signatures, and compared targeted-sequencing burdens with matched WGS burdens. For SBS1, SATS showed the strongest continuous concordance with WGS burden, whereas other methods produced weak or non-estimable sample-level associations. For the high-burden flat SBS5/SBS40 signatures, all methods showed positive concordance with WGS burden (Supplementary Note 8 and Supplementary Fig. 7). This paired-platform benchmark indicates that SATS better preserves lower-prevalence signature burdens when targeted-panel mutation counts are sparse.

Together, these simulation and paired-platform benchmarks show that targeted-sequencing signature analysis requires a panel-calibrated workflow rather than direct transfer of an unrestricted WES/WGS signature catalogue. By combining *de novo* detection, recurrent COSMIC signature mapping and panel-calibrated refitting, SATS defines a data-supported reference set for targeted sequencing, providing the basis for the pan-cancer catalogue described below.

### The pan-cancer catalogue of targeted-sequencing-derived mutational signatures

The catalogue comprises 26 SBS signatures identified from 111,711 tumors profiled by targeted sequencing in AACR Project GENIE. Figure 2a summarizes each signature by signature-attributed tumor mutational burden and prevalence across cancer types. Broadly recurrent signatures included SBS1, caused by deamination of 5-methylcytosine, and flat-profile signatures including SBS3, SBS5, SBS40a and SBS40b, which were observed across many cancer types and accounted for substantial mutation burden in several cohorts. Other signatures showed more restricted cancer-type distributions. Together, these patterns show that the targeted-sequencing catalogue captures both common background mutational processes and cancer-type-enriched etiologic signals in real-world oncology cohorts.

**Fig 2.**
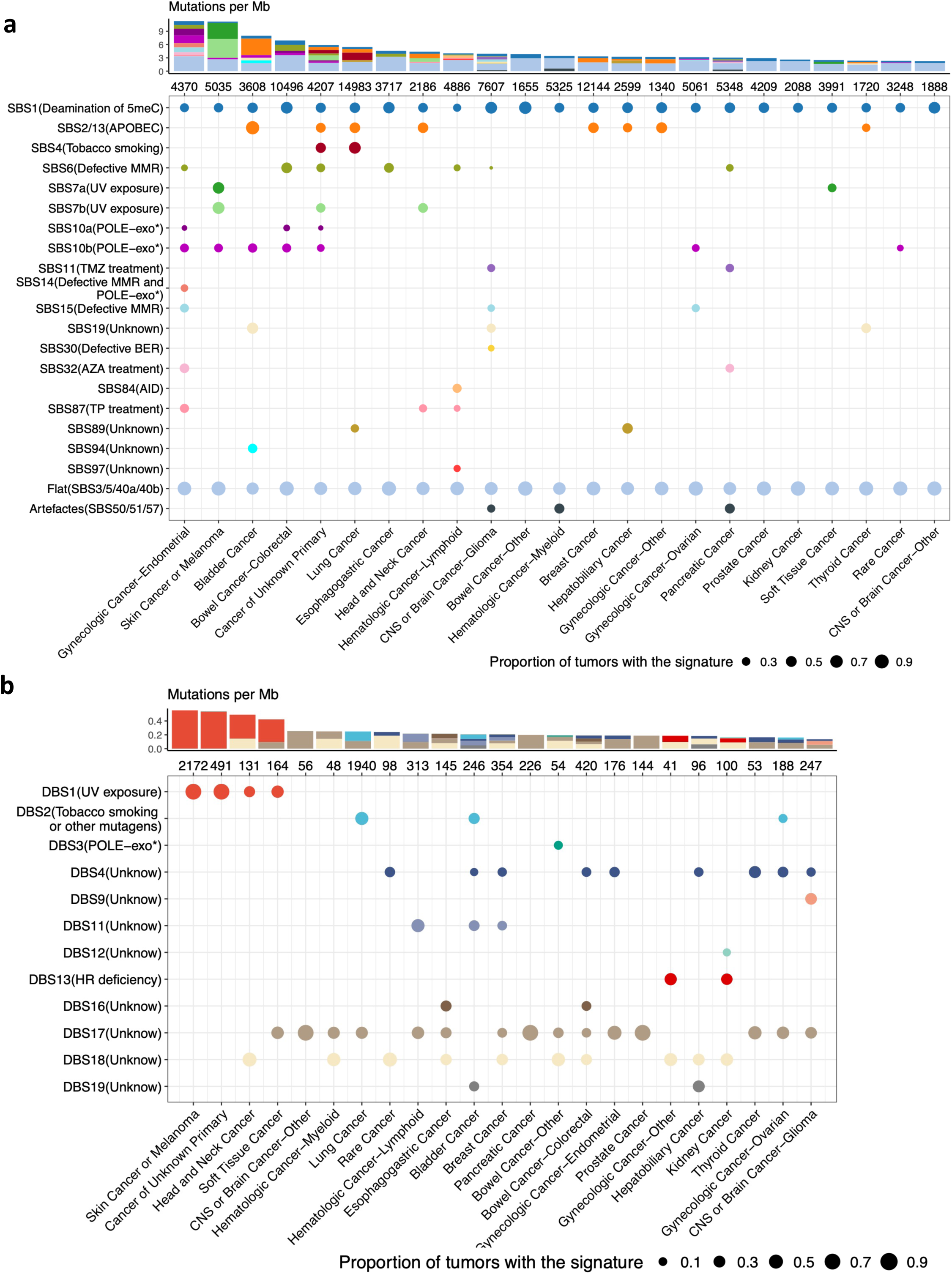
**Pan-cancer catalogue of targeted-sequencing-derived mutational signatures**. a, Single base substitution (SBS) signatures. b, Double base substitution (DBS) signatures. The SBS catalogue includes 26 signatures identified from 982,095 SBSs across 111,711 tumors; the DBS catalogue was generated from 15,149 DBSs in the same cohort. For each panel, the top stacked bar chart shows panel-calibrated signature-attributed TMB (mutations/Mb) for each cancer category, with colors corresponding to signatures. Numbers below bars indicate tumors analyzed in each cancer category. Dot plots show signature prevalence across cancer categories; dot area is proportional to the fraction of tumors in which the signature was detected by SATS refitting. Signature labels list the COSMIC signature or composite signature group and proposed etiology. SBS and DBS signatures were analyzed separately because they use different mutation-type vocabularies and context-opportunity matrices. Composite entries include SBS2/13, flat signatures (SBS3/SBS5/SBS40a/SBS40b) and artefact signatures (SBS50/SBS51/SBS57). Abbreviations: 5meC, 5-methylcytosine; AID, activation-induced deaminase; APOBEC, apolipoprotein B mRNA-editing enzyme catalytic polypeptide; AZA, azathioprine; BER, base-excision repair; HR, homologous recombination; MMR, mismatch repair; POLE-exo*, mutations in the polymerase epsilon exonuclease domain; TMZ, temozolomide; TP, thiopurine; UV, ultraviolet radiation.

The cancer-type-enriched SBS signatures recapitulated several expected biological and exposure-related mutational processes. Endometrial cancers were enriched for mismatch repair deficiency signatures, including SBS6, SBS14 and SBS15, and for POLE-related signatures SBS10a and SBS10b. APOBEC-associated signatures SBS2/13 were most frequent in bladder cancer and were also prominent in lung cancer, where tobacco-associated SBS4 was also detected. UV-associated signatures SBS7a and SBS7b were observed in skin cancers, including melanoma, and in additional cancer groups with plausible ultraviolet exposure contributions, including head and neck cancer, soft tissue cancer and cancers of unknown primary. Treatment-associated signatures were also detected, including SBS11, associated with temozolomide exposure, in glioma^28^ and pancreatic cancer^29^; SBS32, associated with azathioprine exposure, in pancreatic cancer; and SBS87, associated with thiopurine treatment, in lymphoid-derived hematologic cancer^30^.

Because AACR Project GENIE is a clinically ascertained targeted-sequencing cohort, signatures in this catalogue should be interpreted as representations of known mutational processes in real-world oncology practice rather than as *de novo* discovery of new processes. SBS32 illustrates this point. In our catalogue, SBS32 was detected in pancreatic cancer, whereas it was originally described in cutaneous squamous cell carcinoma^31^ and was not reported as a pancreatic cancer signature in prior WGS catalogues^1^. Its presence may reflect treatment exposure, subtype composition or other features of the clinically sequenced population. Prior studies linking azathioprine use to acute pancreatitis^32,33^, a known risk factor for pancreatic cancer^34,35^, provide relevant clinical context, but do not establish a causal explanation for SBS32 in pancreatic tumors. Similarly, SBS6 was detected in colorectal cancer, esophagogastric cancer and glioma in our targeted-sequencing catalogue, whereas it was not reported for the same cancer types in the PCAWG WGS study^1^. These examples suggest that clinical ascertainment, treatment history, subtype composition and sampling patterns can influence where known signatures are observed across cancer types.

We also generated a pan-cancer DBS signature catalogue for tumors profiled by targeted sequencing (Fig. 2b). Although DBS burdens were generally low, as expected given the smaller number of double-base substitutions captured by targeted panels, several DBS signatures showed cancer-type patterns consistent with known etiologies. DBS1, associated with UV exposure, was present in head and neck cancer, skin cancer or melanoma, soft tissue cancer and cancers of unknown primary, paralleling the SBS UV signatures observed in these cancer types. DBS2, associated with tobacco smoking and other mutagens, was identified in bladder, ovarian and lung cancers. We also observed DBS3, associated with POLE-related replication repair deficiency, in non-colorectal bowel cancer, and DBS13, linked to homologous recombination deficiency, in kidney cancer and gynecologic malignancies excluding endometrial and ovarian cancers. Other DBS signatures remain of uncertain etiology. Together, the SBS and DBS catalogues show that real-world targeted-panel sequencing captures a clinically grounded spectrum of known mutational processes, ranging from ubiquitous background signatures to cancer-type-enriched endogenous, environmental and treatment-associated signatures.

### Reproducibility of targeted-sequencing-derived mutational signatures

We assessed the reproducibility of the targeted sequencing-derived, panel-calibrated signature catalogue using a held-out AACR Project GENIE version 17.0 cohort and three external targeted-sequencing datasets. This analysis tested whether the panel-calibrated signatures identified in GENIE version 13.0-public could be detected again in independent cohorts analyzed with the same SATS framework.

In the held-out GENIE version 17.0 cohort, comprising 13,237 tumors (6,405 lung, 4,128 colorectal and 2,704 breast tumors) that were absent from GENIE version 13.0-public, SATS recapitulated the expected GENIE version 13.0-public catalogue signatures in all three validation cancer types (Fig. 3a). Lung cancer showed SBS1, APOBEC-associated SBS2/13, tobacco-related SBS4/SBS29, SBS89 and flat signatures; breast cancer showed SBS1, SBS2/13 and flat signatures; and colorectal cancer showed SBS1, mismatch-repair-related SBS6, POLE-related SBS10a/SBS10b and flat signatures. An additional signature, SBS15 in colorectal cancer, was also observed in the held-out cohort. Signature SBS15 is biologically plausible but was not recovered with the same frequency or robustness as the core GENIE version 13.0-public catalogue signatures. We therefore treated it as a suggestive validation-cohort signature rather than the catalogue-defining signature, preserving a conservative catalogue focused on signatures repeatedly detected across independent analyses.

**Fig 3.**
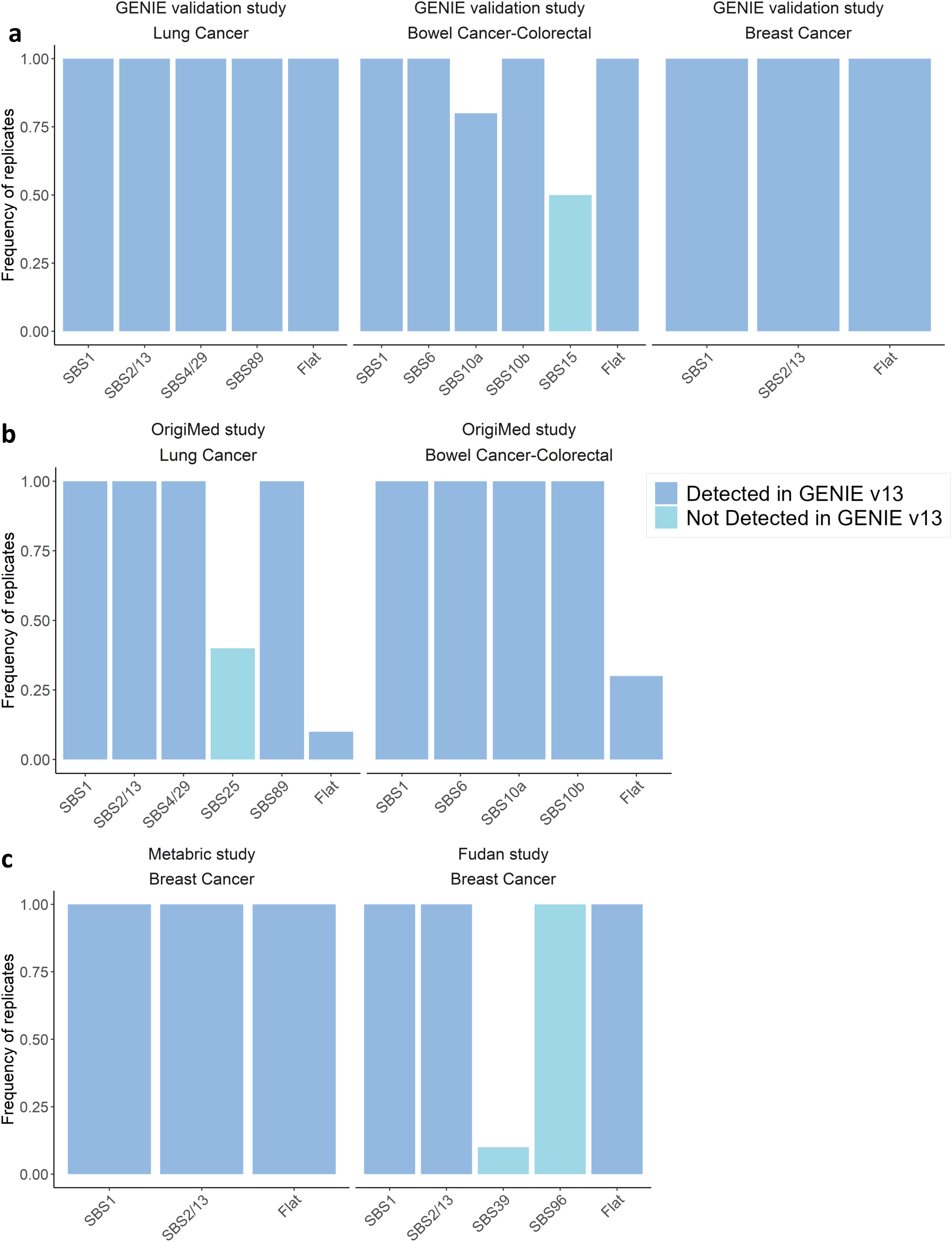
**Reproducibility assessment of targeted-sequencing-derived mutational signatures**. Bar plots show the fraction of ten repeated SATS validation runs, using different random seeds and 50-tumor sample groupings, in which each signature was recovered. a, Held-out AACR Project GENIE version 17.0 tumors absent from GENIE version 13.0-public (n = 13,237; 6,405 lung, 4,128 colorectal and 2,704 breast tumors). b, External OrigiMed lung (n = 2,143) and colorectal (n = 1,206) cohorts. c, External breast cancer cohorts from METABRIC (n = 2,346) and FUSCC/Fudan (n = 3,456). Blue bars denote signatures detected in the GENIE version 13.0-public catalogue; cyan bars denote validation-cohort signals not detected in GENIE version 13.0-public.

We next evaluated reproducibility in three external targeted-sequencing datasets: OrigiMed for lung and colorectal cancer, and METABRIC and FUSCC/Fudan for breast cancer. In OrigiMed, an Asian cohort with 2,143 lung and 1,206 colorectal tumors after outlier filtering, SATS detected the dominant GENIE version 13.0-public lung catalogue signatures SBS1, SBS2/13, SBS4/SBS29 and SBS89, as well as the colorectal catalogue signatures SBS1, SBS6, SBS10a and SBS10b (Fig. 3b). Breast cancer reproducibility was evaluated in METABRIC (n = 2,346) and FUSCC/Fudan (n = 3,456), because OrigiMed included only 306 breast tumors; both breast cohorts recovered the GENIE version 13.0-public breast catalogue signatures SBS1, SBS2/13 and flat-signature groups (Fig. 3c). Additional signals, including SBS39/SBS96 in FUSCC/Fudan and SBS25 in OrigiMed lung, were detected in individual external cohorts but were not recurrent across validation datasets. These validation-only findings may reflect cohort-specific exposures, treatment histories, ancestry, case composition or panel design, but they did not expand the core catalogue without repeated validation. Overall, the internal and external validations show that most panel-calibrated signatures are reproducibly detected across independent targeted-sequencing cohorts, supporting a conservative catalogue built around frequently validated signatures.

### Heterogeneous prevalence of mutational signatures between and within cancer types

Having established the reproducibility of the targeted sequencing-derived catalogue, we next asked whether these signatures captured biologically meaningful heterogeneity across tumors. Compared with WES/WGS-based mutational signature catalogues, this targeted-sequencing-derived catalogue includes a broader range of cancer types and subtypes, enabling systematic evaluation of SBS signature patterns in real-world clinical cohorts. In tumors from Memorial Sloan Kettering Cancer Center, clustering based on signature burdens separated tumors largely by cancer type: lung tumors formed one distinct cluster, whereas tumors with UV-associated signatures, including skin cancer or melanoma and subsets of head and neck cancer, formed another (Fig. 4a, Methods). Similar clustering patterns were observed in tumors sequenced at Dana-Farber Cancer Institute (Supplementary Fig. 8). Tumors of unknown primary clustered with lung tumors, UV-signature-enriched tumors, glioma and pancreatic tumors, suggesting that signature burden profiles may provide complementary information relevant to tissue-of-origin assessment, although this observation requires dedicated validation.

**Fig 4.**
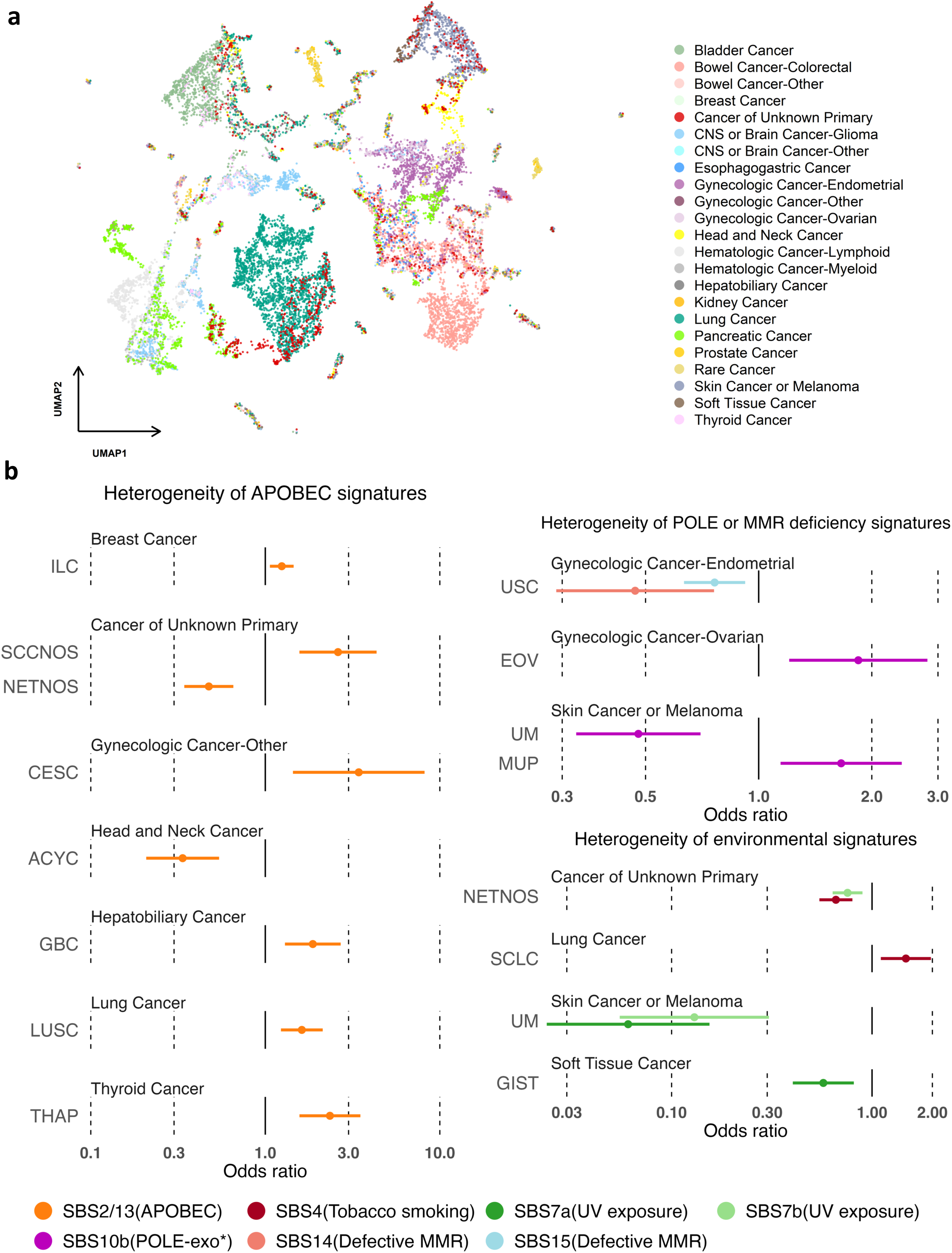
**Heterogeneous prevalence of mutational signatures across tumor types and subtypes**. a, Two-dimensional UMAP (Uniform Manifold Approximation and Projection) embedding of SBS signature-burden profiles for tumors sequenced at Memorial Sloan Kettering Cancer Center (n = 52,334). Within each cancer category, tumors were clustered by signature-burden profiles, and each point represents the median signature-burden profile of a tumor cluster with similar signature composition, colored by cancer category. b, Subtype-specific enrichment or depletion of APOBEC, POLE or MMR deficiency, and environmental signatures. Points and horizontal bars denote adjusted odds ratios (ORs) and 95% confidence intervals for SBS signature presence in each cancer subtype, estimated relative to the corresponding cancer-category background. Signature presence was modeled using generalized linear mixed models adjusted for age, self-reported race/ancestry, sex where applicable and tumor status, with tumor subtype and sequencing center as random effects; only significant subtype-signature associations are shown (FDR < 0.1). Values to the right of one indicate enrichment, and values to the left of one indicate depletion. Cancer subtype abbreviations: ACYC, Adenoid Cystic Carcinoma; CESC, Cervical Squamous Cell Carcinoma; EOV, Endometrioid Ovarian Cancer; GBC, Gallbladder Cancer; GIST, Gastrointestinal Stromal Tumor; ILC, Invasive Lobular Carcinoma (Breast); LUSC, Lung Squamous Cell Carcinoma; MUP, Melanoma of Unknown Primary; NETNOS, Neuroendocrine Tumor, Not Otherwise Specified; SCCNOS, Squamous Cell Carcinoma, Not Otherwise Specified; SCLC, Small Cell Lung Cancer; THAP, Anaplastic Thyroid Cancer; UM, Uveal Melanoma; USC, Uterine Serous Carcinoma/Uterine Papillary Serous Carcinoma.

The extensive subtype coverage of the catalogue further enabled fine-scale analysis of within-cancer heterogeneity. We first examined signatures linked to endogenous DNA-repair processes, including mismatch repair and POLE deficiency. Among endometrial cancers, Uterine Serous Carcinoma (USC), a non-endometrioid subtype, showed significantly lower prevalence of mismatch repair- or POLE-deficiency-related signatures than other subtypes (SBS14: OR = 0.47, 95% CI = 0.29-0.76; SBS15: OR = 0.76, 95% CI = 0.63-0.92). Conversely, Endometrioid Ovarian Cancer (EOV), which frequently co-occurs with endometrioid endometrial carcinoma^36^, was enriched for the POLE-deficiency-related signature SBS10b relative to other ovarian cancer subtypes (OR = 1.84, 95% CI = 1.21-2.81, Fig. 4b). SBS10b was also elevated in Melanoma of Unknown Primary (MUP) (OR = 1.66, 95% CI = 1.14-2.40), but depleted in Uveal Melanoma (UM) (OR = 0.48, 95% CI = 0.33-0.70). Together, these patterns indicate that mismatch repair and POLE-related mutagenesis varies substantially across histologic and clinical subtypes.

Subtype-specific heterogeneity was also evident for APOBEC-associated signatures SBS2 and SBS13, two common endogenous signatures in cancer^37–39^. Consistent with prior reports^37^, Cervical Squamous Cell Carcinoma (CESC) showed higher SBS2/13 prevalence than other non-endometrial and non-ovarian gynecologic cancers (OR = 3.43, 95% CI = 1.44-8.17, Fig. 4b).

Within head and neck cancers, SBS2/13 was significantly depleted in Adenoid Cystic Carcinoma (ACYC) (OR = 0.34, 95% CI = 0.21-0.54), consistent with biological differences between salivary-gland tumors and more typical squamous epithelial tumors. SBS2/13 was also enriched in Lung Squamous Cell Carcinoma (LUSC) (OR = 1.62, 95% CI = 1.23-2.13), Gallbladder Cancer (GBC)^40^ (OR = 1.87, 95% CI = 1.30-2.70), and Anaplastic Thyroid Cancer (THAP)^41,42^ (OR = 2.35, 95% CI = 1.57-3.51) within lung, hepatobiliary and thyroid cancers, respectively. These findings indicate that APOBEC mutagenesis is not uniformly distributed within broad cancer subtypes, but is concentrated in specific histologic and clinical subgroups.

Environmental exposure signatures showed similarly interpretable subtype-specific patterns. UV-induced signatures SBS7a and SBS7b were markedly depleted in Uveal Melanoma compared with other skin cancer or melanoma subtypes (SBS7a: OR = 0.06, 95% CI = 0.02-0.15; SBS7b: OR = 0.13, 95% CI = 0.06-0.31, Fig. 4b), consistent with the low sunlight exposure of uveal tissue^43^. The smoking-associated signature SBS4 was enriched in Small Cell Lung Cancer (SCLC) (OR = 1.48, 95% CI = 1.11-1.97), consistent with the strong tobacco-smoking etiology of this subtype^44^.

Together, these analyses show that targeted sequencing-derived mutational signatures capture both between-cancer and within-cancer heterogeneity in real-world clinical cohorts. The catalogue reveals variation in endogenous DNA-repair processes, APOBEC activity and environmental mutagen exposure across cancer types and subtypes, providing a basis for interpreting etiologic differences across tumors and motivating downstream analyses of clinical associations.

### Targeted sequencing-based mutational signatures associated with clinical outcomes

A key advantage of this real-world targeted-sequencing catalogue is the availability of matched clinical data, which allowed us to relate signature-attributed mutation burden to disease presentation, prognosis and treatment response. We illustrate this utility through three analyses: early-onset colorectal cancer, overall survival across cancer types and immune checkpoint inhibitor response in non-small cell lung cancer.

#### Mutational signatures enriched in early-onset hypermutated colorectal cancer

Previous work reported that early-onset non-hypermutated colorectal cancers had lower overall TMB than late-onset cases, whereas early-onset hypermutated cases showed the opposite pattern^45^. To examine whether specific mutational processes contributed to this difference, we analyzed TMB attributed to individual signatures in non-hypermutated and hypermutated colorectal cancers (Methods). In non-hypermutated tumors, signature-attributed TMBs were collectively associated with lower odds of early-onset disease (OR = 0.898, 95% CI = 0.874-0.922, Fig. 5a left panel). In hypermutated tumors, early-onset cases showed higher total TMB than late-onset cases (OR = 1.004, 95% CI = 1.001-1.007, Fig. 5a right panel), with the largest positive point estimate for the POLE-deficiency-related signature SBS10a (OR = 1.012, 95% CI = 0.995-1.028). These results identify POLE-related SBS10a as a candidate contributor to the elevated mutation burden observed in early-onset hypermutated colorectal cancer.

**Fig 5.**
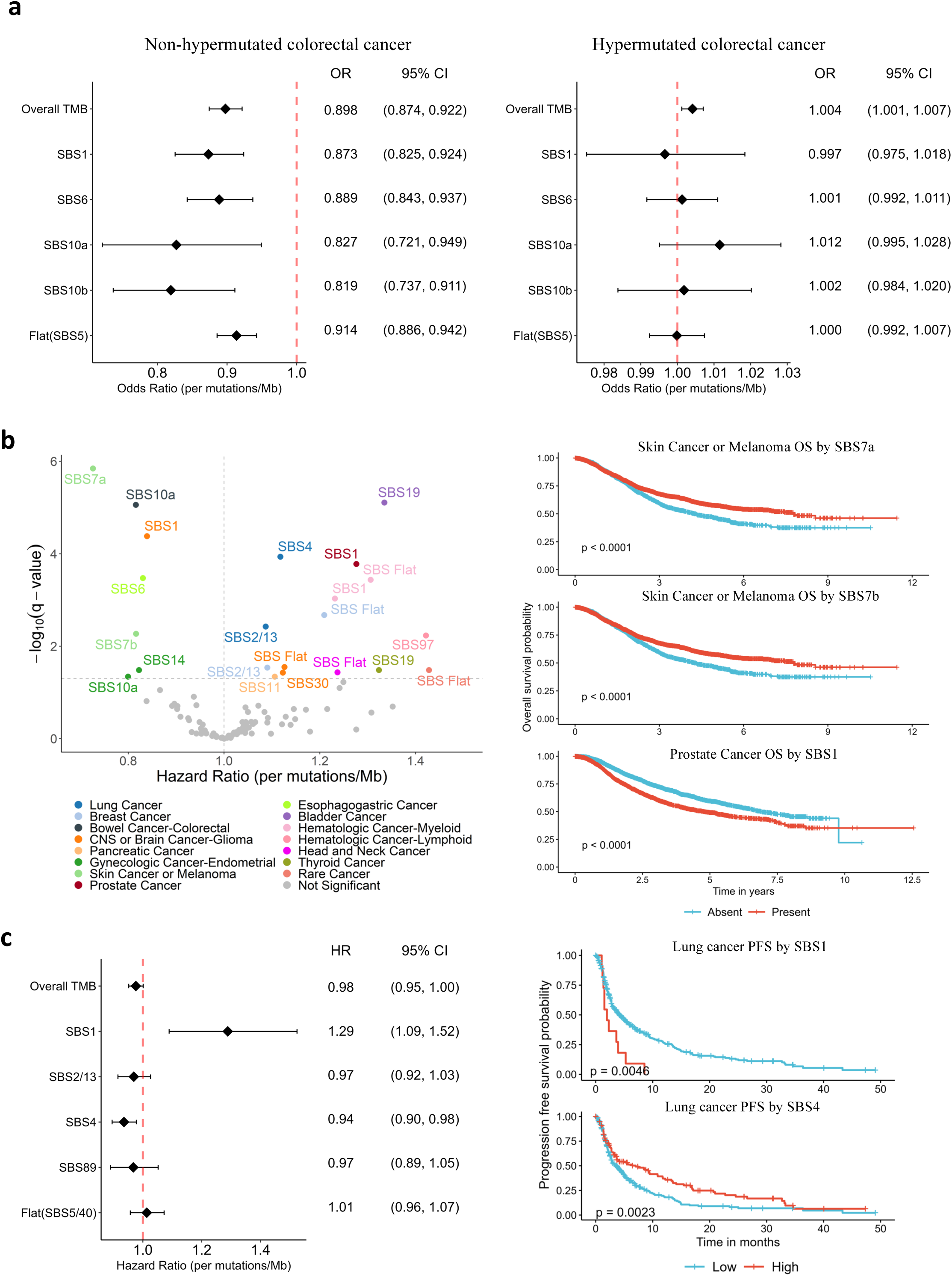
**Integration of targeted-sequencing-derived mutational signatures with clinical outcomes**. a, Early-onset versus late-onset colorectal cancer. Forest plots show adjusted odds ratios (ORs) for early-onset status associated with overall tumor mutational burden (TMB) and TMB attributed to SBS1, SBS5, SBS6, SBS10a and SBS10b. ORs are shown per mutation/Mb increase; error bars show 95% confidence intervals (CIs). Analyses included 9,562 colorectal cancer patients and were stratified into non-hypermutated tumors (TMB < 10 mutations/Mb; 2,495 early-onset and 6,009 late-onset cases; left) and hypermutated tumors (790 early-onset and 268 late-onset cases; right). Generalized linear mixed models adjusted for tumor status, self-reported race/ancestry, sex and selected signature-specific TMBs, with sequencing center and tumor subtype as random effects. b, Mutational signatures associated with overall survival. Left, volcano plot of hazard ratios (HRs) for overall survival versus -log10 false-discovery-rate (FDR)-adjusted q-values; HRs are shown per mutation/Mb increase in signature-attributed TMB, and associations with q < 0.1 are color-coded by cancer category. Survival time was measured in years from targeted sequencing to death or last known vital-status follow-up. Mixed-effects Cox models adjusted for tumor status, age, self-reported race/ancestry and sex where applicable, with sequencing center and tumor subtype as random effects. Right, Kaplan-Meier curves for overall survival probability over follow-up time, stratified by signature presence or absence for three representative associations; log-rank P values are shown. c, Progression-free survival (PFS) in immunotherapy-treated lung cancer. Left, mixed-effects Cox-model HRs with 95% CIs for overall TMB and signature-attributed TMBs for SBS1, SBS2/13, SBS4, SBS89 and flat SBS5/40 in 470 patients with non-small cell lung cancer who received immune checkpoint inhibitors. PFS was defined as time from immune checkpoint inhibitor initiation to radiologically or clinically confirmed progression or death. Models adjusted for age, sex, smoking history, stage IV disease and signature-specific TMBs, with sequencing center and tumor subtype as random effects. Top right, Kaplan-Meier curves for PFS stratified by high (>2 mutations/Mb) versus low (≤2 mutations/Mb) SBS1-attributed TMB; bottom right, analogous curves for SBS4-attributed TMB using the same cutoff. Log-rank P values are indicated.

#### Mutational signatures associated with overall survival across cancer types

We next evaluated associations between individual mutational signatures and overall survival across cancer types (Methods). Several signatures were associated with longer overall survival (Fig. 5b), including UV exposure signatures in skin cancer or melanoma (SBS7a, HR = 0.73, 95% CI = 0.64-0.82; SBS7b, HR = 0.82, 95% CI = 0.72-0.92). Other favorable associations involved defective POLE DNA replication or mismatch repair, including SBS10a in colorectal cancer (HR = 0.82, 95% CI = 0.75-0.89) and SBS6 in esophagogastric cancer (HR = 0.83, 95% CI = 0.76-0.91). Several signatures were also associated with worse prognosis, including smoking-related SBS4 in lung cancer (HR = 1.12, 95% CI = 1.06-1.18), consistent with clinical observations^46^, and APOBEC-associated SBS2/13 in lung cancer (HR = 1.09, 95% CI = 1.04-1.14) and breast cancer (HR = 1.09, 95% CI = 1.02-1.16). Clock-like or flat components showed cancer-type-dependent associations, including worse survival for SBS1 in prostate cancer (HR = 1.28, 95% CI = 1.14- 1.43) and flat profiles in myeloid-derived hematologic malignancies (HR = 1.31, 95% CI = 1.15- 1.49), but improved survival for SBS1 in glioma (HR = 0.84, 95% CI = 0.78-0.90). These findings indicate that targeted sequencing-derived SBS signatures can reveal cancer-type-specific survival associations after adjustment for clinical and sequencing variables.

#### Mutational signatures associated with immunotherapy response

Using linked treatment and response records, we examined whether signature-specific TMB was associated with progression-free survival (PFS) among 470 patients with non-small cell lung cancer who received immune checkpoint inhibitors. In multivariable models adjusted for clinical and demographic variables, SBS1-attributed TMB was associated with shorter PFS (HR = 1.29, 95% CI = 1.09-1.52, Fig. 5c left panel and top right panel), whereas SBS4-attributed TMB was associated with longer PFS (HR = 0.94, 95% CI = 0.90-0.98, Fig. 5c left panel and bottom right panel). TMB attributed to SBS2/13, SBS89 and SBS5/40 showed no significant association.

Overall TMB showed only a marginal association with PFS (HR = 0.98, 95% CI = 0.95-1.00, Supplementary Fig. 9a), and similar patterns were observed for overall survival (Supplementary Fig. 9b). These results indicate that partitioning TMB by mutational process can reveal outcome associations that are not captured by total TMB alone in immunotherapy-treated NSCLC.

## Discussion

In this study, we present SATS, a computational framework for mutational signature analysis in targeted sequencing data, and establish a real-world, panel-calibrated pan-cancer catalogue of SBS and DBS signatures from 111,711 tumors in AACR Project GENIE. The catalogue spans 102 OncoTree cancer types, organized into 23 cancer categories used for downstream analysis, and 757 cancer subtypes, enabling targeted sequencing-derived signatures to be evaluated across the breadth of tumors encountered in routine oncology practice. Using this resource, we characterized cancer-type and subtype-specific signature patterns and linked signature-attributed mutation burden to clinical phenotypes, including early-onset colorectal cancer, overall survival and immunotherapy outcome. Together, these findings show that targeted sequencing-derived signatures can be analyzed at scale in clinically annotated cohorts when panel design and mutation context are modeled explicitly.

Our study makes several practical contributions. First, SATS accounts for panel size and trinucleotide context, allowing mutational signatures to be analyzed across heterogeneous targeted sequencing panels rather than assuming uniform genomic coverage. Second, SATS estimates multiple co-occurring signatures in individual tumors, extending beyond clustering-based approaches that assign a dominant signature category. Third, this catalogue provides one of the largest pan-cancer mutational-signature resources for tumors profiled by targeted sequencing, including many cancer subtypes that were underrepresented in previous WES/WGS studies.

The real-world clinical setting is central to the value and interpretation of this resource. Unlike research-sequencing cohorts such as TCGA and PCAWG, AACR Project GENIE captures tumors tested across hospital and cancer-center laboratories as part of clinical care and links routine targeted sequencing to patient-level clinical information. This makes the catalogue particularly relevant to real-world oncology settings and enables analyses of prognosis and treatment response, while also reflecting variation in panel design and treatment history. SATS addresses the panel-opportunity component of this heterogeneity, and the catalogue provides a conservative, panel-calibrated reference set built around signatures repeatedly recovered across discovery and validation analyses. In this sense, the resource complements COSMIC and WES/WGS discovery catalogues rather than replacing them.

A further implication of this work is that targeted sequencing can move beyond total TMB toward mutational-process-specific burden estimates. Total TMB aggregates mutations generated by distinct biological and exposure-related processes, which may have different clinical associations. By partitioning TMB into panel-calibrated signature components, SATS enables analyses that distinguish smoking-related, POLE/MMR-related, APOBEC-associated and background mutational processes. This distinction was illustrated by the immunotherapy analysis, in which SBS4-attributed TMB showed a stronger association with outcome than total TMB in NSCLC. These findings support signature-specific TMB as a biologically interpretable extension of targeted-panel sequencing, while requiring further validation before prospective biomarker use.

This study also has limitations. SATS is not intended to replace WES/WGS for de novo mutational-process discovery when high-quality exome or genome data are available; those platforms remain preferable for discovering new signatures and resolving rare or weakly separated mutational processes. The intended use of SATS is large targeted-panel cohorts and retrospective clinical archives where WES/WGS is unavailable but panel-calibrated refitting and signature-specific TMB estimation are needed. Its performance depends on panel size, mutation burden and the number of signatures being evaluated. For small panels or low-TMB tumors, signature estimates should therefore be interpreted cautiously, particularly when several low-prevalence or similar signatures are included in the same refitting set.

Validation in GENIE version 17.0, OrigiMed, METABRIC and FUSCC/Fudan supports recovery of most panel-calibrated signatures across independent cohorts, including cohorts with different racial, ancestral and geographic compositions. However, these analyses do not constitute comprehensive population-level or platform-level validation. Lower-prevalence or less distinctive signatures, including SBS3, SBS5 and SBS40, remain challenging to separate in sparse targeted-panel data. Our simulations indicate that sufficiently large targeted-sequencing cohorts can improve separation of these signatures, suggesting that continued growth of real-world clinical sequencing datasets will increase the resolution of targeted-panel signature catalogues.

Future work should broaden validation across underrepresented populations, rare cancer subtypes and additional clinical sequencing platforms. Paired targeted sequencing, WES and WGS datasets will be particularly important for benchmarking panel-derived signatures against broader genomic profiles. Prospective studies are also needed to evaluate signature-specific TMB as a treatment-response biomarker and to define prespecified replication criteria for adding lower-prevalence or cohort-specific signatures to future versions of the catalogue.

In conclusion, SATS enables panel-calibrated mutational signature analysis in targeted sequencing data, and the accompanying real-world pan-cancer catalogue provides a clinically linked reference resource tailored to routine targeted-panel sequencing. Together, these resources support reproducible, clinically interpretable analysis of mutational signatures in oncology cohorts where WES/WGS is unavailable. SATS is freely available through GitHub and CRAN, with web-based tools for exploring the targeted-sequencing signature catalogue and performing online signature refitting.

## Methods

Additional methodological details, including data filtering, panel-context construction, model derivations, validation analyses, simulations, comparator analyses and clinical-outcome models, are provided in the Supplementary Methods.

### AACR Project GENIE genomic data

We retrieved the AACR Project GENIE dataset (version 13.0-public) from Synapse (https://synapse.org/genie). This dataset included tumors profiled by targeted sequencing panels as part of routine clinical care across 16 hospitals or cancer centers in the United States, Canada, and Europe. When multiple tumors were sequenced from the same patient, one tumor was selected at random for analysis. Tumors sequenced with gene panels covering >50 kb of genomic region were included. Patient consent and institutional review board approvals were obtained according to the policies of participating institutions and AACR Project GENIE. The cohort represents a demographically diverse patient population, with self-reported race/ancestry including 5,973 Asian individuals (5.3%), 5,545 Black individuals (5.0%), 78,003 White individuals (69.8%), 5,311 individuals from other groups (4.8%), and 16,879 individuals with unknown race/ancestry (15.1%). Self-reported sex included 58,576 females, 47,694 males, 2 individuals reported as other, and 5,439 individuals with unknown sex.

Sequencing was conducted at CLIA- and/or ISO-certified laboratories with high read depth (median read depth, 519x; interquartile range, 307x-808x). Somatic mutations were called by participating centers using tools such as Mutect2 (ref^47^) and Strelka^48^. Germline variants and artefacts were filtered using pooled external controls and databases of known germline variants, including the Genome Aggregation Database (gnomAD)^49^. Additional details on the GENIE filtering procedure are provided in the AACR GENIE 13.0-public Data Guide (https://www.aacr.org/wp-content/uploads/2023/03/13.0_data_guide-1.pdf). We applied additional uniform filters, retaining somatic mutations with read depth of at least 100 and alternate allele count of at least 5. After filtering, 982,095 single base substitutions (SBSs) and 15,149 double base substitutions (DBSs) from 111,711 tumors were retained for downstream mutational signature analysis.

Because AACR Project GENIE is a real-world consortium dataset, variant calling and filtering were performed by the contributing institutions rather than by a single centralized pipeline. To reduce technical variability before signature analysis, we used GENIE-filtered somatic mutations and applied additional uniform read-depth and allele-count filters. Users applying SATS to other targeted sequencing cohorts should review cohort-specific mutation calling and filtering procedures and follow best practices for somatic mutation calling and filtering^50,51^.

### Cancer categories of AACR Project GENIE

AACR Project GENIE included 102 cancer types (Supplementary Table 1) and 757 cancer subtypes defined by OncoTree^24^. To facilitate cancer type-specific analysis, we consolidated the 102 OncoTree cancer types into 23 cancer categories used for downstream analysis. For example, "breast cancer", "breast cancer, NOS" and "Breast Sarcoma" were grouped into a single breast cancer category (Supplementary Table 2). Within each cancer category, subtypes representing less than 3% of tumors were aggregated into a "rare" category.

### Calculation of panel-specific mutation opportunities

For each targeted sequencing panel in AACR Project GENIE (ranging in size from a few genes to several hundred genes; Supplementary Table 3), we quantified the number of genomic loci at which each SBS or DBS mutation type could be observed (Supplementary Tables 4 and 5).

Genomic coordinates for each panel were obtained from Synapse (https://www.synapse.org/#!Synapse:syn26706790), and the corresponding DNA sequences were extracted from the human reference genome (hg19). For SBSs, we considered the standard 96 substitution types, defined by six classes of base substitution (C>A, C>G, C>T, T>A, T>C and T>G) across 16 trinucleotide contexts. These are encoded as mutations of a central pyrimidine base within its immediate 5’ and 3’ flanking nucleotides, yielding 4 x 6 x 4 = 96 distinct SBS types and 4 x 2 x 4 = 32 trinucleotide mutation contexts. For DBSs, we considered 78 mutation types from 10 dinucleotide mutation contexts (AC, AT, CC, CG, CT, GC, TA, TC, TG and TT). We then enumerated all instances of each mutation context within each panel to build panel-specific context matrices for subsequent SATS modeling.

#### Panel-calibrated Poisson NMF model for signature analysis of tumor mutational burden

SATS models targeted-sequencing mutation counts with a panel-calibrated Poisson non-negative matrix factorization (pNMF). For each tumor and mutation type, the model represents the observed SBS count as a Poisson random variable whose expected value depends on three components: the panel-specific mutation opportunity, the mutational-signature profile and the tumor-level signature activity:

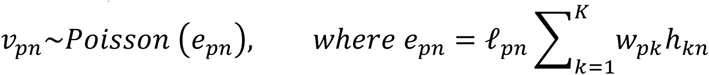

Here, the panel context term records the number of genomic contexts in which a mutation type could have been observed for the panel used to sequence each tumor. The signature-profile matrix represents the relative contribution of each mutation type within each signature, and the activity matrix represents the burden of each signature in each tumor. This formulation is equivalent to the pNMF model used in signeR^4^, with explicit adjustment for panel-specific mutation opportunity.

The signature-profile and activity matrices were estimated by maximizing the pNMF log- likelihood:

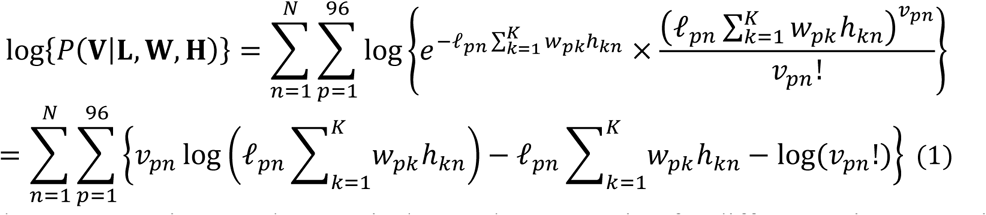

The panel context matrix extends canonical NMF by accounting for differences in sequencing coverage and trinucleotide opportunity across targeted panels. When all tumors are sequenced over the same genomic region, the opportunity term is constant across samples and the model reduces to canonical NMF. The derivation of this special case is provided in the Supplementary Methods.

### Extraction of de novo TMB signatures

We used signeR^4^ to extract de novo mutational signature profiles under the pNMF model with the panel-specific opportunity matrix. Because signeR uses a computationally intensive Markov chain Monte Carlo procedure^4^, tumors were grouped into sets of 100 within each cancer type before extraction. For SBS signatures, candidate ranks 1-5 were evaluated for each cancer type, except CNS/brain cancer-glioma, for which ranks 1-6 were evaluated; for DBS signatures, ranks 1-3 were evaluated. The selected rank was taken from the signeR model-selection output, which includes the BIC profiles and selected number of signatures. A fixed random seed of 4022022 was used for extraction. The grouped-sample derivation is provided in the Supplementary Methods.

Sample grouping substantially improved computational efficiency. Applying SATS to 10,000 tumors grouped into sets of 100 reduced runtime to 28.5 minutes on a standard laptop (Intel Core i7-1165G7, 2.80 GHz, 16 GB RAM), compared with approximately 13 hours without grouping.

### Mapping de novo TMB signatures to COSMIC reference TMB signatures

Targeted sequencing panels often contain too few somatic mutations to resolve every de novo profile as a single known process. Consequently, detected de novo TMB signatures may represent mixtures of related COSMIC reference TMB signatures. SATS therefore maps the de novo signature-profile matrix to a reference TMB signature matrix, such as the COSMIC SBS reference TMB matrix containing 85 signatures, using penalized non-negative least squares (pNNLS)^26^:

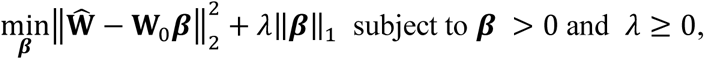

where the coefficient matrix represents the contribution of each reference signature to each de novo profile, and the tuning parameter is selected by cross-validation. Compared with standard NNLS,

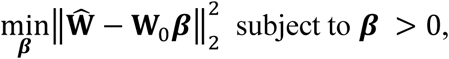

The pNNLS objective introduces an L1-norm penalty on the coefficient matrix, promoting sparsity by shrinking small coefficients toward zero. This favors a smaller, more interpretable subset of reference signatures for downstream refitting.

To reduce instability from cross-validation and finite mutation counts, we repeated the mapping procedure 100 times and retained only reference TMB signatures with coefficients >0.1 in at least 80 of the 100 repetitions. This stability criterion produced a robust subset of COSMIC signatures with recurrent contributions to the de novo TMB profiles.

### Estimation of signature activities using an expectation-maximization algorithm

After mapping, SATS refits the selected reference TMB signatures to individual tumors using an expectation-maximization (EM) algorithm. In this refitting step, the reference TMB signatures are fixed, and the tumor-level signature activities are estimated from the observed mutation-count matrix and the panel context matrix. The algorithm treats the unobserved signature-attributed mutation counts as latent variables and alternates between allocating observed mutation counts across signatures and updating the activity of each signature. The detailed EM derivation and update equations are provided in the Supplementary Methods.

The E-step and M-step were iterated until convergence to produce the estimated activity matrix. In the implementation used for this study, each refitting run used 50 random starts, a maximum of 5,000 iterations per start and a convergence tolerance of 1x10^-5^, defined by the relative change in normalized Poisson log-likelihood between successive iterations. The solution with the highest final log-likelihood across starts was retained for downstream burden estimation. Because the activity update for each tumor depends only on that tumor’s mutation counts and panel opportunity vector, SATS can estimate signature activities for a single tumor or a small subset of tumors without requiring joint modeling across the full cohort.

### Calculation of signature burdens

Signature burden was defined as the expected number of mutations attributed to a given signature in a given tumor. After EM convergence, SATS combines the estimated activity, the fixed reference TMB signature profile and the panel-specific opportunity vector to calculate the signature burden:

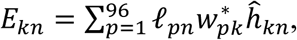

where the burden is summed across the 96 SBS mutation types for the tumor-specific panel opportunity, reference signature profile and estimated activity. This quantity partitions the observed panel mutation burden into signature-attributed burdens while accounting for panel size and context composition.

### Relationship between TMB-normalized and TMC signatures

COSMIC reference signatures are commonly represented as tumor mutational count (TMC) profiles, whereas SATS uses TMB-normalized profiles that account for mutation opportunity. A TMB-normalized signature profile can be obtained from a TMC signature profile by dividing each mutation-type contribution by the number of genomic contexts in which that mutation type can occur and then renormalizing the profile:

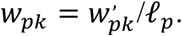

This conversion allows WGS-derived COSMIC TMC signatures^1^ to be used as panel-calibrated TMB reference signatures for SATS mapping and refitting.

### Construction of COSMIC TMB-normalized reference signatures

To generate the reference TMB signatures used in SATS, we downloaded COSMIC SBS and DBS signature profiles (version 3.4) from the COSMIC database (https://cancer.sanger.ac.uk/signatures/). For each SBS signature, the contribution of each mutation type was divided by the number of corresponding trinucleotide contexts in the human reference genome and then rescaled so that the 96 SBS mutation types summed to one for each signature (Supplementary Tables 4 and 6). The same procedure was applied to COSMIC DBS signatures using the corresponding dinucleotide context counts to generate reference DBS TMB signatures (Supplementary Tables 5 and 7).

We constructed a pan-cancer catalogue of mutational signatures from targeted sequencing data in AACR Project GENIE version 13.0-public. The SATS pipeline was applied separately to each cancer category used for downstream analysis, so that signatures were estimated within panel and tumor-type contexts rather than from a pooled pan-cancer count matrix. To improve computational efficiency while preserving cancer-specific signal, tumors were grouped into sets of 100 for signature extraction.

For reproducibility and validation analyses, we used GENIE version 17.0 (downloaded from Synapse: https://www.synapse.org/Synapse:syn55234548) and selected samples that were not present in GENIE version 13.0-public. The internal GENIE held-out analysis focused on lung, colorectal and breast cancers, which had sufficiently large sample sizes for replication. External validation used an independent lung and colorectal cancer cohort from the OrigiMed study in cBioPortal (https://www.cbioportal.org/study/summary?id=pan_origimed_2020), together with breast cancer datasets from METABRIC (https://www.cbioportal.org/study/summary?id=brca_metabric) and FUSCC/Fudan (https://data.3steps.cn/cdataportal/study/clinicalData?id=FUSCC_BRCA_panel_4000).

The same analytical pipeline was applied to the validation cohorts. Because the validation cohorts were smaller than the discovery set, tumors were grouped into sets of 50 and each analysis was repeated 10 times using different random seeds. Hypermutated tumors were excluded from the OrigiMed and FUSCC/Fudan cohorts, and mutations consistent with 8-oxo- guanine sequencing artefacts were removed from the FUSCC/Fudan dataset before signature analysis. The exclusion and artefact-filtering criteria are described in the Supplementary Methods.

### Visualization of SBS mutational signature profiles using UMAP

To visualize SBS mutational signature profiles, we applied UMAP^52^ (Uniform Manifold Approximation and Projection) to signature burden profiles. Within each cancer category, tumors were first clustered by signature burden using hierarchical clustering with Ward’s minimum variance method, and clusters were defined using a dissimilarity threshold of 0.05. UMAP was then applied to the median signature burden profile of each cluster; therefore, each point in the two-dimensional map represents a group of tumors with similar signature composition rather than a single tumor.

### Analysis of subtype-specific heterogeneity of mutational signatures

To evaluate subtype-level heterogeneity in mutational signature presence, we fitted generalized linear mixed models (GLMMs) within cancer categories. Signature presence was modeled as a binary outcome, with age, self-reported race/ancestry, sex where applicable and tumor status (primary versus metastatic) included as fixed effects. Tumor subtype and sequencing center were modeled as random intercepts to account for subtype-specific enrichment and institutional differences in panel design or sequencing practice. Analyses used tumors with available values for the variables included in each model.

The presence of a signature was defined as a binary variable indicating whether the signature accounted for more than 1% of SBSs observed in the targeted sequencing panel. Only tumors sequenced using panels larger than 1 Mb were included. Panel size, measured in megabases, was included as an offset term as specified in the model below to account for the higher probability of detecting a signature in larger panels.

The GLMM was specified as:

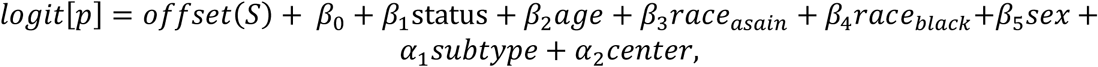

where the left-hand side represents the log-odds of signature presence. Tumor status (reference: primary), self-reported race/ancestry (reference: White), sex (reference: female) and age were modeled as fixed effects. Tumor subtype and sequencing center were included as random effects, with random intercepts modeled as:

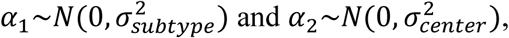

where the random effects were assumed to follow normal distributions with mean zero and their corresponding variance parameters. Sex was excluded as a covariate in sex-specific cancer categories, including breast, ovarian, endometrial, other gynecologic cancers and prostate cancer. GLMMs were fitted using the ’lme4’ package in R, and multiple comparisons across signatures and cancer categories were adjusted using the Benjamini-Hochberg false-discovery-rate procedure^53^.

Association between mutational signatures and early-onset colorectal cancer status To evaluate associations between mutational signatures and early-onset colorectal cancer status, defined as sequencing age <50 years, we analyzed SBS mutational signature profiles and clinical data from 9,562 colorectal cancer patients. Patients were stratified into non-hypermutated tumors (TMB < 10 mutations/Mb; early-onset: 2,495 cases; late-onset: 6,009 cases) and hypermutated tumors (early-onset: 790 cases; late-onset: 268 cases).

Within each TMB stratum, we used GLMMs to compare early-onset and late-onset cases. Tumor status (primary versus metastatic), self-reported race/ancestry, sex and signature-specific TMBs for SBS1, SBS6, SBS10a, SBS10b and flat SBS were included as fixed effects. Sequencing center and tumor subtype were included as random effects, and models were fitted using the ’lme4’ package in R.

### Prognostic analysis of mutational signatures

To evaluate associations between SBS mutational signatures and overall survival, we fitted mixed-effects Cox proportional hazards models within each cancer category. The hazard function was defined as:

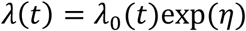

where time was measured in years from targeted sequencing to death or last follow-up with known vital status. Death was treated as the event, and living patients were censored at last follow-up. The second term denotes the baseline hazard function.

The linear predictor *η* was modeled as:

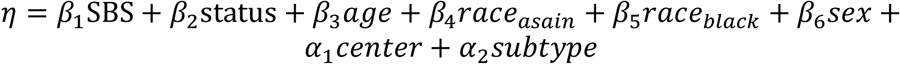

The fixed effects included the presence or absence of each SBS signature, tumor status (primary versus metastatic), age, self-reported race/ancestry and sex where applicable. Sequencing center and tumor subtype were included as random effects, with random-effect terms assumed to follow normal distributions as specified above. Analyses used patients with available values for the variables included in each model.

Mixed-effects Cox proportional hazards models were fitted using the ’coxme’ package in R. Sex was excluded as a covariate in models for breast, ovarian, endometrial, other gynecologic cancers and prostate cancer.

### Association between mutational signatures and immunotherapy outcomes

The AACR Project GENIE Biopharma Collaborative released detailed clinical data for 1,846 patients with non-small cell lung cancer (NSCLC; v2.0-public) treated at four institutions: MSKCC, DFCI, VICC and UHN. From this dataset, we analyzed 470 patients who received immune checkpoint inhibitor therapy, including pembrolizumab, nivolumab, atezolizumab or durvalumab. We evaluated whether mutational signature-specific TMBs were associated with immunotherapy outcomes.

We fitted mixed-effects Cox proportional hazards models to assess associations between mutational signature-specific TMB and progression-free survival (PFS). PFS was defined as the time from ICI initiation to radiologically or clinically confirmed disease progression or death. Age, sex, smoking history, tumor stage (stage IV versus others) and signature-specific TMBs for SBS1, SBS2/13, SBS4, SBS89 and flat SBS were modeled as fixed effects. Sequencing center and tumor subtype were modeled as random effects. The same modeling framework was applied to overall survival (OS), defined as the time from ICI initiation to death or last follow-up. Analyses used patients with available values for the variables included in each model.

### Software

The SATS R package source code (version v1.0.10), source archive, manual, user guide, example data, regression tests, utilities for generating mutation-count and panel-context matrices, lightweight converters for simple single-sample Variant Call Format (VCF) and Browser Extensible Data (BED) input preparation, input-validation functions, Docker assets and Nextflow-compatible examples are available at https://github.com/binzhulab/SATS. The repository provides the versioned package and reproducibility resources used for this study. The SATS R package is also available from CRAN at https://CRAN.R-project.org/package=SATS.

Interactive catalogue plots displaying the frequencies and etiologies of SBS and DBS signatures are available at https://analysistools.cancer.gov/mutational-signatures/#/catalog/STS. An online signature refitting tool is accessible at https://analysistools.cancer.gov/mutational-signatures/#/refitting.

## Data Availability

The AACR Project GENIE dataset could be retrieved from Synapse (https://synapse.org/genie)

https://github.com/binzhulab/SATS

## Author contributions

SJC, JS, MTL, and BZ contributed to the study concept and design and obtained the funding. DL, MH, DW, LS, TZ, XH and BZ conducted bioinformatic and biostatistical analyses. KY, XRY, SJC, JS, MTL, and BZ performed data interpretation. DL, MH and BZ drafted the initial manuscript. All authors contributed to the critical revision of the manuscript and approved the final manuscript.

## Acknowledgments

This research was supported by the Intramural Research Program of the National Institutes of Health (NIH). The contributions of the NIH authors were made as part of their official duties as NIH federal employees, are in compliance with agency policy requirements, and are considered Works of the United States Government. However, the findings and conclusions presented in this paper are those of the authors and do not necessarily reflect the views of the NIH or the U.S. Department of Health and Human Services. Additionally, Dr. Lee was supported by the National Research Foundation of Korea (NRF) grant funded by the Korea government (MSIT, No. RS- 2023-00213625). The authors acknowledge the American Association for Cancer Research and its financial and material support in the development of the AACR Project GENIE registry, as well as members of the consortium for their commitment to data sharing. Interpretations are the responsibility of study authors. This study used the high-performance computational capabilities of the Biowulf Linux cluster at the National Institutes of Health, Bethesda, MD: https://biowulf.nih.gov. We thank Bill Wheeler (Information Management Services) for computation support.

---

**Supplementary Fig 1.**
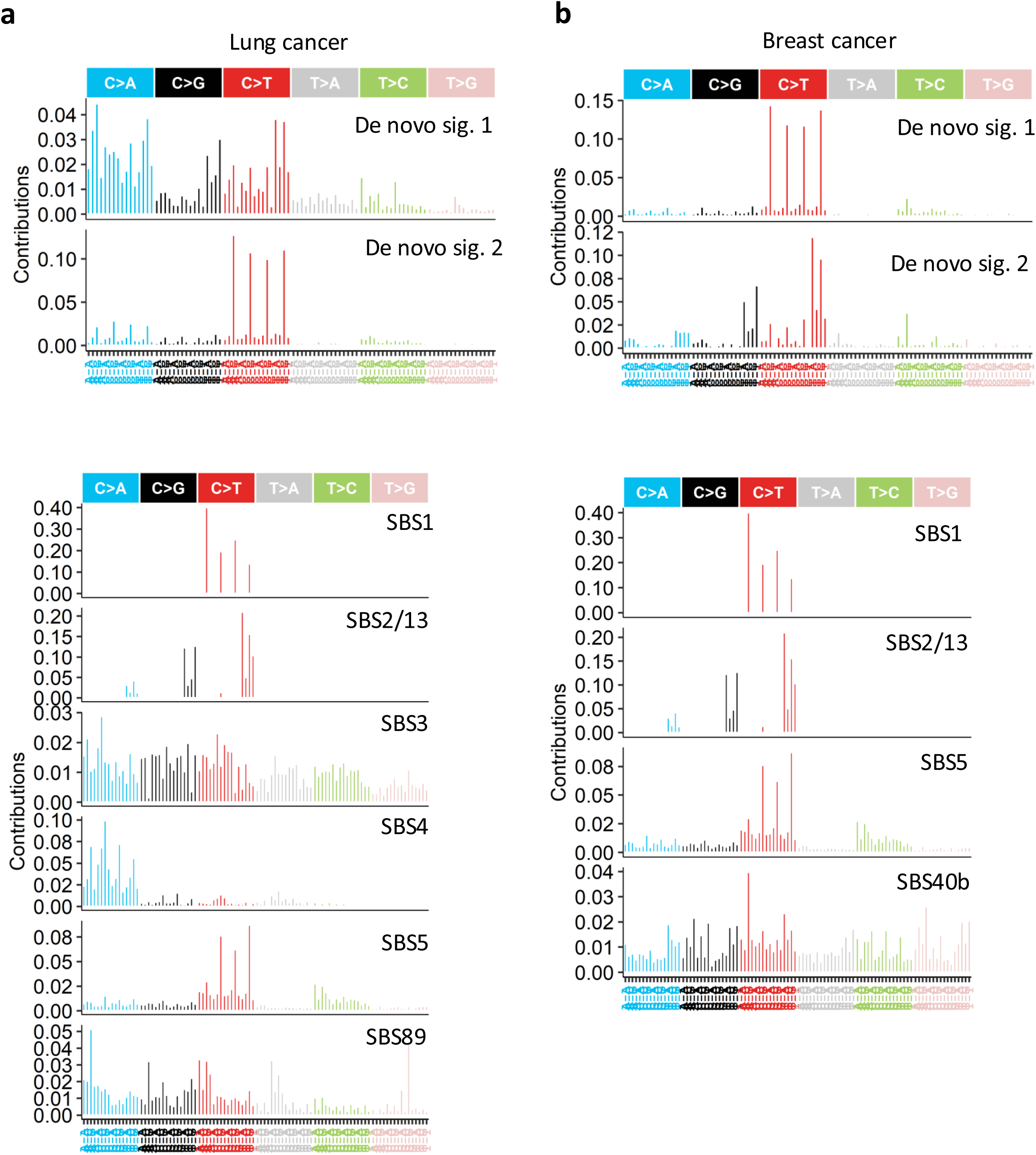
De novo and COSMIC-mapped targeted-sequencing signatures. a,b, De novo SATS tumor mutational burden (TMB) signature profiles across 96 SBS mutation types and recurrent COSMIC v3.4 TMB signature mappings for lung cancer (a) and breast cancer (b). De novo profiles were extracted from targeted-sequencing data and mapped to COSMIC reference signatures by recurrent penalized non-negative least squares. Labels above mapped profiles indicate the corresponding COSMIC SBS signatures or composite signature groups.

**Supplementary Fig 2.**
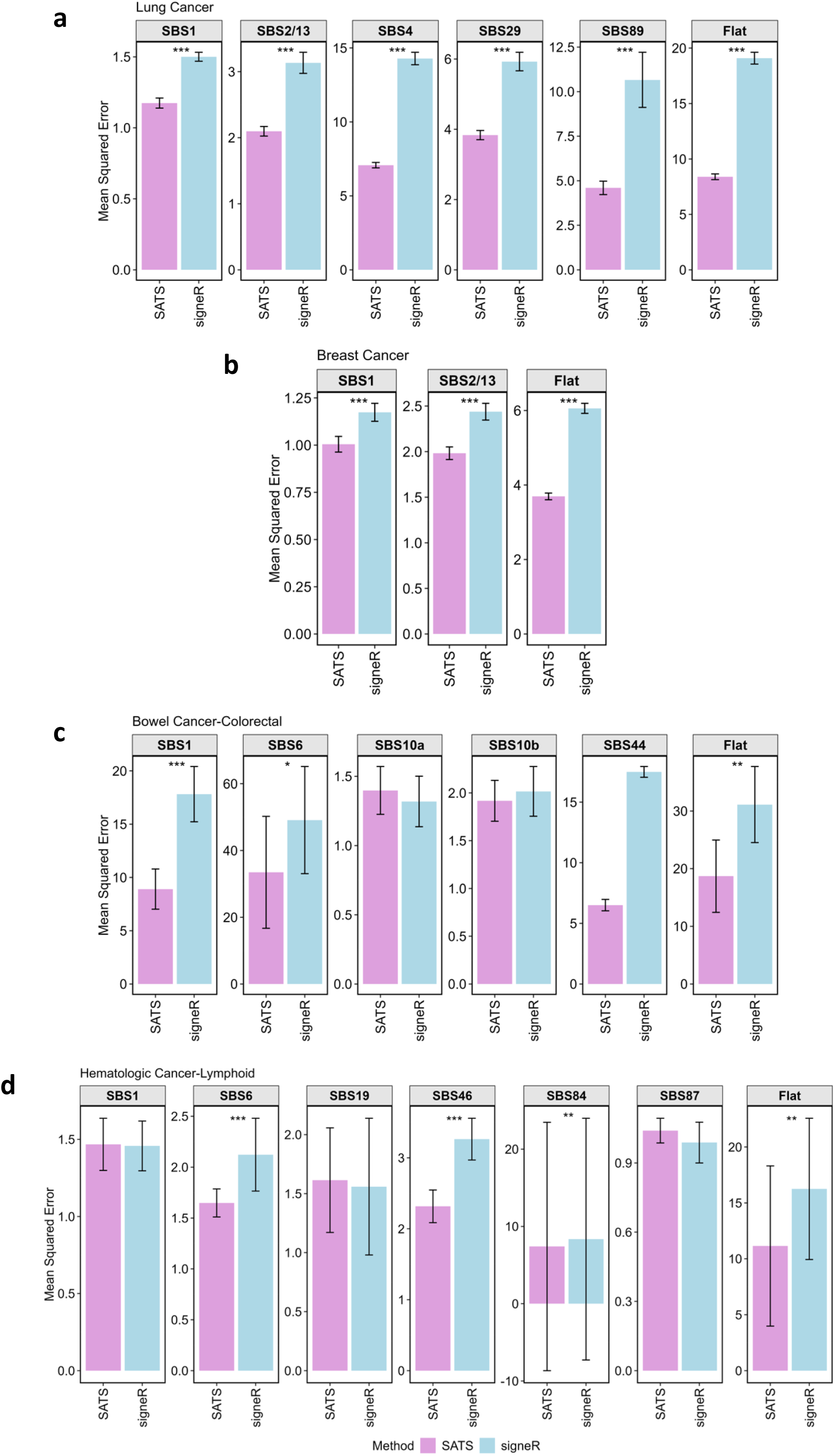
SATS refitting accuracy compared with signeR. a-d, Mean squared error (MSE) of signature-burden estimates from SATS and signeR in ten simulated targeted-sequencing replicates for lung cancer (a), breast cancer (b), colorectal cancer (c) and lymphoid-derived hematologic cancer (d). Bars show mean MSE across replicates; error bars show s.d. Asterisks indicate two-sided Wilcoxon rank-sum tests comparing MSE distributions between methods (*P < 0.05, **P < 0.01 and ***P < 0.001).

**Supplementary Fig 3.**
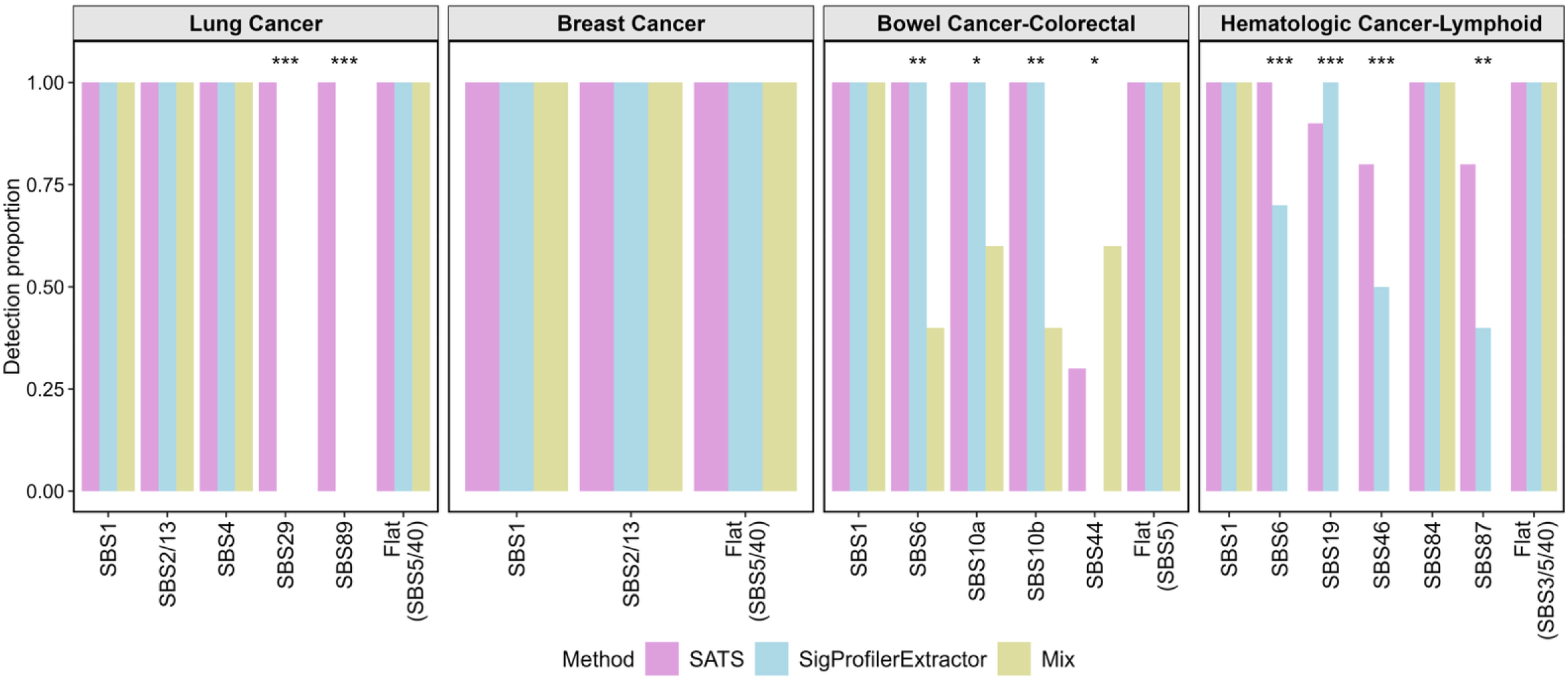
**Active-signature detection benchmark**Bars show the proportion of ten simulation replicates in which each ground-truth signature was detected in lung, breast, colorectal and lymphoid-derived hematologic cancer simulations. Mutation counts were generated by Poisson sampling from prespecified signature mixtures with cancer-type sample sizes matched to AACR Project GENIE cohorts. Asterisks indicate Kruskal-Wallis rank-sum tests comparing detection proportions across methods for each signature (***P < 0.001, **P < 0.01 and *P < 0.05).

**Supplementary Fig 4.**
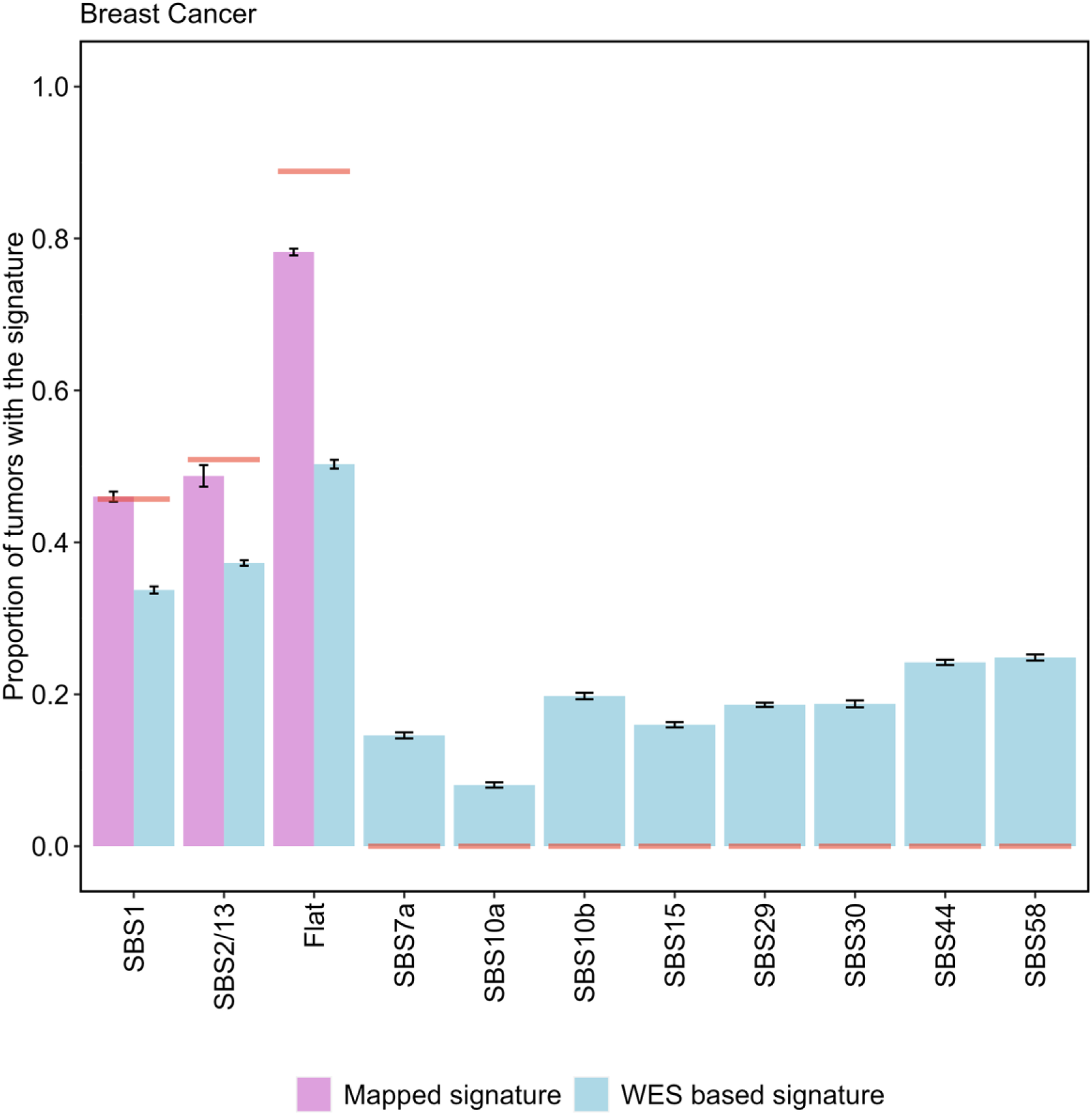
False attribution from broad WES-derived reference sets. Breast cancer targeted-sequencing data were simulated from SBS1, SBS2/13 and SBS5 and refitted using either the matched mapped signature set or a broader TCGA WES-derived breast cancer set. The WES-derived set includes SBS1, SBS2/13, SBS5, SBS3, SBS7a, SBS10a, SBS10b, SBS15, SBS29, SBS30, SBS44 and SBS58, each contributing >1% of SBSs in TCGA WES breast cancer; all except SBS1, SBS2/13 and SBS5 are absent from the simulated ground truth. Horizontal lines show true simulated proportions; error bars show mean ± s.d.

**Supplementary Fig 5.**
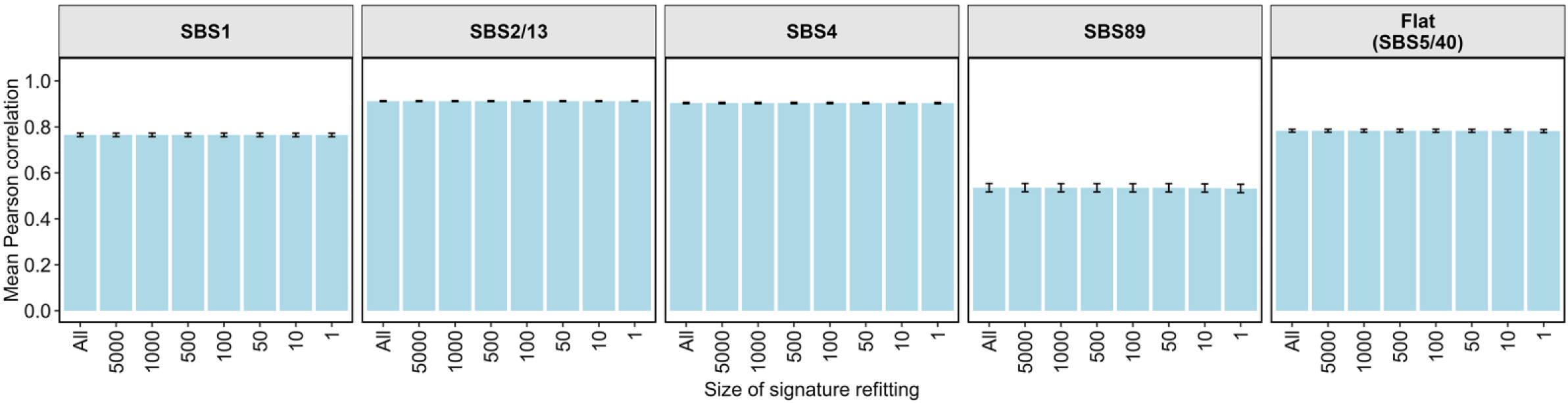
SATS refitting accuracy across lung cancer sample sizes. Bars show mean Pearson correlations between simulated and SATS-estimated SBS signature burdens across ten lung-cancer simulation replicates. Refitting was performed with 100-sample, 10-sample and single-sample subsets drawn from the same simulated targeted-sequencing datasets. Error bars show s.d.

**Supplementary Fig 6.**
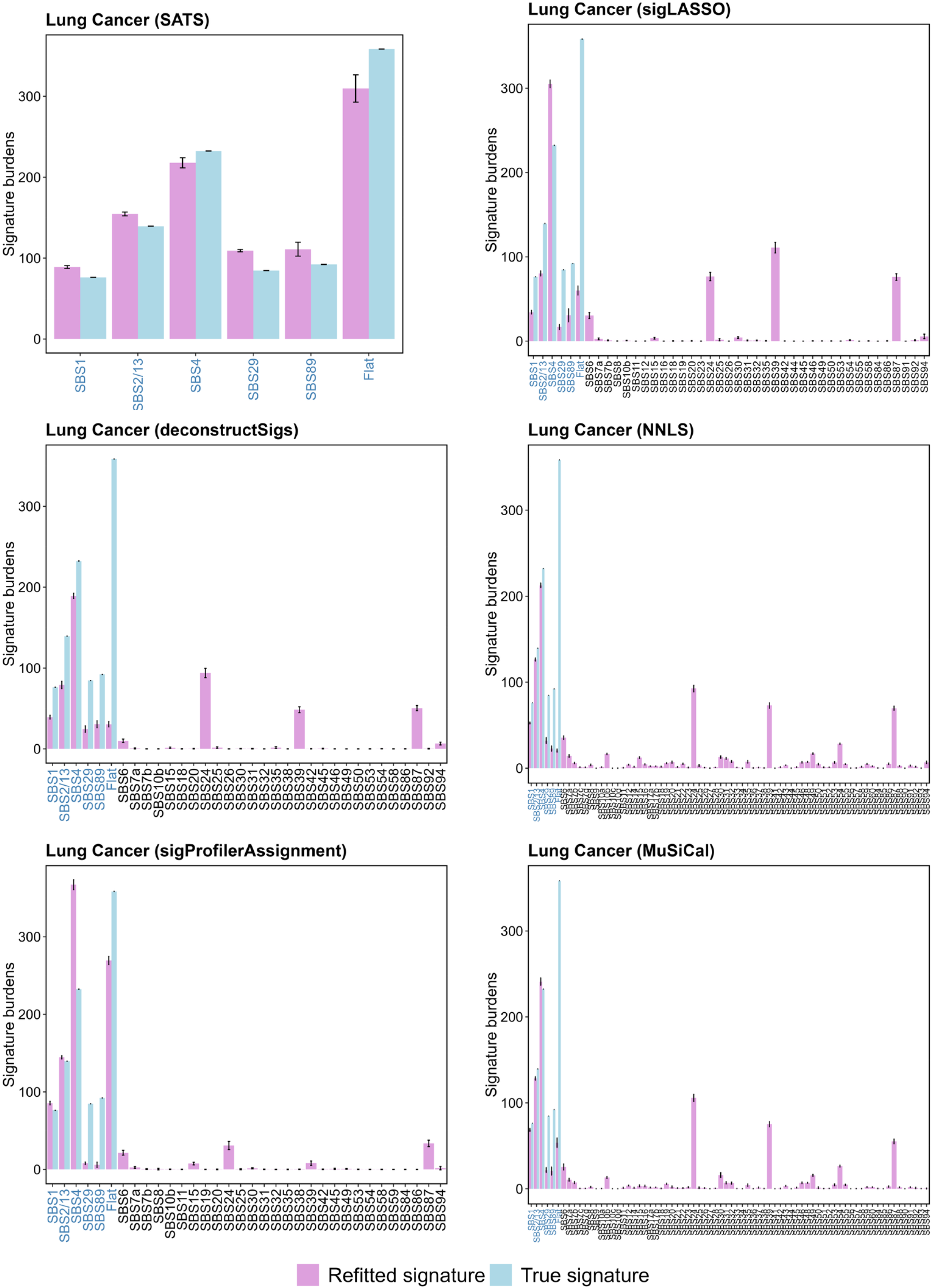
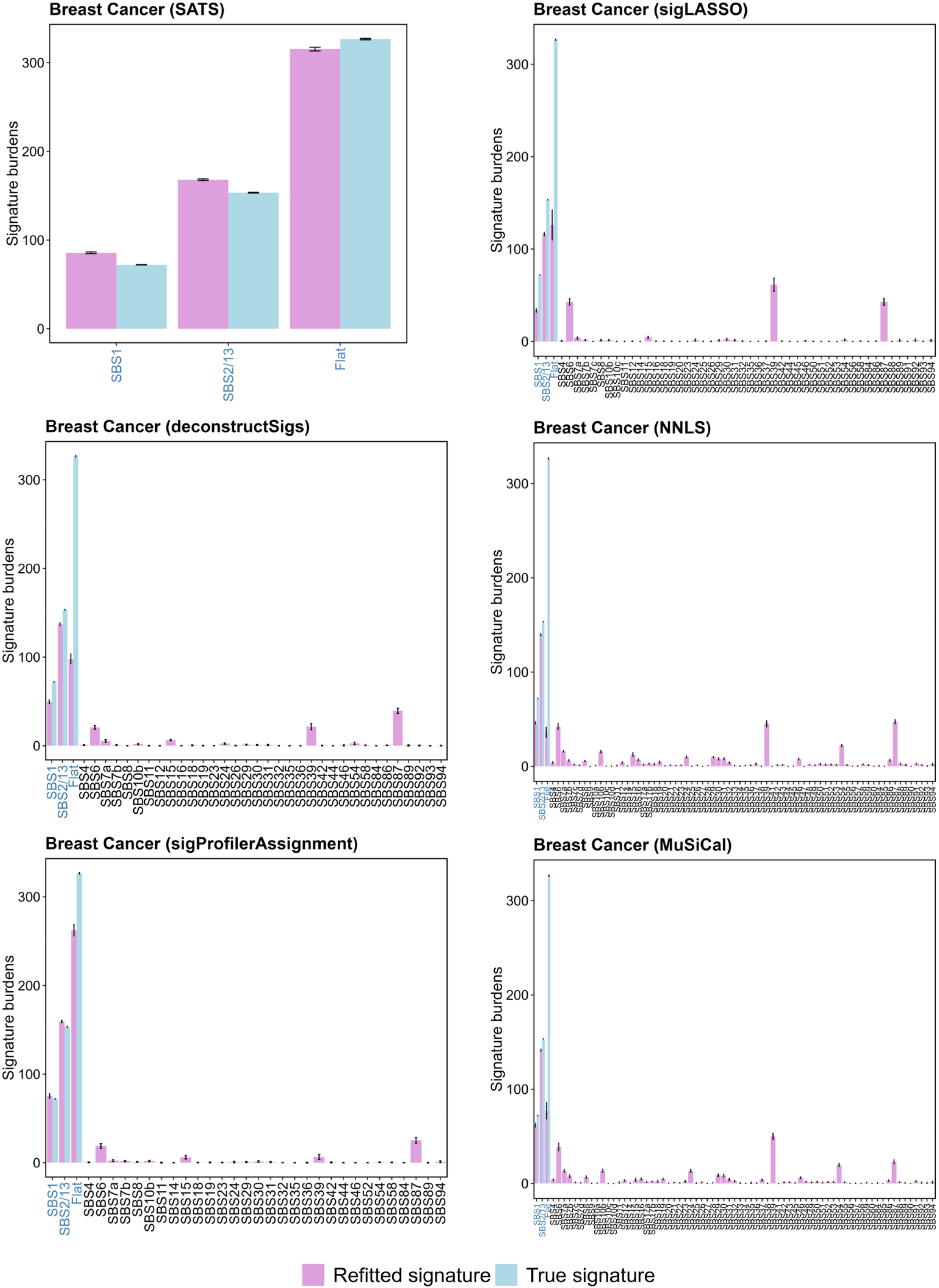
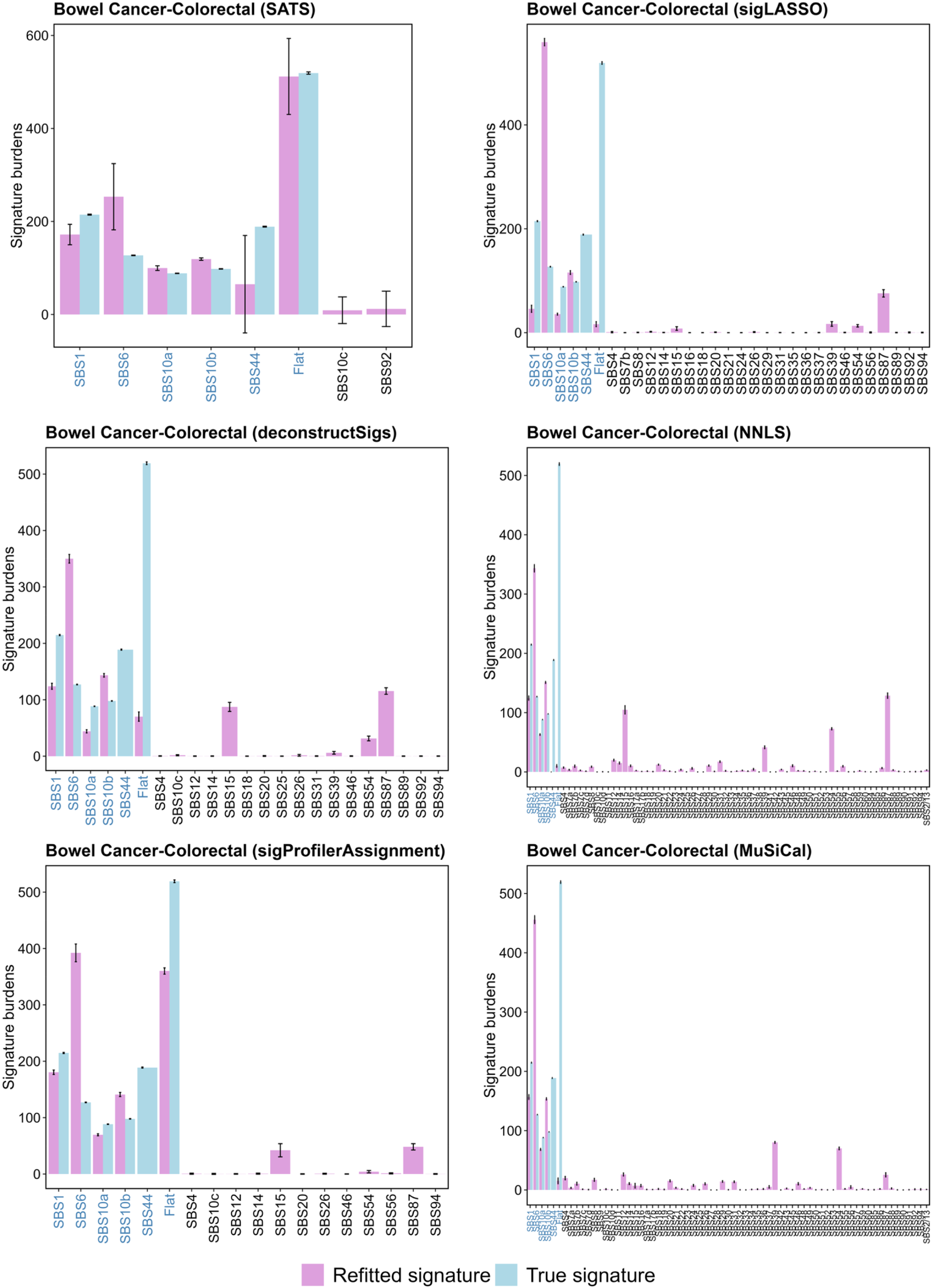
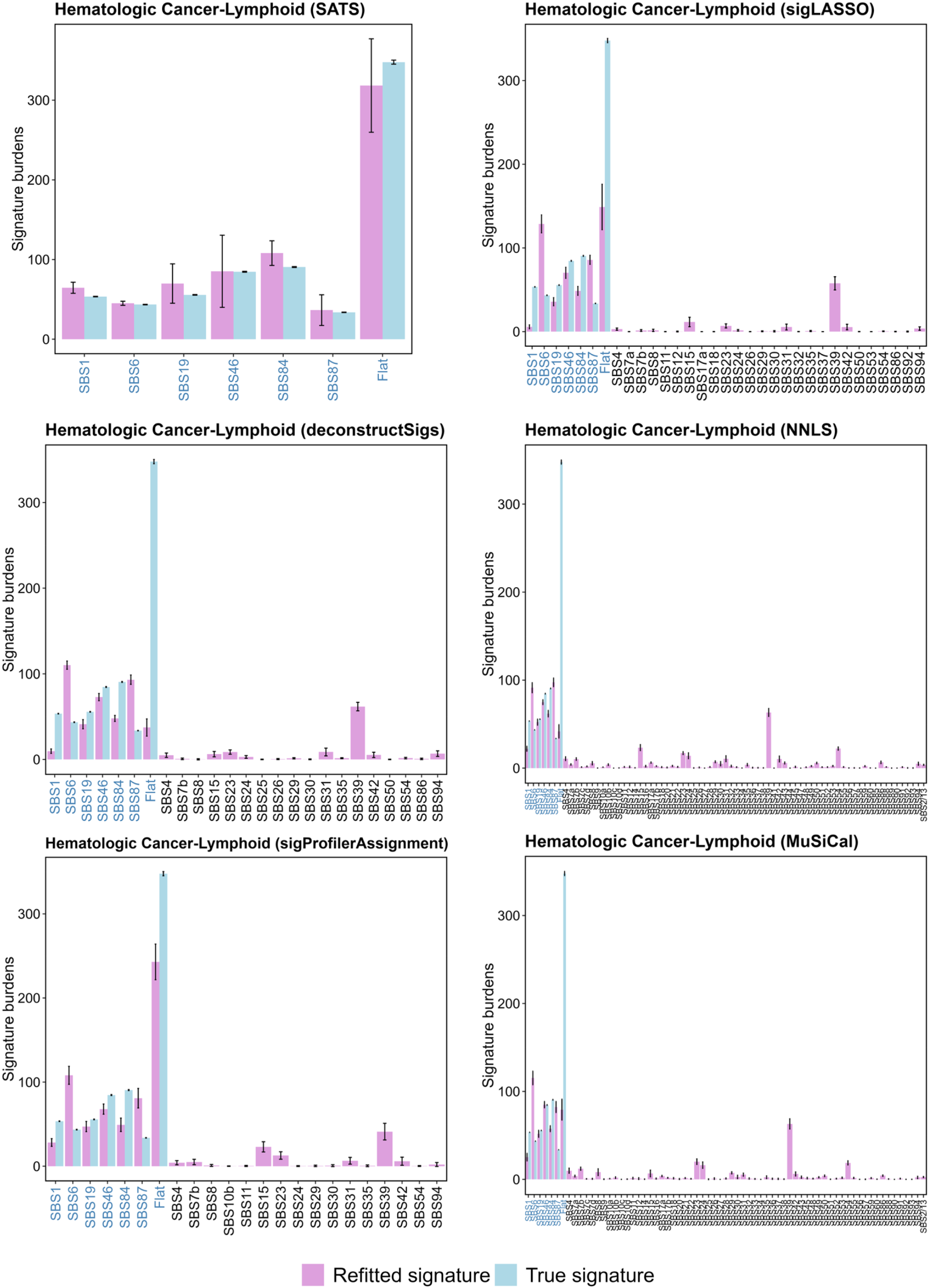
Broad-catalogue refitting stress test. Signature-burden estimates from SATS, sigLASSO, deconstructSigs, NNLS, SigProfilerAssignment and MuSiCal are shown for simulated lung, breast, colorectal and lymphoid-derived hematologic cancer data. Each cancer type is represented by one representative 100-tumor composite from the same simulation framework. Refitting-only methods are supplied with the full tumor mutational count (TMC) signature catalogue because these tools require prespecified reference signatures. Pink bars show refitted signature burdens and blue bars show simulated true burdens. Non-ground-truth assignments represent false or off-target attribution. This analysis is an exploratory specificity stress test of broad-catalogue refitting, not a formal like-for-like benchmark of complete workflows.

**Supplementary Fig 7.**
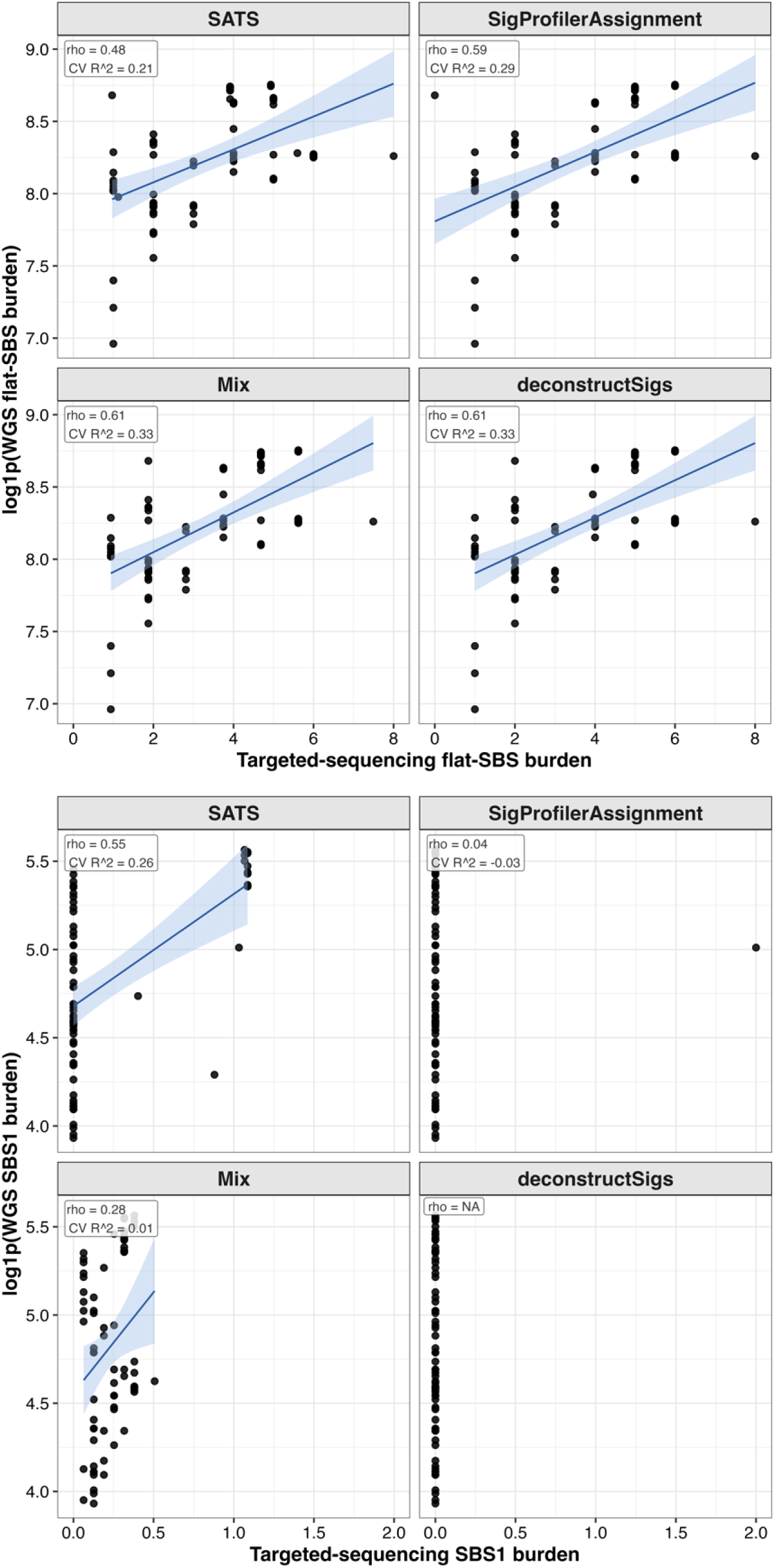
**Matched WGS and targeted-sequencing benchmark**Targeted-sequencing-derived burdens estimated by SATS, SigProfilerAssignment, Mix and deconstructSigs are plotted against log1p-transformed matched WGS burdens for the flat SBS5/SBS40 category and SBS1 in 72 kidney tumors profiled by both WGS and a custom 254-gene, 1.90-Mb targeted panel. Each point represents one tumor. Blue lines show linear fits with 95% CIs. Inset statistics show Spearman rho and leave-one-out cross-validated R2 for predicting log1p(WGS burden) from targeted-sequencing burden. Correlation was not estimable when a method assigned constant zero burden.

**Supplementary Fig 8.**
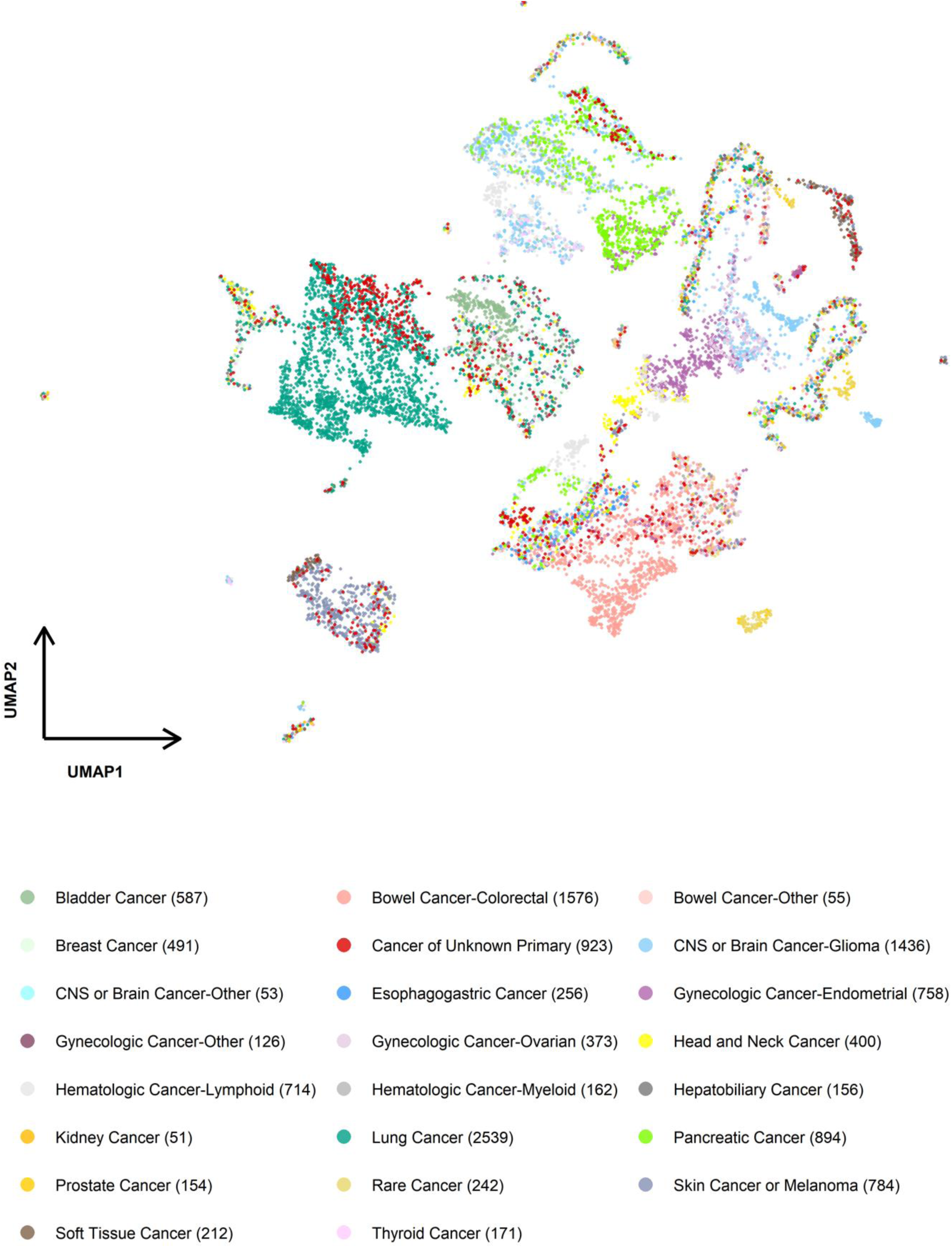
**SBS signature-burden landscape in Dana-Farber tumors**Uniform Manifold Approximation and Projection (UMAP) of SBS signature-burden profiles from 28,957 tumors sequenced at Dana-Farber Cancer Institute. Tumors were clustered within each cancer category using hierarchical clustering of signature-burden profiles, and UMAP was applied to the median profile of each cluster; each point therefore represents a tumor cluster rather than a single tumor. Points are colored by cancer type.

**Supplementary Fig 9.**
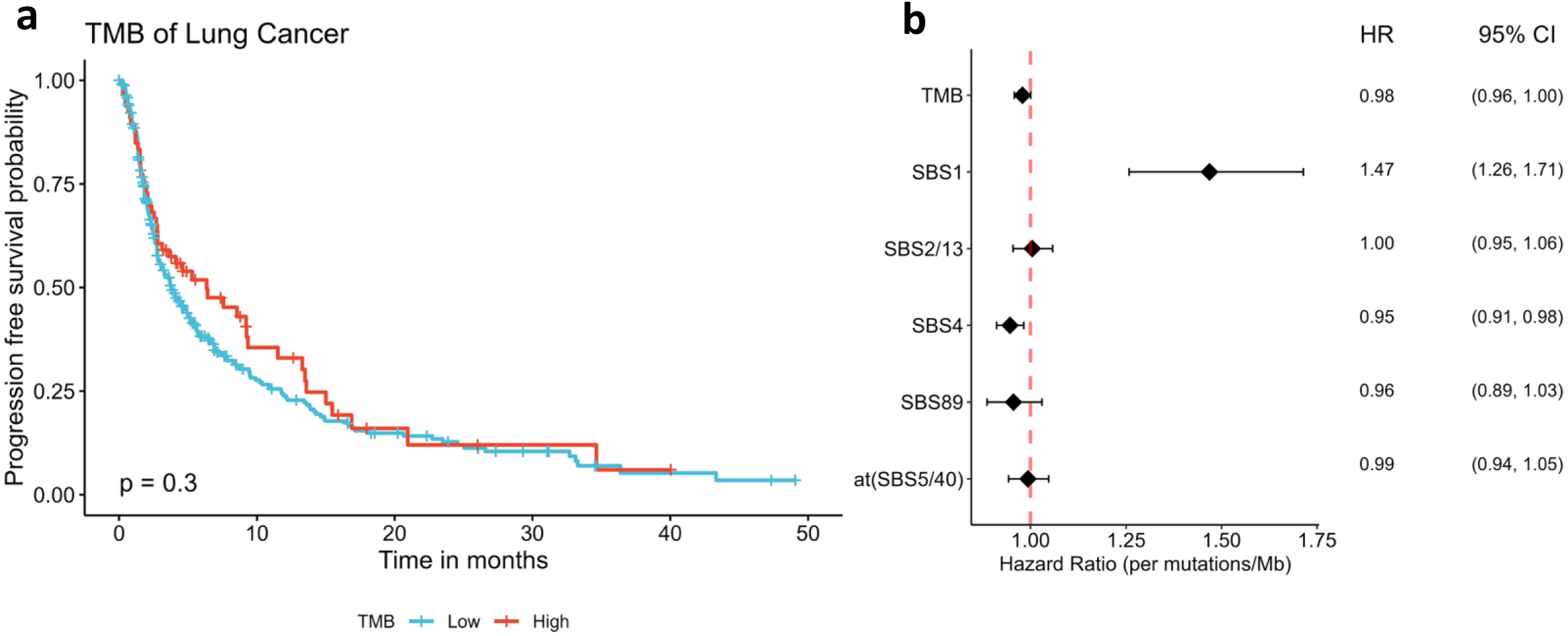
Mutational signatures and immune-checkpoint blockade outcomes. a, Kaplan-Meier curves show progression-free survival after immune-checkpoint inhibition in 470 patients with non-small cell lung cancer stratified by high (>10 mutations/Mb) versus low overall tumor mutational burden (TMB); the log-rank P value is shown. b, Forest plot shows hazard ratios (HRs) and 95% confidence intervals (CIs) for overall survival associated with signature-specific TMB in the same cohort. Mixed-effects Cox models adjusted for age, sex, smoking history, stage IV disease and signature-specific TMBs, with sequencing center and tumor subtype modeled as random effects.

**Supplementary Fig 10.**
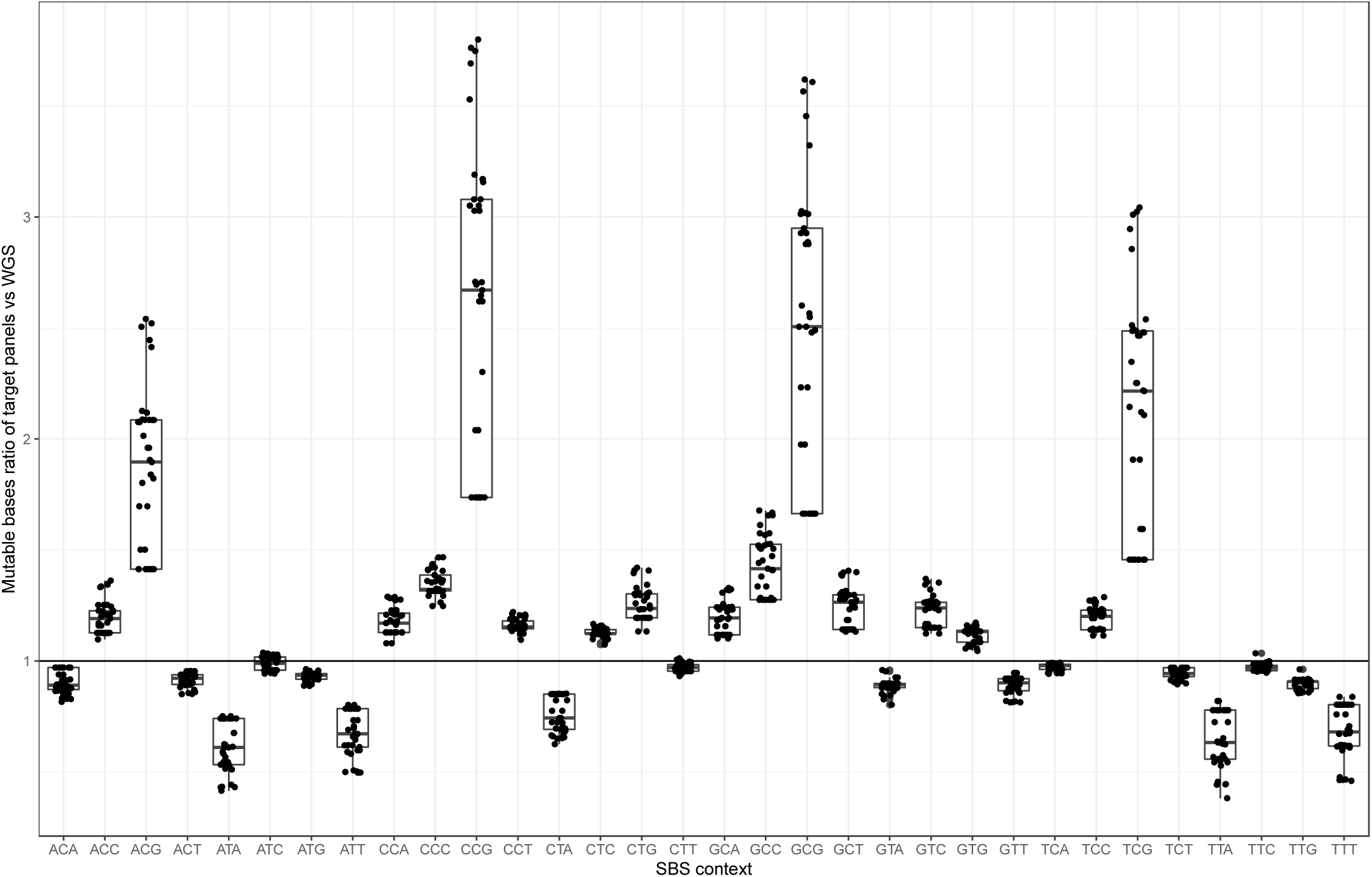
Trinucleotide-context opportunity in targeted panels and the genome. Box plots compare the proportion of each trinucleotide context in regions covered by 36 targeted panels with its proportion in the hg19 human reference genome. Each point represents one context- panel ratio; boxes show the median and interquartile range, and whiskers show 1.5 times the interquartile range. Values above or below one indicate enrichment or depletion in targeted-panel regions relative to the genome.

**Supplementary Fig 11.**
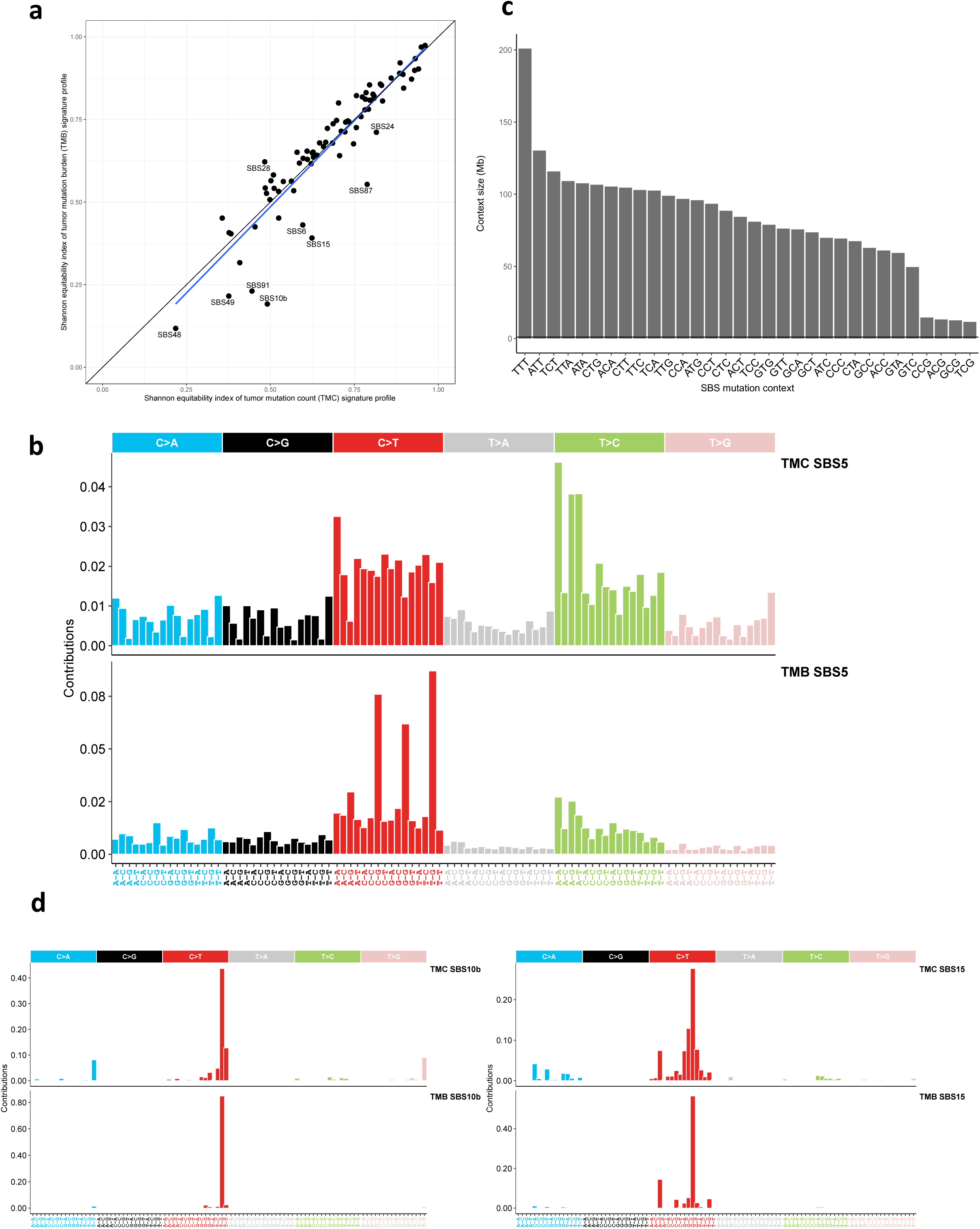
TMC and TMB signature profiles after panel-context adjustment. a, Shannon equitability indices for tumor mutational count (TMC) and tumor mutational burden (TMB) SBS signature profiles. Higher values indicate flatter profiles. TMB profiles were calculated after normalizing mutation counts by trinucleotide opportunity. The black diagonal denotes y = x; the blue line is a linear regression with a 95% confidence band. Signatures with an absolute TMC-TMB difference >0.1 are labeled. b, SBS5 profiles derived from TMC and TMB representations across 96 SBS mutation types. c, Frequencies of 32 trinucleotide contexts in the human genome, highlighting depletion of ACG, TCG, GCG and CCG contexts. d, SBS10b and SBS15 profiles derived separately from TMC and TMB representations.

**Supplementary Fig 12.**
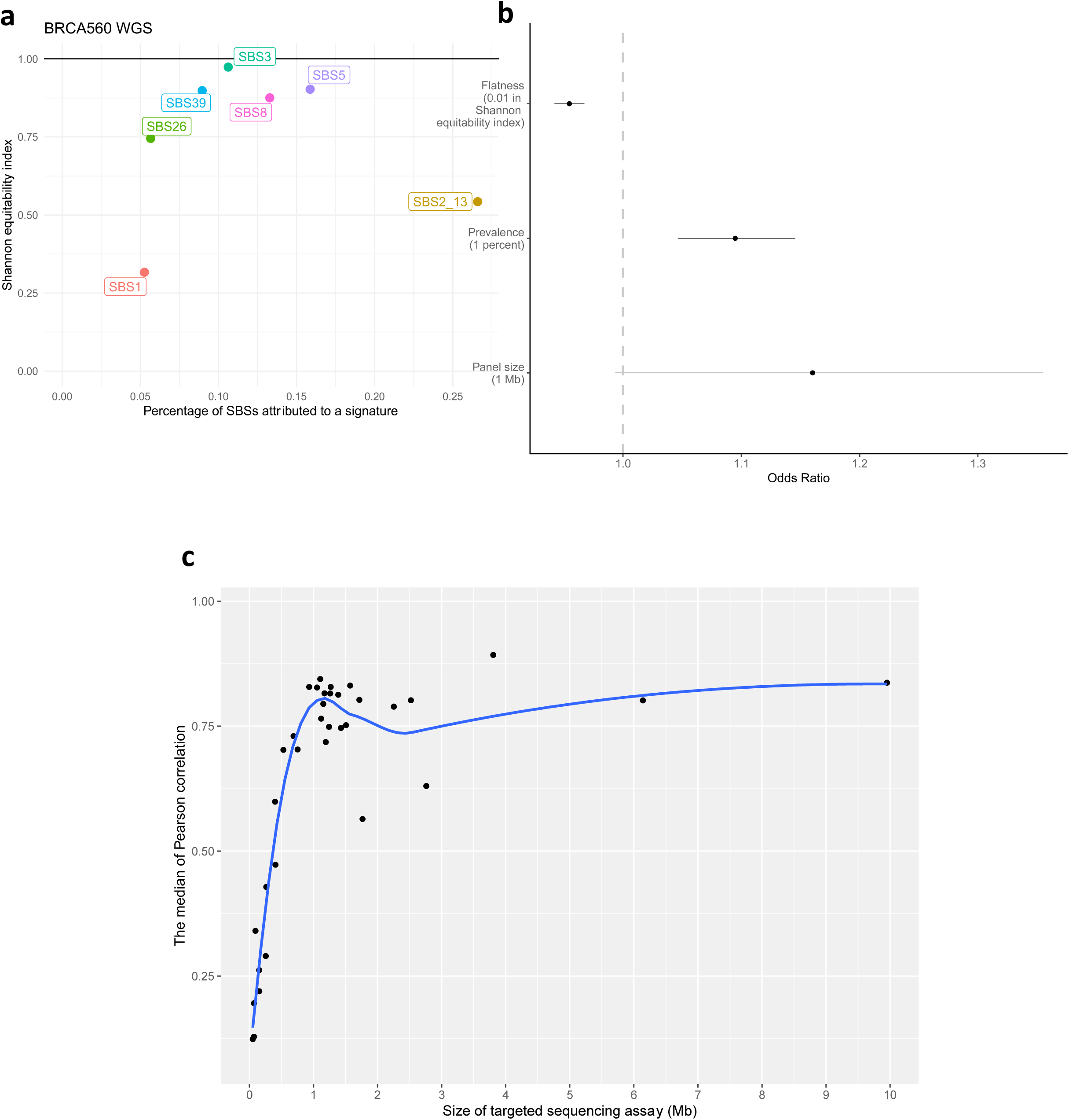
Pseudo-targeted sequencing analysis of the Sanger BRCA560 WGS cohort. a, Shannon equitability index versus the percentage of SBSs attributed to each signature in the Sanger BRCA560 WGS cohort; signatures present in >5% of tumors are colored and remaining signatures are gray. Pseudo-targeted datasets were generated by retaining WGS SBSs that intersected each of 36 targeted panels. b, Odds ratios and 95% CIs for associations of signature flatness, signature frequency and simulated panel size with detection probability. c, Median Pearson correlations between targeted-panel signature burdens and matched whole-genome burdens for each signature; the blue curve is a LOWESS fit with a 95% confidence band.

**Supplementary Fig 13.**
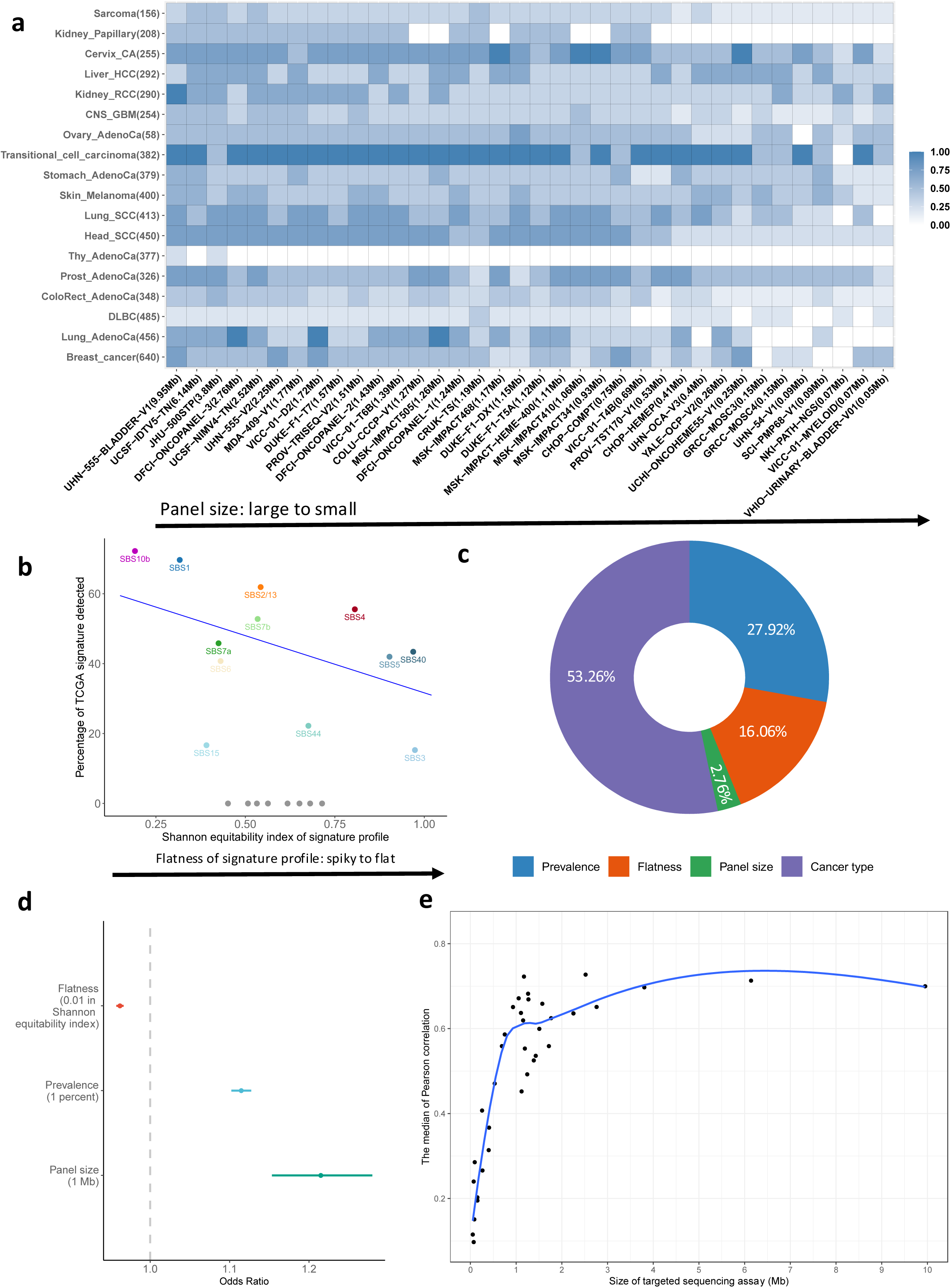
Determinants of SATS detection and burden-estimation performance. a, Detection probability for common TCGA WES signatures, defined as signatures contributing at least 5% of SBSs in a cancer type, across 648 pseudo-targeted datasets from 18 TCGA cancer types and 36 targeted panels. Values on the y-axis give the median effective sample size for each cancer type; panel sizes are shown on the x-axis. b, Detection probability plotted against Shannon equitability index; the blue line is a linear regression fit. Colored points indicate detected signatures and gray points indicate undetected signatures. c, Proportion of variance in detection probability explained by cancer type, signature flatness, signature prevalence and panel size in a generalized linear mixed model. d, Odds ratios and 95% CIs for the same determinants. e, Median Pearson correlation between WES-derived and pseudo-targeted SATS signature-burden estimates across 18 TCGA cancer types; the blue curve is a LOWESS fit with a 95% confidence band. Cancer-type abbreviations: CA, cervical; HCC, hepatocellular carcinoma; RCC, renal cell carcinoma; CNS, central nervous system; GBM, glioblastoma; AdenoCa, adenocarcinoma; SCC, squamous cell carcinoma; Thy, thyroid; Prost, prostate; Colorect, colon/rectum; DLBC, diffuse large B-cell lymphoma.

**Supplementary Fig 14.**
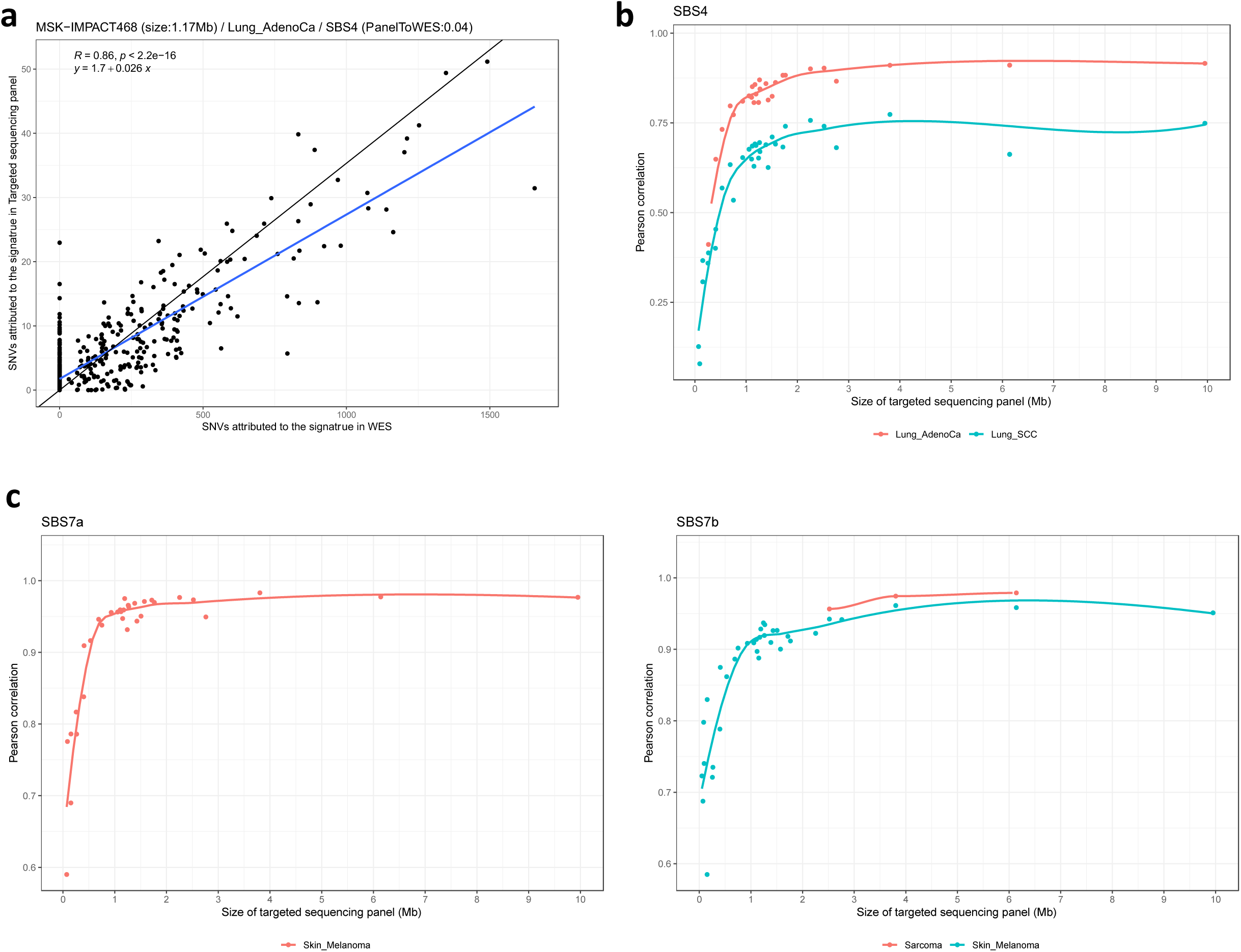
**Signature-burden estimation in pseudo-targeted TCGA WES data**a, SBS4-attributed mutation burdens estimated from pseudo-targeted MSK-IMPACT468 data are compared with SBS4 burdens from TCGA WES in the same lung cancer tumors. Pseudo-targeted data were generated by retaining TCGA WES SBSs that intersected panel-covered regions. The black line indicates the expected scaling by panel size relative to WES; the blue line is a linear regression fit with a 95% CI. b,c, Median Pearson correlations between pseudo-targeted and WES-derived burdens for SBS4 (b) and SBS7a/b (c) across panels. Blue curves are LOWESS fits with 95% confidence bands.

**Supplementary Fig 15.**
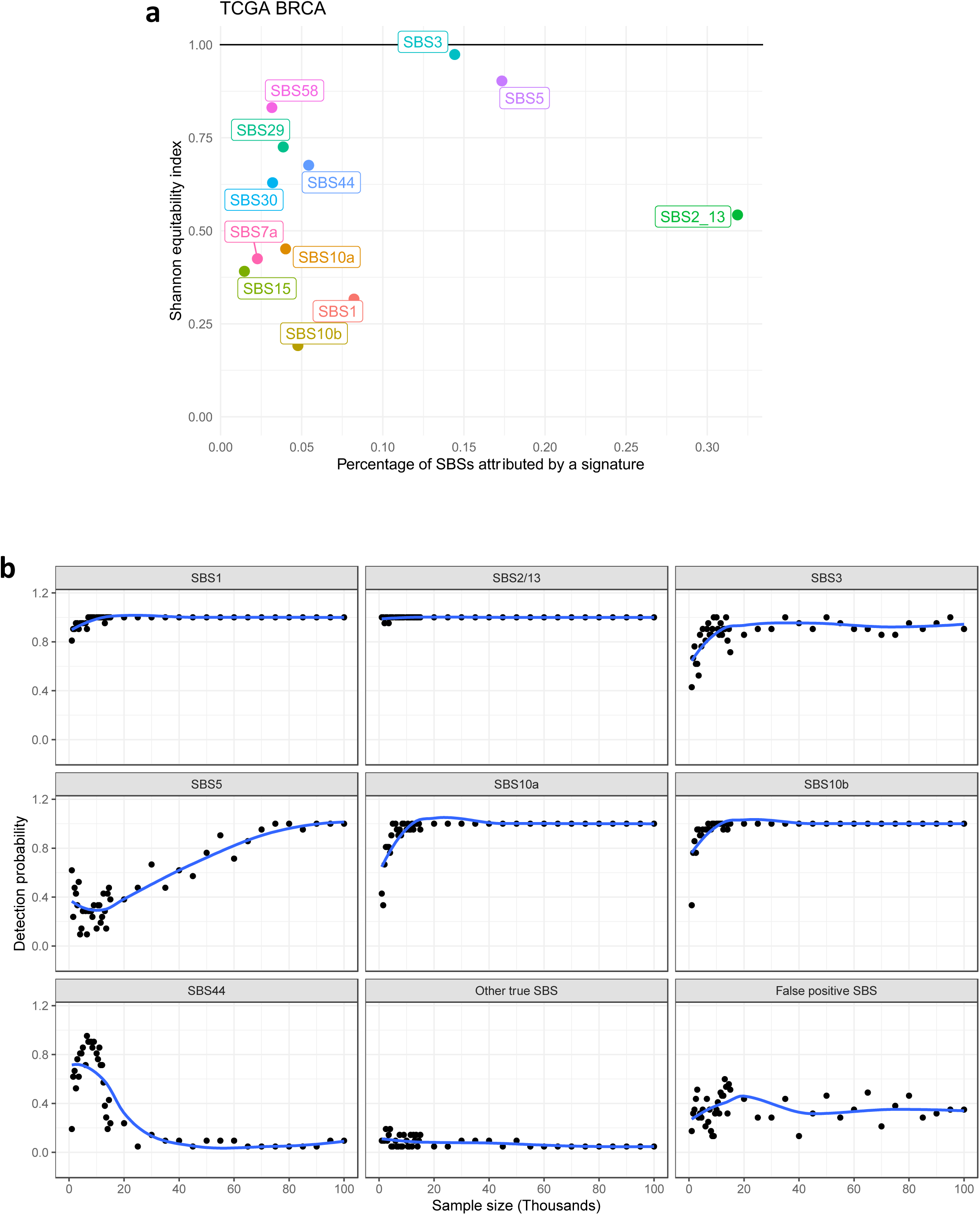
Sample-size effects on targeted-sequencing signature detection. a, Shannon equitability index versus SBS prevalence for 12 breast cancer signature groups derived from TCGA WES and contributing at least 1% of SBSs. The reference line at Shannon equitability index = 1 indicates the theoretical maximum for a completely flat profile. b, Detection probability across sample sizes from 1,000 to 1,000,000 tumors for targeted panels >1 Mb. Each point is the proportion of 21 panels in which the signature is detected at that sample size; the blue curve is a LOWESS fit with a 95% confidence band.

**Supplementary Fig 16.**
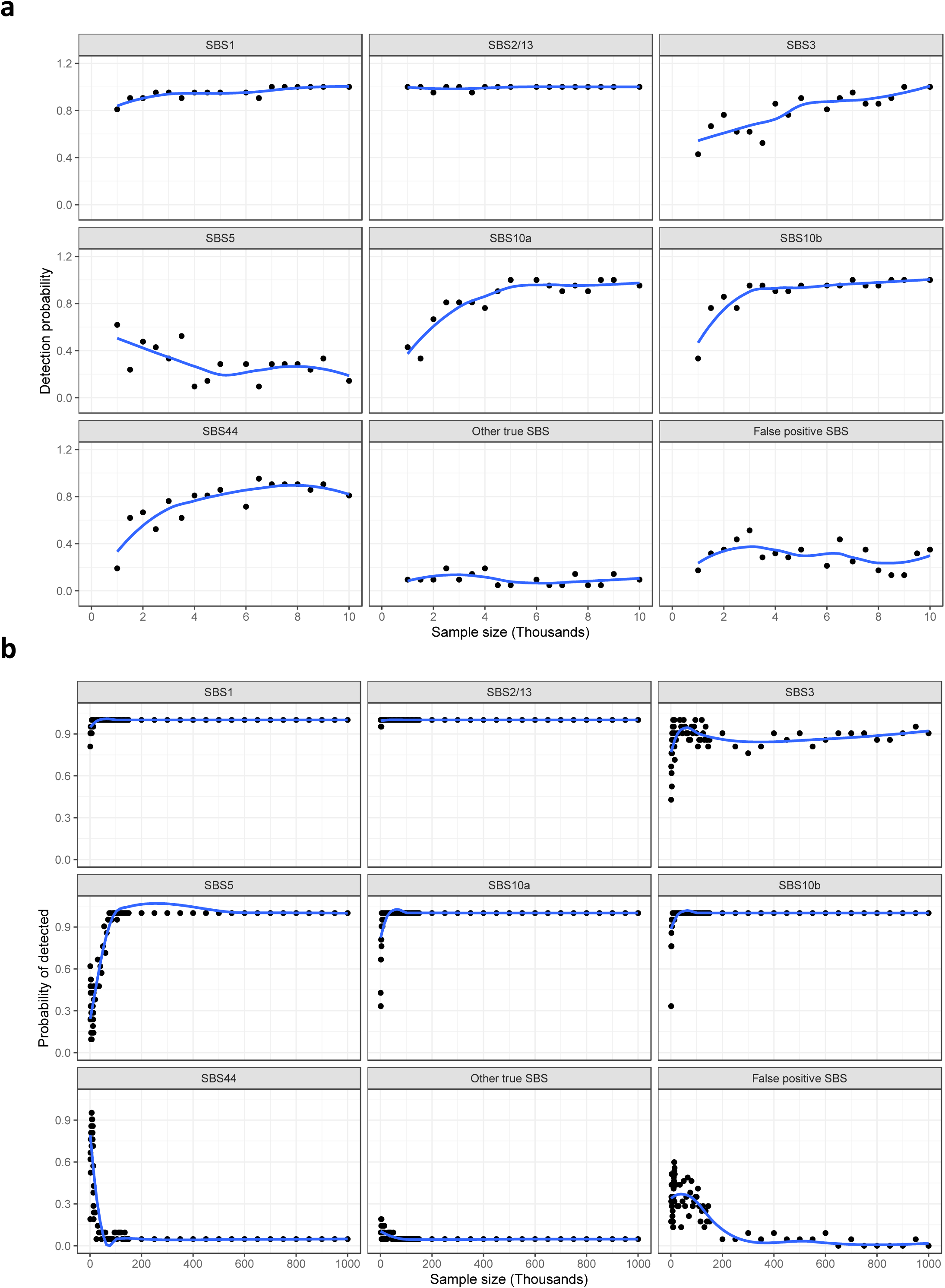
Detection probability and false-positive rate in breast cancer sample- size simulations. a, Detection probability for breast cancer signatures across increasing sample sizes up to 10,000 tumors. b, Detection probability across larger simulated cohorts up to 1,000,000 tumors, including false-positive detections. Simulations used 12 WES-derived breast cancer signature groups and 21 targeted panels >1 Mb. Each point represents the proportion of panels in which the signature is identified at the corresponding sample size. False positives are signatures not used to simulate the data but identified by de novo extraction and mapping. Blue curves are LOWESS fits with 95% confidence bands.

## Notes

### Competing Interest Statement

The authors have declared no competing interest.

### Author Declarations

The study used ONLY openly available human data that were originally located at https://synapse.org/genie, https://cancer.sanger.ac.uk/signatures/, https://gdc.cancer.gov/about-data/publications/mc3-2017, ftp://ftp.sanger.ac.uk/pub/cancer/Nik-ZainalEtAl-560BreastGenomes, https://www.synapse.org/#!Synapse:syn26706790, https://www.synapse.org/#!Synapse:syn11726618, and https://www.synapse.org/#!Synapse:syn11801497.

### Summary of Updates

A number of clinical application cases were added.

