## Supplementary material for "A real-world pan-cancer catalogue of mutational signatures from 111,711 tumors": Supp. Note

Table of Contents

*1. Limitations of existing methods*

*2. SATS estimates panel-calibrated tumor mutational burden signatures*

*3. Comparison of TMB and TMC signature profiles*

*4. Factors impacting signature detection and signature burden estimation by SATS*

**Signature detection performance**

**Signature burden estimation**

*5. Impact of sample sizes on SATS signature detection*

*6. Evaluation of SATS and other methods for signature detection*

**Benchmark design for signature-detection comparisons**

**SATS signature detection**

**SATS versus SigProfilerExtractor signature detection**

**SATS versus Mix signature detection**

*7. Evaluation of SATS and other methods for signature refitting*

**Benchmark design for signature-refitting comparisons**

**SATS versus signeR refitting**

**SATS versus sigLASSO refitting**

**SATS versus NNLS refitting**

**SATS versus MuSiCal refitting**

**SATS versus deconstructSigs refitting**

**SATS versus SigProfilerAssignment refitting**

*8. Evaluation of SATS and other methods in tumors with both WGS and targeted sequencing*

*9. Applicability of SATS to single-sample signature refitting*

*10. Impact of including irrelevant WES-derived signatures*

### 1. Limitations of existing methods

The analysis of mutational signatures in targeted sequencing data presents unique challenges, largely due to the heterogeneity of clinical panels and the sparse mutations they detect. Unlike whole-exome or whole-genome sequencing (WES/WGS), which provide more comprehensive mutational landscapes, targeted panels often cover distinct genomic regions and generate limited numbers of mutations per sample. As a result, traditional *de novo* signature extraction[1-3] or signature refitting methods[4,5], optimized for WES/WGS, are ill-suited to these heterogeneous and limited data.

De novo signature extraction requires a large sample size, sufficient mutational burden and consistent genomic coverage, conditions rarely satisfied in real-world targeted sequencing datasets. Conversely, signature refitting can be applied to limited samples, including individual tumors, by estimating the activities of predefined reference signatures. However, the accuracy of this method depends on selecting relevant reference signatures. Incorporating signatures not truly active in the tumor type can misassign mutations to irrelevant signatures6.

Although pan-cancer reference signatures derived from WES/WGS tumors, such as those from the COSMIC catalogue[6], are well-established, their applicability to targeted sequencing is limited.
Mutational signatures observed in WES/WGS studies may not accurately reflect those present in tumors profiled by targeted panels in real-world clinical settings. For example, treatment-induced signatures included in the COSMIC catalogue might be associated with different cancer types in the targeted sequencing studies. In addition, some rare signatures in the COSMIC catalogue are unlikely to be detected in targeted sequencing due to its narrow genomic coverage. Finally, some rare cancer subtypes that undergo targeted sequencing may lack WES/WGS-derived reference signatures altogether, making it challenging to refit signatures.

To address these limitations, recent methods have explored clustering-based strategies for identifying signatures in targeted data. SigMA[7], for instance, is tailored to detect the HRD-associated signature SBS3, requiring pre-training with WGS data from individual tumors. However, it is limited to the detection of a single signature and does not quantify multiple concurrent signature activities. The Mix9 method presents an alternative clustering strategy that does not rely on pre-training but estimates signature activities at the cluster rather than the individual sample level. Thus, specialized analytical methods and a comprehensive catalogue of mutational signatures tailored to tumors profiled by targeted sequencing are needed.

**2. SATS estimates panel-calibrated tumor mutational burden signatures**
A central feature of SATS is its ability to estimate tumor mutational burden (TMB) signature profiles without requiring uniform genomic coverage across tumor samples. Traditional signature analysis methods, such as those based on non-negative matrix factorization (NMF), assume that all samples are sequenced across the same genomic regions, typically through whole-exome or whole-genome sequencing. These methods decompose the mutation count matrix V into a product of a signature profile matrix W′ (96 mutation types × K signatures) and a signature activity matrix H′ (K signatures × N samples), that is, $V \approx W'H'$, where W′ characterizes raw tumor mutational count (TMC) signatures.

In contrast, SATS introduces a modified non-negative matrix factorization:

$$\mathbf{V} \approx \mathbf{L} \circ \mathbf{WH}$$

where L is the panel-context matrix and the product of L and WH is evaluated element-wise. The panel-context matrix records the mutation opportunity for each mutation type in the genomic regions covered by each panel, measured in megabases (Mb). This formulation allows SATS to adjust for panel design across targeted-sequencing assays. By incorporating L, SATS estimates W as a signature-profile matrix expressed as mutation burden per megabase, thereby capturing

TMB signatures rather than raw TMC signatures. This distinction is important in pan-cancer clinical cohorts that combine multiple gene panels with different genomic coverage. Because TMB signature profiles correct for variability in sequenced regions (Supplementary Fig. 10), they provide a panel-aware basis for signature analysis across targeted panels. The panel-aware pNMF derivation and grouped-extraction details are provided in the Supplementary Methods.

**3. Comparison of TMB and TMC signature profiles**

We assessed differences in the shape of TMB and TMC signature profiles using the Shannon equitability index (Supplementary Methods), where higher values indicate flatter, more uniform profiles, and lower values reflect more distinct or spiked patterns. Overall, the two types of profiles are broadly similar (Pearson correlation coefficient $r = 0.915$, Supplementary Fig. 11a). However, there are several notable discrepancies. For example, the TMC SBS5 profile exhibits a nearly uniform distribution across the 16 trinucleotide contexts of C>T mutations (Shannon equitability index $E_H = 0.941$). In contrast, the corresponding TMB profile ($E_H = 0.903$) shows elevated C>T mutations specifically at NCG trinucleotides (N represents any nucleotide, Supplementary Fig. 11b), since these trinucleotides are depleted in the human genome due to frequent deamination of 5-methylcytosine to thymine[10,11] (Supplementary Fig. 11c). In addition, TMB signature SBS10b and SBS15 ($E_H = 0.192$ and 0.391, respectively) exhibit more pronounced spikes compared to their TMC counterparts ($E_H = 0.491$ and 0.624, respectively, Supplementary Fig. 11d). These differences underscore the impact of incorporating panel-specific genomic context in shaping TMB signature profiles.

**4. Factors impacting signature detection and signature burden estimation by SATS**

We systematically evaluated factors influencing the performance of SATS in detecting signatures and estimating signature burdens from targeted sequencing data. Key variables considered included targeted gene panel size, signature prevalence, the shape of the TMB signature profile (quantified by the Shannon equitability index), and cancer type. To simulate realistic pseudo-targeted sequencing data, we subsampled SBSs from The Cancer Genome Atlas (TCGA) WES studies[7,12] and from WGS data of 560 breast tumors[13], restricting mutations to genomic regions covered by various targeted gene panels (Supplementary Methods, Supplementary Fig. 12a). Our analysis focused on common signatures contributing more than 5% of SBSs based on WES or WGS data for a given cancer type. While sample size may also affect signature detection, the limited availability of pseudo-targeted data precluded direct evaluation. We therefore conducted in silico simulations to assess its impact separately.

**Signature detection performance**

Using pseudo-targeted sequencing data based on WES, we found that SATS reliably detects common mutational signatures, though detection probabilities vary by cancer type, panel design, and individual signature characteristics. Larger gene panels generally uncovered more signatures (Supplementary Fig. 13a). We also observed a negative correlation between detection probability and the Shannon equitability index of signature profiles (Pearson $R = -0.452$; Supplementary Fig. 13b). This pattern is consistent with “spikier” signatures being more readily detected than flatter ones, as reported in WES/WGS analyses[14].

To jointly quantify the influence of multiple factors, we fit a generalized linear mixed model (GLMM) incorporating cancer type, panel size, signature prevalence, and profile flatness (Supplementary Methods). Cancer types accounted for 53.26% of the variance in detection probabilities on the logit scale (Supplementary Fig. 13c), likely reflecting differences in active signatures across tumor types. High-TMB cancer types, such as lung squamous cell carcinoma (median TMB: 10.07 mutations/Mb), showed greater signature detectability than low-TMB cancers such as thyroid adenocarcinoma (median TMB: 0.47 mutations/Mb).

Within cancer types, the GLMM further showed that spikier profiles, higher signature prevalence and larger panel size were each associated with higher detection probability (Supplementary Fig. 13d). Specifically, detection probability decreased as the Shannon equitability index increased (odds ratio (OR) = 0.962, 95% confidence interval (CI) = 0.956-0.967 for a 0.01 increase), whereas larger panels were associated with increased detection (OR = 1.21, 95% CI = 1.14-1.27 for a 1 Mb increase). These trends were also corroborated in analyses of pseudo-targeted sequencing data based on WGS (Supplementary Fig. 12b). These results help explain the challenge in detecting signatures in thyroid adenocarcinoma, where the spikier SBS1 signature is uncommon (prevalence, 6.58%), while the most common signature, SBS5 (28.26%), is flat and potentially confounded with other flat signatures such as SBS3 or SBS40.

**Signature burden estimation**

We next assessed SATS's performance in estimating signature burdens (defined as the number of mutations attributed to each signature) via its refitting step. We compared the signature burdens computed from WES data7 with those estimated using SATS from pseudo-targeted sequencing data of the same tumors (as an example, see Supplementary Fig. 14a for SBS4 in lung cancer based on the MSK-IMPACT468 panel). Strong concordance between these estimates would support the feasibility of using targeted sequencing as a surrogate for WES-derived signature burdens. We observed strong correlations for panels exceeding 1 Mb in size across signatures (median Pearson correlation coefficient r = 0.7; Supplementary Fig. 13e). Signature-specific results were even stronger, with SBS4 in lung adenocarcinoma achieving r = 0.91, and SBS7a and SBS7b in melanoma reaching r = 0.98 and 0.95, respectively (Supplementary Figs. 14b and 14c). Similar results were observed in pseudo-targeted sequencing data based on WGS (Supplementary Fig. 12c).

**5. Impact of sample sizes on SATS signature detection**

Pseudo-targeted sequencing datasets typically contained hundreds of tumors, which constrained the number of detectable mutational signatures. In contrast, real-world targeted sequencing cohorts often include thousands of tumors, potentially enhancing detection power. To explore the impact of sample size on mutational signature detection, we conducted in silico simulations using breast cancer as a case study. This setting consisted of 12 WES-derived breast cancer signature groups with prevalence of at least 1% in TCGA WES data (Supplementary Fig. 15a).

We simulated SBS mutation profiles across up to one million tumors, randomly generating mutations to 96 mutation types according to the true signature mixture observed in breast cancer. Simulations were performed across 21 targeted gene panels, each covering more than 1 Mb of the genome (Supplementary Methods). Because the ground truth signature composition was known *in silico*, we were able to benchmark SATS detection accuracy under varying sample sizes.

Our results showed that relatively spiky and prevalent signatures, such as SBS1 and SBS2/13, can be reliably detected with just a few thousand samples (Supplementary Fig. 15b). In contrast, less spiky or rarer signatures, such as SBS10a, required substantially more data for confident detection (Supplementary Fig. 16a). The flattest profiles, notably SBS3 and SBS5, required much larger cohorts, approximately 40,000 and 80,000 tumors, respectively, to be detected consistently across all panels (Supplementary Fig. 15b). Interestingly, the detection probability of SBS44 declined after surpassing 10,000 samples, coinciding with a rise in the detection of SBS5. This suggests that as flatter signatures (e.g., SBS3 and SBS5) become detectable, they may obscure or be confounded with other flat signatures like SBS44. This behavior mirrors observations from WGS-based analyses, where flat signatures are more prone to misclassification14. Signatures with prevalence below 5% remained largely undetectable even with large sample sizes (Supplementary Fig. 15b), consistent with prior findings using WGS and WES data1. Notably, the false-positive detection rate decreased markedly with increasing sample size, from 0.35 at 10,000 samples to below 0.01 at 200,000 samples (Supplementary Fig. 16b).

These simulations indicate that larger cohorts improve detection of most common breast cancer signatures, including the homologous recombination deficiency (HRD)-associated SBS3 under the simulated settings, while some flat or rare signatures remain difficult. They also indicate that cohort size is a key determinant of mutational-signature extraction from targeted sequencing data.

### 6. Evaluation of SATS and other methods for signature detection

We next evaluated active-signature detection, defined here as identifying which mutational signatures are present in targeted-sequencing mutation counts. This analysis differs from the refitting comparisons in Section 7 because the primary endpoint is whether the ground-truth signatures are detected, not whether their burdens are estimated after a candidate set has already been supplied.

#### Benchmark design for signature-detection comparisons

We conducted in silico simulations across four cancer types: lung, breast, colorectal and lymphoid-derived hematologic malignancies. For each cancer type, mutation counts were generated from prespecified mutational signatures, providing a known ground-truth signature set. SATS, SigProfilerExtractor and Mix were then applied to the same simulated datasets, and the proportion of 10 replicates in which each true signature was detected was compared across methods (Supplementary Fig. 3). Asterisks in Supplementary Fig. 3 denote Kruskal-Wallis rank-sum tests comparing detection proportions between methods. Simulation and comparator implementation details are provided in the Supplementary Methods.

#### SATS signature detection

SATS showed consistently high detection sensitivity across the simulation benchmarks (Supplementary Fig. 3). In lung cancer, SATS detected all evaluated true signatures in every replicate, including SBS29 and SBS89, which were not detected by the comparator methods. In breast cancer, SATS, SigProfilerExtractor and Mix all detected SBS1, SBS2/13 and the flat SBS5/SBS40 component in every replicate. In colorectal cancer, SATS detected SBS1, SBS6, SBS10a, SBS10b and the flat SBS5/SBS40 component in every replicate, whereas SBS44 was detected less frequently, consistent with the expected difficulty of resolving this relatively flat signature from other flat profiles. In lymphoid-derived hematologic cancers, SATS detected SBS1, SBS6, SBS84 and the flat SBS3/SBS5/SBS40 component in every replicate and retained high, but not complete, detection for SBS19, SBS46 and SBS87.

**SATS versus SigProfilerExtractor signature detection**

SigProfilerExtractor matched SATS for several high-signal or broadly detectable signatures, including all three breast cancer signatures and multiple colorectal signatures (Supplementary Fig. 3). However, SigProfilerExtractor failed to detect the lung-specific signatures SBS29 and SBS89 in this benchmark and did not detect colorectal SBS44. In lymphoid-derived hematologic cancers, SigProfilerExtractor showed reduced detection for selected signatures, including SBS6, SBS46 and SBS87, although it retained high detection for SBS1, SBS19, SBS84 and the flat component. These results indicate that SigProfilerExtractor performed well for some strong signals but was less reliable for several cancer-type-specific or more difficult signatures in the targeted-panel simulations.

**SATS versus Mix signature detection**

Mix also detected several common signatures, including all three breast cancer signatures and the flat components in lung, colorectal and lymphoid-derived hematologic cancers (Supplementary Fig. 3). In lung cancer, however, Mix did not detect SBS29 or SBS89, paralleling the SigProfilerExtractor result. In colorectal cancer, Mix showed lower detection proportions than SATS for SBS6, SBS10a, SBS10b and SBS44. In lymphoid-derived hematologic cancers, Mix detected SBS1, SBS84 and the flat component, but did not detect several other simulated lymphoid signatures, including SBS6, SBS19, SBS46 and SBS87. Thus, Mix recovered some dominant or broadly detectable signals but showed lower sensitivity than SATS for multiple signatures in colorectal and lymphoid-derived hematologic simulations.

Overall, these detection benchmarks show that SATS recovered the prespecified active signatures more consistently than SigProfilerExtractor or Mix in several targeted-panel simulation settings, particularly for lung SBS29/SBS89, colorectal SBS6/SBS10a/SBS10b/SBS44 and multiple lymphoid-associated signatures (Supplementary Fig. 3).

### 7. Evaluation of SATS and other methods for signature refitting

We next evaluated signature refitting, defined here as estimating the contribution of prespecified mutational signatures to observed targeted-sequencing mutation counts. We separated these analyses from signature detection because refitting methods start from a candidate signature set, whereas SATS first extracts and maps active signatures before estimating burdens. The comparisons below therefore focus on burden estimation after candidate signatures have been specified.

**Benchmark design for signature-refitting comparisons**

The refitting comparisons used a common benchmark logic across four cancer types: lung, breast, colorectal and lymphoid-derived hematologic malignancies. For each cancer type, simulated targeted-sequencing mutation-type matrices were generated from mutational signature profiles and activity distributions derived from AACR Project GENIE data, providing known ground-truth signature burdens for comparison (Supplementary Methods). Each method was then asked to estimate signature burdens from targeted-panel mutation counts, and the refitted burdens were compared with the simulated truth.

We applied this benchmark design at two levels. First, because SATS uses signeR for de novo signature extraction but implements a separate panel-aware EM procedure for burden estimation, we compared SATS with signeR using the same mapped signatures and quantified the mean squared error (MSE) between estimated and true burdens across simulation replicates (Supplementary Fig. 2). Second, to stress-test broad-catalogue refitting specificity in targeted-panel data, representative 100-tumor composites from the same cancer-type simulation framework were refitted with the full TMC signature set using sigLASSO, NNLS, MuSiCal, deconstructSigs and SigProfilerAssignment (Supplementary Fig. 6). Because these refitting-only methods require a prespecified signature catalogue and do not implement the de novo extraction and recurrent pNNLS mapping steps used by SATS, the Supplementary Fig. 6 analysis should be interpreted as an exploratory specificity analysis rather than a formal end-to-end workflow benchmark. Implementation details for the simulation framework and comparator methods are provided in the Supplementary Methods.

**SATS versus signeR refitting**
Using the same mapped signatures for each cancer type, we evaluated how well SATS and signeR recovered simulated signature burdens. Across most signatures and cancer types, SATS had lower MSE than signeR, consistent with improved refitting accuracy in these simulations (Supplementary Fig. 2). The difference was most apparent in higher-error settings, including flat signatures, whereas several low-error signatures showed smaller or non-significant differences. These results support the panel-aware EM burden-estimation step used by SATS beyond the signeR extraction component.

**SATS versus sigLASSO refitting**
sigLASSO uses regularization to encourage sparse signature assignments. Under the broad TMC candidate set, it recovered several major smoking- and APOBEC-related or clock-like components, but still assigned substantial burden to non-ground-truth signatures (Supplementary Fig. 6). Examples included off-target assignments to SBS23, SBS34 and SBS91 in lung cancer, SBS37 and SBS87 in breast cancer, and additional non-ground-truth components in colorectal and lymphoid-derived hematologic composites. Thus, sigLASSO improved sparsity relative to unconstrained refitting but did not fully control false-positive attribution when the candidate catalogue was large.

**SATS versus NNLS refitting**
NNLS served as the unconstrained non-negative least-squares baseline. As expected, it produced the most diffuse profiles, distributing non-zero burden across many candidate signatures rather than concentrating burden on the simulated ground-truth set (Supplementary Fig. 6). Although dominant true signatures were often represented, additional low- and high-level off-target peaks appeared across lung, breast, colorectal and lymphoid-derived hematologic composites. This pattern indicates that broad-catalogue NNLS refitting is vulnerable to over-attribution in targeted-panel data unless the candidate set is biologically restricted before refitting.

**SATS versus MuSiCal refitting**
MuSiCal recovered portions of the dominant simulated signals but produced a diffuse tail of non-ground-truth assignments when refitted against the full TMC catalogue (Supplementary Fig. 6). Across lung, breast, colorectal and lymphoid-derived hematologic composites, major signatures were captured to varying degrees, but burden was spread across a larger set of candidate signatures than in SATS. These results suggest that MuSiCal can recover dominant components, but its specificity depends on candidate-set control in the targeted-sequencing setting.

**SATS versus deconstructSigs refitting**
deconstructSigs produced more concentrated profiles than NNLS and MuSiCal in some panels, but it still assigned appreciable burden to signatures absent from the simulated ground truth (Supplementary Fig. 6). In lung and breast cancer, it captured several expected components while adding off-target contributions, including SBS24, SBS39 and SBS91 in lung cancer and SBS7a, SBS36 and SBS87 in breast cancer. Similar additional assignments appeared in colorectal and lymphoid-derived hematologic composites. This behavior was less pronounced when comparisons were restricted to a biologically focused signature set in the matched kidney WGS/targeted analysis (Supplementary Fig. 7), emphasizing that deconstructSigs performance is sensitive to the supplied candidate catalogue.

**SATS versus SigProfilerAssignment refitting**
SigProfilerAssignment was generally more concentrated than NNLS and MuSiCal, but the full-catalogue stress test still showed off-target attribution and incomplete recovery of some simulated components (Supplementary Fig. 6). In lung cancer, it assigned substantial burden to several expected signatures while adding non-ground-truth components; in breast cancer, the dominant flat and APOBEC-related contributions were concentrated more clearly than with the most diffuse comparators, but extra signatures remained. In colorectal and lymphoid-derived hematologic composites, SigProfilerAssignment captured several high-burden true signatures but also over- or under-estimated selected components and assigned burden outside the simulated set. These findings indicate that SigProfilerAssignment can perform well when the candidate set is appropriate, but broad-catalogue refitting alone does not replace the panel-aware candidate selection and mapping steps used by SATS.
Together, these refitting analyses support SATS as an integrated workflow when evaluated on outputs matched to its intended task: active-signature detection, recurrent mapping to a concise candidate set and panel-aware burden estimation. The signeR comparison supports improved burden estimation within the SATS framework, whereas the broader refitting-only comparisons show that large prespecified candidate sets can recover dominant true signatures but can also over-assign burden to irrelevant signatures. These results define a practical limitation of broad-catalogue refitting in panel data rather than a general claim that SATS is superior to all refitting tools in every use case.

**8. Evaluation of SATS and other methods in tumors with both WGS and targeted sequencing**
To further evaluate SATS, we analyzed 72 kidney tumors that had undergone both WGS and targeted sequencing. Given the moderate cohort size, we focused on signature refitting using a restricted set of common signatures (SBS1, SBS5 and SBS40) previously identified in kidney tumors from the AACR Project GENIE, as well as through WGS-based mutational signature

analyses of 72 kidney tumors15. We compared SATS to three established methods: SigProfilerAssignment, Mix and deconstructSigs. Experimental and analytical details for this paired WGS/targeted benchmark are provided in the Supplementary Methods.

To avoid dichotomizing tumors by targeted-sequencing burden, we replotted this matched-platform benchmark as continuous targeted-sequencing burden versus log1p-transformed matched WGS burden for each method (Supplementary Fig. 7). Cross-platform concordance was quantified using Spearman correlation and leave-one-out cross-validated $R^2$ from linear models predicting log1p(WGS burden) from targeted-sequencing burden. For SBS1, SATS showed the strongest concordance with matched WGS burden (Spearman rho = 0.55; cross-validated $R^2$ = 0.26), whereas SigProfilerAssignment showed little association (rho = 0.04; cross-validated $R^2$ = -0.03), Mix showed weaker association (rho = 0.28; cross-validated $R^2$ = 0.01), and deconstructSigs assigned a constant SBS1 burden of zero, precluding correlation estimation. For the flat SBS5/SBS40 category, all methods showed positive concordance with matched WGS burden: SATS had rho = 0.48 and cross-validated $R^2$ = 0.21, while SigProfilerAssignment, Mix and deconstructSigs had rho values of 0.59-0.61 and cross-validated $R^2$ values of 0.29-0.33. Thus, the paired WGS/targeted benchmark suggests that SATS better resolved the less prevalent SBS1 signal under sparse targeted-panel counts, whereas the broad, high-burden flat SBS5/SBS40 component was captured by all evaluated refitting methods.

**9. Applicability of SATS to single-sample signature refitting**
SATS can perform signature refitting at the resolution of individual tumors, provided that a predefined set of relevant signatures for the cancer type is available. To illustrate this capability, we performed *in silico* simulations of lung cancer targeted-sequencing data and estimated signature burdens using SATS on varying subset sizes, including single-sample scenarios. Simulation details are provided in the Supplementary Methods.

We observed high concordance between estimated and true simulated signature burdens across all subset sizes, including single-sample scenarios (Supplementary Fig. 5). These results support tumor-level signature-burden estimation under the simulated conditions and indicate that single-sample refitting is feasible when an appropriate cancer-type-specific reference set is available.

**10. Impact of including irrelevant WES-derived signatures**
To assess the consequences of including irrelevant signatures during refitting, we simulated breast cancer targeted-sequencing data using three known signatures: SBS1, SBS2/13 and SBS5. Signature refitting was then performed using a broader set of 12 WES-derived breast cancer signature groups drawn from the TCGA WES breast cancer signature study, which included the three true signatures alongside nine extraneous ones. Simulation and refitting details are provided in the Supplementary Methods.

This analysis showed that, when the broader WES-based signature set was used, signatures not present in the simulated data were assigned to a substantial fraction of tumors (Supplementary Fig. 4), highlighting the risk of overfitting when irrelevant signatures are included. These findings underscore the importance of selecting a reference signature set calibrated to targeted sequencing to support accurate mutational-signature estimation in panel-based studies.
