## Supplementary material for "A real-world pan-cancer catalogue of mutational signatures from 111,711 tumors": Supp. Methods

### AACR Project GENIE cohort and mutation filtering

The primary targeted-sequencing analysis used AACR Project GENIE version 13.0-public, obtained from Synapse. The dataset includes tumors sequenced during routine clinical care at 16 participating hospitals or cancer centers in North America and Europe. When multiple tumors were available from the same patient, one tumor was selected at random to avoid over-representing patients with repeated sampling. Tumors sequenced with gene panels covering ≤ 50 kb of genomic region were excluded because very small panels provide too few mutation opportunities for stable signature analysis.

Sequencing was performed in CLIA- and/or ISO-certified laboratories with high read depth (median read depth, 519×; interquartile range, 307× to 808×). Somatic variants were called by participating centers using local pipelines that included tools such as Mutect2 and Strelka, followed by filtering of germline variants and artefacts using external control sets and germline-variant resources such as gnomAD. For this analysis, we further required somatic mutations to have read depth of at least 100 and alternate allele count of at least 5. After these filters, 982,095 SBSs and 15,149 DBSs from 111,711 tumors were retained for mutational-signature analysis.

Differences in local mutation-calling pipelines can affect individual variant calls, but the SATS workflow is based on mutation-type counts aggregated across tumors and panels. We therefore treated the filtered GENIE mutation table as the analysis input and modeled panel-specific mutation opportunity explicitly, rather than assuming that all tumors had been sequenced over the same genomic territory.

### Cancer-type and subtype harmonization

Tumors were annotated using OncoTree cancer-type and subtype labels. The GENIE release contained 102 cancer types and 757 cancer subtypes; for cancer-type-specific signature analysis, related OncoTree labels were consolidated into 23 cancer categories used for downstream analysis. For example, breast cancer, breast cancer not otherwise specified and breast sarcoma were grouped into the breast cancer category. Within each cancer category, subtypes representing less than 3% of tumors were combined into a rare-subtype category to reduce instability in subtype-specific models.

Clinical and technical covariates used in downstream analyses included sequencing center, tumor status (primary, metastatic or unspecified/hematologic), age, sex and race when available. Sex was not included as a covariate for sex-specific cancer groups, including breast, ovarian, endometrial, other gynecologic and prostate cancers.

### Construction of targeted-panel mutation opportunity matrices

For each targeted-sequencing panel, genomic coordinates were obtained from the AACR Project GENIE panel definition file (Synapse ID: syn26706790). Panel-covered DNA sequences were extracted from the hg19 human reference genome, and mutation opportunities were counted for each SBS and DBS class. For SBS analysis, we used the standard 96-channel representation defined by six base-substitution classes (C>A, C>G, C>T, T>A, T>C and T>G) within the 16 possible trinucleotide contexts around the mutated pyrimidine base. The corresponding 32 trinucleotide contexts were also tabulated because they determine the panel-specific opportunity available for each SBS class.

For DBS analysis, we considered 78 DBS mutation types arising from 10 dinucleotide contexts: AC, AT, CC, CG, CT, GC, TA, TC, TG and TT. Each tumor inherited the opportunity vector of the panel used to sequence it. These opportunity counts formed the panel context matrix used throughout SATS, allowing mutation counts from panels of different sizes and target compositions to be analyzed in a common likelihood framework.

**Panel-aware pNMF model and grouped de novo signature extraction**

SATS models targeted-sequencing mutation counts with a panel-aware Poisson non-negative matrix factorization (pNMF). For each tumor and mutation type, the expected mutation count is represented as the product of the panel-specific mutation opportunity and the sum of signature-specific contributions. This formulation distinguishes the biological activity of a signature from the number of genomic contexts in which that signature could have been observed on the panel. The canonical NMF special case is derived below.

**Derivation of the canonical NMF special case.**

When all tumors are sequenced over the same genomic region, the panel opportunity term is constant across samples. Under this condition, the panel-aware pNMF likelihood reduces to the canonical NMF objective based on generalized Kullback-Leibler divergence. The derivation below shows this relationship and motivates the use of the pNMF model as a direct extension of standard NMF for heterogeneous targeted-panel data.

When genomic regions are the same across tumors, the log-likelihood function in equation (1) simplifies to:

$$\log\{P(\mathbf{V}|\mathbf{L},\mathbf{W},\mathbf{H})\} = -D_{KL}(\mathbf{V}|\mathbf{W}',\mathbf{H}) + C,$$

where $w'_{pk} = \ell_p w_{pk}$, $C$ is a constant irrelevant to $\mathbf{W}$ and $\mathbf{H}$ and

$$D_{KL}(\mathbf{V}|\mathbf{W}',\mathbf{H}) = \sum_{n=1}^{N}\sum_{p=1}^{96}\left\{v_{pn}\log\left(\frac{v_{pn}}{\sum_{k=1}^{K} w'_{pk}h_{kn}}\right) + \sum_{k=1}^{K} w'_{pk}h_{kn} - v_{pn}\right\} \quad (2)$$

is the generalized Kullback–Leibler divergence. This is the standard objective function minimized in canonical NMF[1], confirming that the pNMF model includes NMF as a special case. De novo TMB signature profiles were extracted with signeR under the pNMF model. Because signeR uses a computationally intensive Markov chain Monte Carlo procedure, tumors were grouped into sets of 100 before extraction in the primary GENIE analysis. For SBS signatures, candidate ranks 1 to 5 were evaluated for each cancer category, except for CNS/brain cancer-glioma, for which ranks 1 to 6 were evaluated. For DBS signatures, candidate ranks 1 to 3 were evaluated. The selected rank was taken from the signeR model-selection output, which includes BIC profiles and the selected number of signatures. A fixed random seed of 4022022 was used for the primary extraction.

The grouping step preserves the pNMF likelihood structure because the sum of independent Poisson mutation counts is also Poisson distributed. Within each group, mutation counts and panel opportunities are aggregated, whereas the underlying TMB signature profile remains invariant. This substantially improves computational feasibility; in the main analysis, grouping 10,000 tumors into sets of 100 reduced the runtime from approximately 13 hours without grouping to 28.5 minutes on a standard laptop with an Intel Core i7-1165G7 processor and 16 GB RAM.

**Grouped-sample likelihood derivation.**

For each group of tumors, SATS aggregates mutation counts and panel opportunities before de novo extraction. Because independent Poisson counts remain Poisson after summation, the grouped count retains the same likelihood form as the individual-tumor model. The equations below define the grouped counts, aggregated panel opportunity and panel-size-weighted activity used in this argument.

Specifically, let C = {1, 2, ..., N} denote the index set of all samples, and let C_m (m = 1, ..., M) denote mutually exclusive subsets whose union is C. For each group C_m, we define the aggregated mutation counts as:

$$v_{pm}^{\#} = \sum_{n \in C_m} v_{pn},$$

which is the total mutation count for mutation type p across all tumors in group $C_m$. The expected value of $v_{pm}^{\#}$ is:

$$E\left(v_{pm}^{\#}\right) = \sum_{k=1}^{K} \sum_{n \in C_m} e_{kpn} = \sum_{k=1}^{K} l_{pm}^{\#} w_{pk} h_{km}^{\#},$$

where $l_{pm}^{\#} = \sum_{n \in C_m} \ell_{pn}$ is the aggregated panel size for group $C_m$, and $h_{km}^{\#} = \frac{\sum_{n \in C_m} \ell_{pn} h_{kn}}{\sum_{n \in C_m} \ell_{pn}}$ is the panel-size–weighted average signature activity. The TMB signature profile $w_{pk}$ remains invariant under this grouping. Since the sum of independent Poisson random variables is also Poisson distributed, the grouped mutation count $v_{pm}^{\#}$ follows a Poisson distribution. This property ensures that sample grouping preserves the statistical structure required for valid signature extraction under the pNMF model.

**Reference TMB signatures and mapping of de novo profiles**

The de novo signatures estimated from targeted-sequencing data represent TMB signature profiles rather than the usual tumor mutational count (TMC) profiles. To create compatible reference profiles for mapping, COSMIC SBS and DBS signature profiles (version 3.4) were transformed from TMC signatures to TMB signatures by dividing each mutation-type contribution by the number of genomic contexts in which that mutation type can occur in the human reference genome and then renormalizing each signature to sum to one.

The SBS context counts used for this conversion are provided in Supplementary Table 4, the resulting SBS reference TMB signatures in Supplementary Table 6, the DBS context counts in Supplementary Table 5 and the resulting DBS reference TMB signatures in Supplementary Table 7.

Detected de novo TMB signatures were mapped to COSMIC reference TMB signatures by penalized non-negative least squares. The penalty promotes sparsity, which is important because targeted panels often observe too few mutations to cleanly separate mixtures of similar known processes. The tuning parameter was selected by cross-validation. To reduce instability from cross-validation and finite mutation counts, the mapping procedure was repeated 100 times, and a reference signature was retained only if its coefficient exceeded 0.1 in at least 80 of the 100 repetitions. This recurrent mapping rule produced a stable candidate set for downstream refitting.

#### EM refitting and signature-burden estimation

After mapping, SATS refits the selected reference TMB signatures to individual tumors using an expectation-maximization (EM) algorithm. The observed mutation-type count for each tumor is modeled as the sum of latent signature-specific counts. In the E-step, the latent counts are allocated across signatures according to their current expected contributions, conditional on the observed mutation count and the panel opportunity matrix. In the M-step, the signature activities are updated to maximize the expected complete-data log-likelihood.

**EM update derivation.**
The EM derivation treats the signature-attributed mutation counts as latent variables. This construction gives a complete-data log-likelihood, an E-step that allocates each observed mutation type across reference signatures according to current expected contributions and an M-step with a closed-form update for the activity of each signature in each tumor.
We developed an expectation-maximization (EM) algorithm to estimate the signature activity matrix H, given the mutation-type matrix V, the panel context matrix L and the mapped reference TMB signature profiles W. For each tumor, the observed count for mutation type p is modeled as the sum of latent counts attributed to K signatures. These latent counts are treated as missing data and are modeled as Poisson random variables.

$$v_{kpn} \sim Poisson\left(\ell_{pn} w^*_{pk} h_{kn}\right), \qquad k = 1,2,\dots,K.$$

Introducing latent counts enables computation of the complete data log-likelihood as:

$$\sum_{p=1}^{96} \sum_{n=1}^{N} \sum_{k=1}^{K} \left\{-\ell_{pn} w^*_{pk} h_{kn} + v_{kpn}\, log\left(\ell_{pn} w^*_{pk} h_{kn}\right) - log\left(v_{kpn}!\right)\right\}.$$

In addition, the conditional distribution of $v_{kpn}$, given $\mathbf{V}$, $\mathbf{L}, \mathbf{W}^*$ and $\mathbf{H}^t$ (the $\mathbf{H}$ at the $t$'th iteration of the EM algorithm), follows a multinomial distribution with parameters $v_{pn}$ and $p_k = w^*_{pk} h^t_{kn} / \sum_{j=1}^{K} w^*_{pj} h^t_{jn}$.

In the E-step, we compute $Q(\mathbf{H}|\mathbf{H}^t)$ as the expected complete data log-likelihood:

$$\begin{aligned} Q(\mathbf{H}|\mathbf{H}^t) &= E\left[\sum_{p=1}^{96} \sum_{n=1}^{N} \sum_{k=1}^{K} \left\{-\ell_{pn} w^*_{pk} h_{kn} + v_{kpn}\, log\left(\ell_{pn} w^*_{pk} h_{kn}\right)\right\} | \mathbf{V}, \mathbf{L}, \mathbf{W}^*, \mathbf{H}^t\right] \\ &= \sum_{p=1}^{96} \sum_{n=1}^{N} \sum_{k=1}^{K} \left\{-\ell_{pn} w^*_{pk} h_{kn} + \log\left(\ell_{pn} w^*_{pk} h_{kn}\right) v_{pn} \frac{w^*_{pk} h^t_{kn}}{\sum_{j=1}^{K} w^*_{pj} h^t_{jn}}\right\}. \end{aligned}$$

In the M-step, the maximizer of $Q(\mathbf{H}|\mathbf{H}^t)$ is obtained by setting the derivative with respect to $h_{kn}$ to 0,

$$\frac{\partial}{\partial h_{kn}} Q(\mathbf{H}|\mathbf{H}^{t}) = -\sum_{p=1}^{96} \ell_{pn} w_{pk}^{*} + \frac{1}{h_{kn}} \left( \sum_{p=1}^{96} v_{pn} \frac{w_{pk}^{*} h_{kn}^{t}}{\sum_{j=1}^{K} w_{pj}^{*} h_{jn}^{t}} \right) = 0,$$

and the updated activity value $h_{kn}^{t+1}$ is given by:

$$h_{kn}^{t+1} = h_{kn}^{t} \frac{\sum_{p=1}^{96} v_{pn} \left( \frac{w_{pk}^{*}}{\sum_{j=1}^{K} w_{pj}^{*} h_{jn}^{t}} \right)}{\sum_{p=1}^{96} \ell_{pn} w_{pk}^{*}}.$$

The M-step update for each signature activity depends only on the current activity values from the same tumor. Therefore, although the EM algorithm updates the entire activity matrix simultaneously, it is effectively equivalent to updating each tumor's activities one at a time. This property allows SATS to estimate signature activities for a single tumor or a small subset of samples without requiring joint modeling across the entire cohort.

For the analyses reported here, each refitting run used 50 random starts, a maximum of 5,000 iterations per start and a convergence tolerance of $1 \times 10^{-5}$, defined by the relative change in normalized Poisson log-likelihood between successive iterations. The solution with the highest final log-likelihood across random starts was retained for burden estimation.

Signature burdens were calculated as the expected number of mutations attributed to each signature in each tumor after EM convergence. The burden combines the estimated signature activity, the reference TMB signature profile and the panel-specific opportunity for the tumor. This definition allows SATS to partition the observed panel mutation burden into signature-specific burdens while accounting for differences in panel size and context composition.

**Generation and validation of the targeted-sequencing signature catalogue**

The pan-cancer targeted-sequencing mutational-signature catalogue was constructed by applying the SATS workflow separately within each cancer category used for downstream analysis in GENIE version 13.0-public. For each cancer category, SATS extracted de novo TMB signatures from grouped tumors, mapped the de novo profiles to COSMIC reference TMB signatures by recurrent pNNLS and then refitted the mapped signature set to individual tumors by the EM algorithm. SBS and DBS catalogues were analyzed separately because they use different mutation-type vocabularies and context-opportunity matrices.

Reproducibility was evaluated in held-out and external targeted-sequencing cohorts using the same analytical workflow. GENIE version 17.0 was downloaded from Synapse, and samples already present in GENIE version 13.0-public were excluded before validation. The held-out GENIE analysis focused on lung, colorectal and breast cancers, which had sufficiently large sample sizes.

Independent validation cohorts included lung and colorectal tumors from OrigiMed and breast tumors from METABRIC and FUSCC/Fudan. To accommodate smaller validation cohorts, tumors were grouped into sets of 50 tumors, and the analysis was repeated 10 times with different random seeds. Hypermutated tumors were excluded from OrigiMed and FUSCC validation analyses, and FUSCC mutations consistent with 8-oxo-guanine sequencing artefacts were removed before signature analysis.

### Shannon equitability index of TMB mutational-signature profiles

We quantified the diversity, or "flatness", of each TMB mutational-signature profile using the Shannon equitability index. This index measures how evenly the 96 SBS mutation types are represented within a signature and was calculated as follows:

$$Shannon\ equitability\ index\ = -\frac{\sum_{p=1}^{96} w_p log(w_p)}{\log(96)},$$

where $w_p$ denotes the normalized level of signature profile at the $p$th mutation type, such that the total sum across all 96 mutation types is 1.

The Shannon equitability index ranges from 0 to 1. Higher values indicate a more even distribution of mutation types, with a value of 1 corresponding to a completely flat profile in which all mutation types contribute equally. Values approaching 0 indicate a spiked profile dominated by one mutation type.

Several SBS signatures illustrate this contrast. SBS1 is strongly enriched for C>T substitutions at NCG trinucleotides and has a Shannon equitability index of 0.317, whereas SBS10a is dominated by T[C>A]T substitutions and has an index of 0.192. By comparison, SBS3, SBS5 and SBS40 are distributed more evenly across mutation types, with indices of 0.974, 0.903 and 0.969, respectively, and were therefore treated as relatively flat signatures.

### Generation of pseudo-targeted sequencing data

We generated pseudo-targeted sequencing datasets to evaluate how panel design, cohort size and signature properties affect mutational-signature detection. Somatic SBSs were subsampled from large-scale WES and WGS datasets, including TCGA WES studies[8,56] and the Sanger Institute BRCA560 WGS study[57]. We assumed that SBSs identified by WES or WGS would also be detectable in a targeted-sequencing experiment when they fell within the genomic regions covered by a given panel, which is reasonable because targeted panels are typically sequenced at greater depth than WES or WGS assays.

TCGA WES mutations were obtained from the MC3 mutation call set (mc3.v0.2.8.PUBLIC) through the Cancer Genome Data Portal (https://gdc.cancer.gov/about-data/publications/mc3-2017), and WGS data for the Sanger BRCA560 study were downloaded from ftp://ftp.sanger.ac.uk/pub/cancer/Nik-ZainalEtAl-560BreastGenomes. Genomic coordinates for each targeted panel were retrieved from the AACR Project GENIE panel definition file (Synapse ID: syn26706790), which records the chromosome, start and end positions of each panel-covered region.

For each WES or WGS sample and each targeted panel, we retained only SBSs that intersected the panel-covered genomic regions. These filtered mutations were used to construct a 96-class SBS mutation-type matrix that mimicked the output of targeted sequencing. This procedure produced 648 pseudo-targeted sequencing datasets across 18 TCGA cancer types and 36 targeted panels, with panel sizes ranging from 0.05 Mb to 9.95 Mb.

Each cancer type contained at least 200 samples in the original WES dataset. Because panel coverage varied, some WES samples contributed no usable mutations for a given panel, so the effective sample size was often smaller than the source cohort (Supplementary Table 8). For example, among 208 TCGA sarcoma cases, 169 and 172 cases yielded SBSs within the DFCI-ONCOPANEL-3 and MSK-IMPACT505 panels, respectively. The same panel definitions were also used to generate 36 pseudo-targeted sequencing datasets from the 560 WGS breast cancer samples.

The pseudo-targeted datasets were processed with the same 96-channel SBS encoding and panel opportunity calculations used for the GENIE targeted-sequencing analysis. This ensured that performance comparisons reflected the consequences of panel restriction rather than a change in mutation-type representation. Samples with no retained SBSs after panel intersection were not informative for signature estimation in that pseudo-panel and were therefore absent from that panel-specific mutation matrix.

**Analysis of pseudo-targeted sequencing data**

We evaluated mutational-signature detectability in panel-based sequencing by calculating the signature detection probability for common signatures. A common signature was defined as a signature that contributed at least 5% of SBSs in the corresponding TCGA WES cancer type. For each cancer type and targeted panel, the detection probability was the fraction of these common WES signatures recovered by SATS in the pseudo-targeted sequencing data.

To identify determinants of this probability, we fitted a generalized linear mixed model to the 648 pseudo-targeted sequencing datasets, corresponding to 18 TCGA cancer types and 36 panels. The response was whether a common TCGA WES signature was detected in the pseudo-targeted dataset. Fixed effects included signature flatness measured by the Shannon equitability index, signature prevalence in the original TCGA WES data measured as the percentage of total SBSs attributed to the signature, and panel size in megabases.

Cancer type was included as a random intercept to account for tumor-type-specific differences in mutation burden, cohort composition and signature composition. This model allowed us to estimate how signature shape, signature prevalence and panel coverage each contributed to signature detectability in targeted-sequencing data, while avoiding the assumption that all cancer types had the same baseline probability of detection.

**Sample-size simulations for targeted-sequencing signature detection**

We assessed the effect of sample size on signature detection from targeted-sequencing data using an in silico simulation study in breast cancer. The simulation was designed to distinguish spiky signatures, which can often be detected from fewer mutations, from flatter signatures that require larger cohorts and broader context sampling. We used the TCGA breast cancer WES dataset (Synapse: syn11726618) and selected 12 WES-derived breast cancer signature groups with at least 1% prevalence (SBS1, SBS2/13, SBS3, SBS5, SBS7a, SBS10a, SBS10b, SBS15, SBS29, SBS30, SBS44 and SBS58), based on Synapse: syn11801497. These WES-derived signature groups were refitted in the WES data to estimate the signature activity distribution used for simulation.

Signature refitting was performed using the EM algorithm to estimate the signature activity matrix from the observed mutation matrix, the WES panel context matrix and the fixed TMB signature-profile matrix. For each of 21 targeted-sequencing panels larger than 1 Mb, we simulated mutation matrices for tumor sample sizes ranging from 1,000 to 1,000,000. Mutation counts were sampled from a Poisson distribution with a mean determined by the panel-specific mutation-opportunity matrix, the fixed signature profiles and activities sampled from the estimated WES activity distribution.

Because APOBEC signatures SBS2 and SBS13 were highly correlated, they were treated as a combined SBS2/13 group during sampling. Tumors with zero simulated mutations were excluded. This procedure generated panel-specific tumor mutation matrices across sample sizes from 1,000 to 1,000,000 tumors while preserving the empirical activity structure of the WES data.

For each simulated mutation-type matrix, we applied signeR to extract de novo signatures, mapped the extracted profiles back to the original 12 reference signature groups using penalized non-negative least squares, and estimated signature activities and burdens with the EM algorithm.

For each simulated sample size, detection performance was summarized as the proportion of the 21 panels that successfully recovered each of the 12 ground-truth signature groups. We also recorded false-positive detections, defined as signatures not used to simulate the data but identified by de novo extraction and mapping. This design allowed us to separate failures caused by limited sample size from failures caused by broad candidate reference sets or similarity among flat signatures.

**Four-cancer in silico evaluation of SATS**

We performed in silico simulations to assess the ability of SATS to detect and estimate prespecified mutational signatures under realistic panel and cancer-type conditions. The simulation parameters were derived from AACR Project GENIE rather than from idealized uniform panels, so the expected mutation matrices reflected the observed panel distribution, cancer-type-specific sample sizes and estimated signature activities. When multiple flat signatures were present, such as SBS5 and SBS40 in lung cancer or SBS3, SBS5 and SBS40 in lymphoid-derived hematologic cancers, we merged them into a single flat-signature category. This choice reflects the limited ability of the AACR Project GENIE targeted-sequencing data to distinguish individual flat signatures at the available sample sizes.

For each cancer category, we computed an expected mutation matrix from the element-wise product of the panel context matrix, the cancer-specific signature profile matrix and the corresponding signature activity matrix. Both signature profiles and activities were estimated from AACR Project GENIE, so the simulations preserved the observed panel distribution and cancer-specific activity patterns.

We generated ten replicate datasets for each of four cancer categories: lung cancer, breast cancer, colorectal cancer and lymphoid-derived hematologic cancer. For each replicate, mutation counts were sampled from a Poisson distribution with the expected mutation matrix as the mean, and the number of simulated tumors matched the corresponding AACR Project GENIE cohort. SATS was then applied de novo, without access to the ground-truth labels; extracted profiles were mapped to the reference TMB signature set by recurrent pNNLS, and refitted burdens were compared with the simulated true burdens.

We evaluated two outputs from each replicate. First, extracted profiles were compared with the ground-truth signature groups to assess active-signature detection. Second, EM-refitted signature burdens were compared with the simulated true burdens to assess tumor-level burden estimation.

The lung cancer simulations were also used for subset and single-sample refitting analyses, described below, to test whether SATS could estimate signature activities when only a small number of tumors was available.

**Subset and single-sample refitting simulations**

To evaluate SATS in small-sample settings, we simulated lung cancer mutation-type matrices from GENIE-derived parameters. We used the mapped signature matrix and corresponding signature activity matrix estimated from the AACR Project GENIE lung cancer cohort, together with the panel context matrix, to compute expected mutation counts for each tumor and mutation type. Ten replicate datasets were generated from Poisson distributions with these expected counts as the means.

Each replicate was divided into 1-, 10- and 100-sample subsets, corresponding to the single-sample, 10-sample and 100-sample refitting settings shown in Supplementary Fig. 5. For each subset size, SATS estimated the signature activity matrix with the signature profiles held fixed, and the estimated signature expectation matrices were compared with the true expectations to quantify small-sample refitting accuracy.

**Comparator methods for in silico simulations**

Comparator analyses were separated into active-signature detection and burden refitting. For active-signature detection, comparator methods had to infer signatures directly from the simulated targeted-sequencing count matrices. We applied SigProfilerExtractor[2] and Mix[3] to the simulated mutation-type matrices. Because SigProfilerExtractor was computationally expensive at the full simulated sample size, samples were grouped before analysis. SigProfilerExtractor was run with default settings, with the number of extracted signatures constrained to 1 through 10; extracted de novo profiles were mapped to COSMIC reference signatures version 3.4, and SigProfilerAssignment was used to estimate the corresponding signature burdens.

In parallel, Mix was applied to the original ungrouped mutation matrices. Mix simultaneously clusters samples and learns mutational signatures; we evaluated combinations of 1 to 20 clusters and 1 to 10 signatures, selected the optimal model using the Bayesian Information Criterion, annotated each inferred signature by the COSMIC signature with the highest cosine similarity, and calculated signature burdens by multiplying the inferred exposures by each sample's total mutation count.

For burden-refitting comparisons, sigLASSO, ordinary NNLS, MuSiCal, deconstructSigs and SigProfilerAssignment were applied to representative 100-tumor composites from the same four-cancer simulation framework. These methods require a prespecified reference catalogue and do not perform the SATS sequence of de novo extraction, recurrent pNNLS mapping and panel-aware EM burden estimation. We therefore supplied the full TMC signature set as a specificity stress test rather than as a formal like-for-like benchmark of complete workflows.

**Validation of SATS in tumors with both WGS and targeted sequencing**

We validated SATS using 72 kidney tumors with matched WGS and targeted-sequencing data. For WGS, genomic DNA was extracted from fresh-frozen tumor tissue using the QIAamp DNA Mini Kit (Qiagen) and sequenced on the Illumina HiSeq X platform, yielding mean coverage of 65.7× for tumor samples and 40.1× for matched normal samples.
For targeted sequencing, genomic DNA was purified with Agencourt AMPure XP Reagent (Beckman Coulter, Brea, CA, USA). A custom driver-gene panel covering 1.90 Mb and targeting 254 putative cancer driver genes was used for capture with NimbleGen SeqCap EZ Choice (Roche NimbleGen, Madison, WI, USA), and libraries were sequenced on an Illumina HiSeq 4000 platform to an average depth of 500× for both tumor and matched normal tissues. Sample processing, sequencing, preprocessing, alignment and somatic mutation calling have been described previously[4].

We applied SATS, SigProfilerAssignment, Mix and deconstructSigs to the targeted-sequencing data to refit SBS1, SBS5 and SBS40 and estimate their burdens. Because SBS5 and SBS40 have similar flat profiles, we combined them into an SBS5/40 category for analysis. We then compared targeted-sequencing-derived burdens for SBS1 and SBS5/40 with the corresponding burdens from matched WGS data to assess cross-platform consistency in signature-burden estimation. For visualization, WGS-derived burdens were transformed as log1p(WGS burden), which compresses high-count samples without changing rank-based concordance. For each method and signature class, we quantified continuous concordance using Spearman correlation and leave-one-out cross-validated $R^2$ from a linear model predicting log1p(WGS burden) from targeted-sequencing burden.

**Visualization and subtype heterogeneity analyses**

To visualize tumor-level SBS signature-burden patterns, tumors within each cancer category were clustered by their signature-burden profiles using hierarchical clustering with Ward's minimum variance method. A dissimilarity threshold of 0.05 was used to define clusters. UMAP was then applied to the median signature-burden profile of each cluster, so each point in the two-dimensional embedding represents a group of tumors with similar mutational-signature composition rather than a single tumor.

Subtype-specific signature heterogeneity was evaluated with generalized linear mixed models. Signature presence was encoded as a binary outcome indicating whether the signature accounted for more than 1% of SBSs in the targeted panel. Only tumors sequenced on panels larger than 1 Mb were included.

Tumor status, race, sex and age were modeled as fixed effects when applicable, and tumor subtype and sequencing center were modeled as random effects. Panel size was included as an offset term to account for the higher probability of observing a signature on larger panels. Models were fitted with the lme4 package in R, and multiple comparisons across signatures and cancer categories were controlled by false-discovery-rate adjustment.

### Early-onset colorectal cancer analysis

Associations between mutational signatures and early-onset colorectal cancer were evaluated in 9,562 colorectal cancer patients, with early onset defined as age at sequencing younger than 50 years. Patients were stratified into non-hypermutated tumors (TMB < 10 mutations/Mb; 2,495 early-onset and 6,009 late-onset cases) and hypermutated tumors (790 early-onset and 268 late-onset cases). Within each stratum, generalized linear mixed models compared early-onset with late-onset cases while adjusting for tumor status, race, sex and selected signature-specific TMBs, including SBS1, SBS6, SBS10a, SBS10b and flat SBS signatures. Sequencing center and subtype were included as random effects.

### Survival and immunotherapy analyses

Overall-survival analyses were performed within each cancer category using mixed-effects Cox proportional hazards models. Survival time was measured in years from targeted sequencing to death or last follow-up. Fixed effects included the presence or absence of the SBS signature under evaluation, tumor status, age, race and sex when applicable. Sequencing center and tumor subtype were included as random effects, and sex was omitted for sex-specific cancer groups. Models were fitted with the coxme package in R.

For immunotherapy response, we used AACR Project GENIE Biopharma Collaborative non-small cell lung cancer data (v2.0-public). Of 1,846 patients treated at MSKCC, DFCI, VICC and UHN, 470 patients who received immune checkpoint inhibitors were analyzed.

Progression-free survival was defined as the time from immune checkpoint inhibitor initiation to radiologically or clinically confirmed progression or death, and overall survival was defined as the time from immune checkpoint inhibitor initiation to death or last follow-up. Mixed-effects Cox models included age, sex, smoking history, stage IV disease and signature-specific TMBs for SBS1, SBS2/13, SBS4, SBS89 and flat SBS signatures as fixed effects, with sequencing center and tumor subtype modeled as random effects.
